# A multi-scale cell-type resolved map of *trans*-regulatory architecture in the aging human brain

**DOI:** 10.64898/2026.11.05.26367384

**Authors:** Xuewei Cao, Yifei Wang, Haochen Sun, Ru Feng, Haoxuan Zhang, Yining Li, Hanbyul Lee, Berke Karaahmet, Boxun Li, Charles Gersbach, Gregory Crawford, Mariko Taga, Michal Schnaider Beeri, Aron S. Buchman, Hans-Ulrich Klein, Philip L. de Jager, David Bennett, The Alzheimer’s Disease Functional Genomics Consortium, Gao Wang, Kushal K. Dey

## Abstract

Mapping genetic effects on distal genes and regulatory programs is essential for understanding how genetic variation influences complex diseases; yet *trans*-QTL discovery remains limited by weak effect sizes, extensive multiple testing and poor reproducibility. Here, we introduce *TransMap*, a generalizable computational framework that combines multi-scale *trans*-QTL mapping with multi-phenotype genetic colocalization to improve the discovery and interpretation of distal regulatory variants. Beyond individual *trans*-eGenes, *TransMap* constructs local co-regulatory program phenotypes that account for background expression variation to improve the detection of weak, coordinated distal genetic effects. Across a broad range of simulation settings, *TransMap* achieved 9.2-36.7% higher *trans*-QTL discovery power than existing methods while maintaining well-controlled type I error rates. We applied *TransMap* to bulk and single-nucleus transcriptomic profiles from the aging human brain in the ADSP Functional Genomics Consortium (FunGen-AD), further integrating proteomic, glycoproteomic and metabolomic biomarker measurements (average *N=*476 donors). Genome-wide fine-mapping of *TransMap* single-gene and program-level QTL yielded variant annotations with 2.75-2.88x heritability enrichment across 57 complex diseases and traits (average *N=*307K), with complementary heritability information captured across regulatory scales. Multi-phenotype colocalization identified 49,532 *trans*-regulatory events supported by shared genetic signals with *cis*-eQTL or biomarker-QTL, including 1,825 candidate *cis–trans–biomarker* regulatory configurations. *TransMap* further revealed an asymmetric cross-cell-type regulatory architecture in which neuronal *cis*-regulatory signals preferentially associate with coordinated glial *trans*-regulatory responses, supported by co-culture experiments and spatial transcriptomics. Alzheimer’s disease (AD) GWAS-prioritized colocalization identified *trans*-regulatory associations in 115 AD-associated loci, including a *KNOP1 cis*-eQTL associated in *trans* with a microglial regulatory program containing *MS4A4E*, with additional support from statistical mediation analysis. *TransMap* provides a multi-scale, cell-type-resolved atlas of *trans*-regulatory variation and a generalizable framework for elucidating the regulatory architecture of complex diseases.

## Introduction

Most genetic variants associated with complex diseases lie in noncoding regions of the genome, yet the molecular mechanisms through which their effects propagate beyond proximal or *cis*-regulated genes to downstream cellular pathways remain incompletely understood^1–3^. *Trans*-eQTL mapping, which identifies genetic variants associated with the expression of distant genes, provides a means to uncover regulatory networks and downstream molecular processes through which disease risk may propagate^4–6^. However, *trans*-eQTL discovery remains fundamentally challenging because individual distal effects are typically weak and the genome-wide variant–gene search space imposes a substantial multiple-testing burden^4,7,8^. Large-scale studies such as eQTLGen^4,9^ and MetaBrain^10^ have advanced *trans*-eQTL discovery through extensive sample aggregation, while more recent efforts including OneK1K^11^, SingleBrain^12^ and PsychAD^13^ have extended *trans*-eQTL mapping to individual cell types in immune cells and the human brain. Nevertheless, detecting weak distal genetic effects remains particularly difficult at cell-type resolution, where molecular QTL cohorts typically have smaller effective sample sizes and greater measurement noise than large bulk-tissue studies^9^. These challenges are particularly pronounced across many disease-relevant tissues where molecular QTL datasets remain comparatively limited in sample size, including GTEx tissues^7^, pancreatic islets^14^, and kidney cohorts^15^. Recent methods, such as TransPCO^16^ and ARCHIE^17^, have sought to improve sensitivity by aggregating coordinated distal genetic effects across multiple genes, but distinguishing subtle genetically coordinated regulatory programs from background expression variation remains challenging. In parallel, the growing availability of molecular QTL maps across additional modalities, including large-scale proteomic and metabolomic resources, presents opportunities to leverage shared genetic signals across molecular phenotypes to strengthen *trans*-regulatory inference^18–22^. However, existing *trans*-QTL mapping approaches have not systematically combined gene– and program-level mapping with multi-phenotype genetic colocalization across these molecular modalities.

Here, we introduce *TransMap*, a generalizable computational framework that combines multi-scale *trans*-QTL mapping with multi-phenotype genetic colocalization to improve the discovery and interpretation of distal regulatory variants. *TransMap* captures genetic regulation in a cell type at three complementary scales: individual distal genes, local co-regulatory programs and global genetically coordinated programs. For local programs, *TransMap* uses background-whitened principal component analysis to capture coordinated regulatory variation while accounting for background expression covariance^23^, improving sensitivity to weak distal genetic effects. These complementary gene– and program-level representations are converted into quantitative molecular phenotypes for genome-wide QTL fine-mapping using SuSiE^24^. To further strengthen discovery, *TransMap* integrates cell-type resolved *trans*-eGene and *trans*-eProgram QTL with cell-type-resolved *cis*-eQTL and *trans*-acting biomarker-QTL through multi-phenotype colocalization using ColocBoost^25^. While program-level phenotypes improve the detection of coordinated distal genetic effects, multi-phenotype colocalization leverages shared genetic signals across molecular layers to support weaker *trans*-QTL associations that may not reach conventional single-phenotype significance thresholds. Matching *cis* and *trans* signals across cell types can further nominate candidate cross-cell-type regulatory configurations and potential intercellular relationships for downstream investigation.

We applied *TransMap* to whole-genome genetic variation, bulk and single-nucleus transcriptomic profiles, and proteomic, glycoproteomic and metabolomic measurements from the aging human brain in ROSMAP, as part of the ADSP Functional Genomics Consortium (FunGen-AD) (*N=*361-784 donors; **Supplementary Table 1**), encompassing individual-gene expression phenotypes and transcriptomic program-level phenotypes across 6 cell types and 3 bulk regions^25–27^. *TransMap* was deployed in both disease-agnostic (xQTL-only) and Alzheimer’s Disease (AD) GWAS-prioritized setting (GWAS-xQTL). Gene programs from *TransMap* improved the detection of coordinated distal genetic effects relative to existing program-level approaches. Across 57 relatively independent complex diseases and traits (average *N=*307K, genetic correlation *r*_g_ < 0.7)^25^, *TransMap* eGene and eProgram QTL exhibited stronger disease heritability enrichment than existing methods and captured complementary genetic information across regulatory scales. Multi-phenotype colocalization further identified *trans*-regulatory associations supported by shared genetic signals with proximal *cis*-eQTL and downstream biomarker-QTL, with program-level associations showing stronger concordance with biomarker QTL and functional gene networks. These analyses also revealed an asymmetric cross-cell-type regulatory architecture, particularly linking neuronal *cis*-regulatory signals to glial *trans*-regulatory responses, supported by orthogonal co-culture experiments and spatial transcriptomic analyses. Finally, AD-centered colocalization identified candidate *trans*-regulatory associations at 115 AD-associated loci, connecting disease risk to distal genes, programs and molecular biomarkers. These results establish *TransMap* as a generalizable framework and multi-scale, cell-type-resolved resource for investigating the regulatory architecture of complex diseases.

## Results

### Overview of the *TransMap* framework

We present *TransMap* as an end-to-end framework for defining, genetically mapping and interpreting multi-scale *trans*-acting molecular phenotypes across diverse tissues and cell types (**Figure 1a**). *TransMap* extends existing *trans*-QTL approaches along two major methodological dimensions: (i) from individual distal genes to quantitative phenotypes characterizing coordinated molecular responses within local and global gene programs, and (ii) from single-phenotype association testing to genome-wide multi-phenotype genetic colocalization across molecular modalities, leveraging additional molecular QTL maps, including *cis*-eQTL and biomarker-QTL when available, to strengthen the discovery and interpretation of distal genetic effects. *<u>TransMap</u>* can be applied either in a disease-agnostic manner, with downstream applications across a broad range of complex diseases and traits, or in a disease-specific, GWAS-prioritized manner.

**Figure 1.**
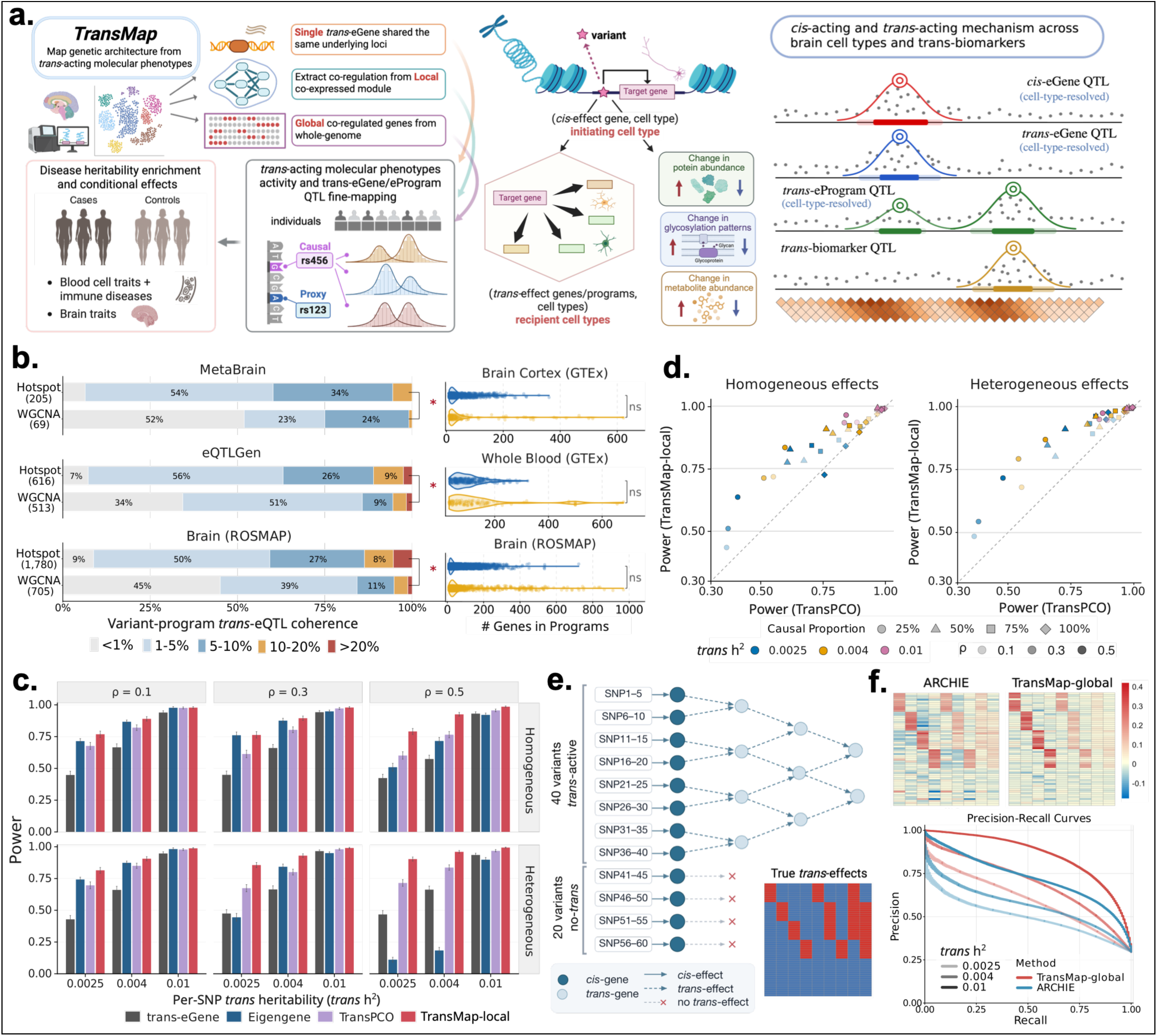
Overview of *TransMap* and benchmarking of local and global *trans*-regulatory programs. **a.** Schematic overview of *TransMap*. *Trans*-acting molecular phenotypes are defined at three scales (a single *trans* gene, a local co-expression program and a global co-regulated program), fine-mapped to identified putative causal variants from LD proxies and evaluated for disease heritability enrichment. Multi-trait colocalization links *cis*-eQTL in initiating cell types to *trans*-eQTL in recipient cell types and to downstream biomarker-QTL. **b.** *Trans*-eQTL coherence of program-active variants for Hotspot and WGCNA modules derived from GTEx brain cortex, GTEx whole blood and ROSMAP brain cortex and evaluated in MetaBrain, eQTLGen and matched ROSMAP brain cortex *trans*-eQTL, respectively (left), and the number of genes per module (right). Asterisks indicate statistical significance with FDR<0.05; ns indicates no significant difference between the program sizes from both methods. **c.** Statistical power of *TransMap-local* compared with existing methods in simulations varying residual expression correlation, per-SNP *trans* heritability and effect-sign heterogeneity across program genes. Error bars indicate 95% confidence intervals. **d.** Power comparison between *TransMap-local* (y-axis) and TransPCO (x-axis) across matched simulation conditions. Colors indicate per-SNP *trans* heritability, shapes indicate the proportion of causal *trans*-eGene in each program, and interior marks indicate the residual correlation. The dashed line indicates equal power between two methods. **e.** Simulation design for global programs following the ARCHIE framework, with 20 additional *cis*-effect only variants. **f.** Recovered variant-gene effect matrices from ARCHIE and *TransMap-global* for one simulated replicate (top), and precision-recall curves for recovering true variant-gene pairs at three per-SNP *trans* heritability (bottom). Numerical results are reported in Supplementary Data.

*TransMap* integrates whole-genome sequencing with donor-level, cell-type-resolved pseudobulk expression profiles to represent *trans* regulation at three complementary molecular scales. At the individual-gene level, *TransMap* uses normalized expression of distal genes as the conventional unit of *trans*-eQTL analysis (**Methods**). Additionally, *TransMap* defines compact local co-expression programs and broader global *trans*-regulatory programs. At the local-program level, which we refer to as *TransMap-local*, *TransMap* identifies co-expression modules using Hotspot^28^ and derives donor-level co-regulatory activity scores for each module using background-whitened principal component analysis (WPC)^23^ **(Methods)**. Unlike conventional principal component analysis, which identifies dominant axes of expression variation, WPC prioritizes program-level variation relative to a background covariance structure, thereby reducing the influence of shared expression variation unrelated to the program. For each gene program *G*, we first create 22 leave-one-chromosome-out (LOCO) programs, *G*^−*c*^, by leaving out all genes from chromosome *c*. WPC identifies a set of directions *k* for the program *G*^−*c*^ by maximizing the signal-to-noise ratio of observed program-level variance to a size-matched background residual variance:

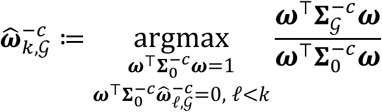

where 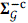 represents the expression covariance matrix corresponding to genes in *G*^−*c*^, and 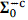 represents the estimated background residual covariance matrix (**Methods**). Let ***E*** ∈ ℝ*^n^* ^×^ *^q^* denote the centered donor-by-gene expression matrix for *n* individuals and *q* genes, after removing relevant covariates. The estimated WPC loading vectors 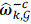 are subsequently projected onto donor-level expression profiles to generate quantitative program activity phenotypes, 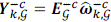. This procedure is repeated across all 22 autosomes to prevent the inclusion of genes located on the tested chromosome in the corresponding program phenotypes, thereby restricting the analysis to inter-chromosomal genetic effects. This approach can also be generalized to pre-defined local gene programs (**Methods**). At the global-program level, which we refer to as *TransMap-global, TransMap* captures genome-wide patterns of genetically coordinated expression by learning sparse variant and gene embeddings from inter-chromosomal variant-gene *trans*-associations using sparse canonical correlation analysis (sCCA)^29^ (**Methods**). The resulting gene embeddings are projected onto donor-level expression profiles in a LOCO manner to derive global program activity phenotypes.

*TransMap* also moves beyond *trans*-eQTL mapping of preselected variants to generate genome-wide, fine-mapped annotations of *trans*-regulatory potential. For each individual-gene or program-level *trans*-acting phenotype (denoted by *trans-eGene* and *trans-eProgram* respectively), *TransMap* performs genome-wide fine-mapping using the SuSiE (Sum of Single Effects)^24^ model across 1,361 approximately independent LD blocks^26^, yielding 95% credible sets (CS) and variant-level posterior inclusion probabilities (PIP) (**Methods**). Unlike previous large-scale *trans*-eQTL studies, such as eQTLGen Phase II, which restricted fine-mapping to loci with significant associations^9^, *TransMap* performs genome-wide fine-mapping across individual-gene and program-level phenotypes, retaining posterior evidence at loci that do not reach conventional association significance thresholds. This strategy is motivated by recent studies demonstrating the utility of genome-wide fine-mapping for characterizing polygenic genetic architectures and informing downstream genetic analyses^30,31^. For each *trans*-acting phenotype, we define significant *trans*-QTL signals as 95% fine-mapping credible sets containing at least one variant passing an FDR < 5% threshold based on the univariate association p-values computed for SuSiE fine-mapping, with FDR correction performed separately for individual-gene and program-level phenotypes (**Methods**). Additionally, we define genome-wide variant annotations using the maximum PIP (*trans*-MaxPIP)^32^ across the corresponding *trans* phenotypes, constructing both cell-type-specific and cross-cell-type annotations. We evaluate the disease relevance of these annotations across 57 complex diseases and traits using stratified LD score regression (S-LDSC)^33,34^ (average *N=*307K, genetic correlation *r*_g_ < 0.7), estimating heritability enrichment and standardized effect sizes after conditioning on 53 baseline annotations comprising coding, conserved and tissue-aggregated epigenomic annotations^25,34^ (**Methods**). Following previous analyses of cell-type-specific functional annotations^35,36^, we did not additionally condition on the LD-related annotations included in the baseline-LD model^37^ which capture aspects of negative selection and may attenuate cell-type-specific heritability signals^38^. We further perform joint heritability analyses of individual-gene and program-level *trans*-MaxPIP annotations, along with *cis*-eQTL-derived MaxPIP annotations, to assess whether distinct scales of distal regulation capture complementary disease-relevant genetic information beyond proximal *cis*-regulatory annotations.

Finally, *TransMap* leverages multi-phenotype genetic colocalization using ColocBoost^25^ to integrate cell-type-resolved *trans*-eGene and *trans*-eProgram QTL with proximal *cis*-eQTL and additional biomarker-QTL, thereby improving both *trans*-QTL discovery and functional interpretation (**Methods**, **Figure 1a**). Because proximal *cis*-eQTL and distal *trans*-QTL can differ substantially in effect size and statistical power, we perform separate genome-wide colocalization analyses: (i) gene-level *cis*-eQTL colocalization across cell types, and (ii) LD-block-level colocalization of *trans*-eGene, *trans*-eProgram and biomarker QTL across molecular phenotypes and cell types. We subsequently integrate events from these analyses based on shared genetic signals. Each colocalization event is characterized by this integrated colocalization confidence set (CoS) and an associated configuration of molecular phenotypes sharing the genetic signal. ColocBoost also provides a variant-level colocalization probability (VCP) in alignment with the posterior inclusion probability (PIP) in standard fine-mapping (**Methods**). This approach provides convergent genetic evidence for distal regulatory associations, including weak *trans*-QTL associations that may not achieve conventional single-phenotype significance thresholds. In the FunGen-AD application, we integrate cell-type-resolved *trans*-QTL and *cis*-eQTL across 6 cell types and 3 bulk regions with molecular QTL for proteins, glycoproteins and metabolites measured in the aging human brain cortex^26,27^. Using shared distal genetic signals through genome-wide multi-phenotype colocalization, we nominate candidate *cis–trans* and *cis–trans–biomarker* regulatory configurations. We define the cell types associated with proximal *cis*-regulated genes as the putative “*initiating cell types*” and those associated with distal *trans*-eGenes or *trans*-ePrograms as the putative “*recipient cell types*” (**Figure 1a**). These designations describe the positions of cell types within candidate regulatory configurations rather than experimentally established directions of signaling. This framework therefore enables the characterization of shared genetic associations across molecular layers and cellular contexts, extending conventional bulk *cis–trans* regulatory mapping. However, colocalization does not establish that a *cis*-regulated gene causally mediates a distal effect or that associations across cell types reflect direct intercellular signaling. Subsequent pathway-level, spatial and co-culture experimental analyses provide orthogonal evidence for the biological coherence, anatomical organization and cellular interaction signatures of the nominated configurations. The AD GWAS-prioritized ColocBoost analysis further assesses whether disease-associated genetic signals are shared with proximal *cis*-eQTL, distal *trans*-QTL or biomarker-QTL, thereby nominating candidate genes, programs, molecular biomarkers and cellular contexts for downstream functional investigation of AD susceptibility loci.

The *TransMap* framework is implemented as a command-line interface with modular components for *trans-* phenotype construction, QTL fine-mapping, and multi-phenotype colocalization (see **Code Availability**). Summary statistics from all program-level QTL maps and colocalization results are made publicly available to facilitate downstream analyses and broader scientific use (see **Data Availability**).

### Local and global *trans*-regulatory programs capture reproducible genetic effects

Having defined local and global program-level phenotypes, we first evaluated whether these representations capture coordinated distal genetic effects, using independent *trans*-eQTL resources and simulation benchmarks. For local programs, we first asked whether Hotspot-derived co-expression modules grouped genes exhibiting shared *trans*-eQTL associations. We quantified “*trans*-eQTL coherence” for each significant variant–program pair as the proportion of constituent genes exhibiting suggestive association with the same distal variant (individual-gene association p-value< 10^−5^) (**Methods**). Higher coherence therefore indicates that a program-level genetic association is supported by a larger fraction of its constituent genes, reflecting tighter coordination of distal genetic associations within the program. Hotspot programs derived from GTEx brain cortex and whole blood expression data showed 1.6-fold and 1.8-fold higher mean *trans*-eQTL coherence compared to WGCNA programs when evaluated against independent MetaBrain and eQTLGen bulk *trans*-eQTL associations respectively (**Figure 1b**). Consistent improvements were observed in the ROSMAP cohort, where co-expression programs and *trans*-eQTL maps were derived from the same single-cell transcriptomic resource. Hotspot programs showed 1.3–2.9-fold higher mean coherence across cell types than WGCNA (**Figure 1b; Supplementary Figures 1a-b**). These differences were not explained by program size, as Hotspot and WGCNA programs had comparable mean sizes and the higher *trans*-eQTL coherence of Hotspot persisted after matching programs by size (**Figure 1b**).

We next benchmarked the WPC-derived local program activity phenotypes (*TransMap-local)* across a range of simulated gene program architectures varying (i) program size, (ii) sample size, (iii) the proportion of causal *trans*-genes, (iv) AR(1) residual expression covariance structure with varying correlation parameter *ρ*, (v) per-SNP *trans* heritability based on causal effect sizes, and (vi) the sign heterogeneity of *trans*-eQTL effect sizes for genes in the program (**Supplementary Figure 1c**; **Methods**). In each simulation, genetic effects propagated through a proximal *cis*-gene to a prespecified subset of distal genes within a program, with effect sizes calibrated to span low-, intermediate– and high *trans* heritability regimes^39^ (**Methods)**. We benchmarked *TransMap-local* against TransPCO^16^, Eigengene^40^ and conventional single gene *trans*-eQTL approach (*trans*-eGene). We evaluated statistical power using Bonferroni-corrected, method-specific significance thresholds, and assessed type I error (T1E) under the 0%-causal null across 1,000 simulation replicates (**Methods**). Across many tested simulation scenarios, *TransMap-local* achieved the highest power while exhibiting comparable or lower T1E than competing methods (**Figure 1c, Supplementary Figure 1d-e**). **Figure 1c** illustrates a simulation setting with a 20-gene program containing 50% causal *trans*-genes, 1,000 samples, varying degrees of AR(1) residual expression correlation, and either homogeneous genetic effects (all causal genes affected in the same direction) or heterogeneous effects (equal proportions of positive and negative effects). Under sign heterogeneity, *TransMap-local* at low per-SNP *trans* heritability (0.0025) achieved 81.6% power, representing a 9.2%-36.7% improvement over competing approaches (**Figure 1c**). Under the sign homogeneous setting, with high per-SNP *trans* heritability and low residual correlation, *TransMap-local* performed similarly to Eigengene; however, its advantage increased when genetic effects were weakly homogeneous or sign heterogeneous. This pattern is consistent with the ability of background whitening and multiple retained WPCs to improve sensitivity to weak *trans*-regulatory effects and capture heterogeneous regulatory architectures, respectively, that may not be adequately represented by the first conventional principal component used in Eigengene. We further examined the effects of residual expression correlation and genetic-effect heterogeneity in direct comparisons with TransPCO, the strongest competing program-level method, which integrates association evidence across multiple conventional principal components but does not explicitly perform background covariance whitening (**Figure 1d**). For 68 out of 72 different simulation settings involving varying causal *trans* gene proportion, per-SNP *trans* heritability, residual correlation and sign heterogeneity, *TransMap-local* exhibited higher power compared to TransPCO. The largest power gains were observed under lower per-SNP *trans* heritability, consistent with improved sensitivity to weak distal genetic effects. Under the 0%-causal null, *TransMap-local* maintained well-calibrated T1E across the evaluated simulation settings, whereas TransPCO showed modest inflation under specific architectures (**Supplementary Figure 1e**).

We next evaluated whether *TransMap-global* improves the recovery of genome-wide variant–gene *trans*-regulatory relationships, complementing the *trans*-QTL detection benchmarks for *TransMap-local*. We benchmarked *TransMap-global* against ARCHIE using simulations that preserved the *cis–trans* directional regulatory graph and core parameter configuration of the ARCHIE simulation design^17^ (**Figure 2e**). We sampled allele frequencies of the simulated variants from ROSMAP WGS data to reflect the distribution of common and low-frequency variants observed in a population cohort (**Methods**, **Supplementary Figure 2a**) and varied two aspects of the simulation architectures: (i) layer-specific *trans* heritability and (iii) embedding sparsity (**Methods**). In these simulations, genetic effects propagated from proximal *cis*-genes through successive layers of distal *trans*-genes, with effect sizes calibrated to span low-, intermediate– and high– *trans* heritability regimes^39^ (**Methods**). We assessed recovery of the simulated variant–gene *trans* effects using pair-level precision–recall curves (AUPRC), where precision was defined as the proportion of selected variant–gene pairs corresponding to true simulated *trans* effects, whereas recall was defined as the proportion of all true variant–gene pairs recovered (**Methods**). *TransMap-global* achieved 18.2%–22.5% higher pair-level AUPRC than ARCHIE across different SNR levels and recovered up to 0.41 higher recall of variant–gene pairs at 80% precision **(Figure 1f).** This advantage persisted after re-tuning ARCHIE over the same sparsity-parameter grid, with *TransMap-global* achieving 15.9%-25.8% higher pair-level AUPRC across different SNR levels and sparsity settings (**Supplementary Figure 2b**). Notably, the relative AUPRC improvement over ARCHIE was greater for low-frequency variants compared to common variants (**Supplementary Figure 2c**).

**Figure 2.**
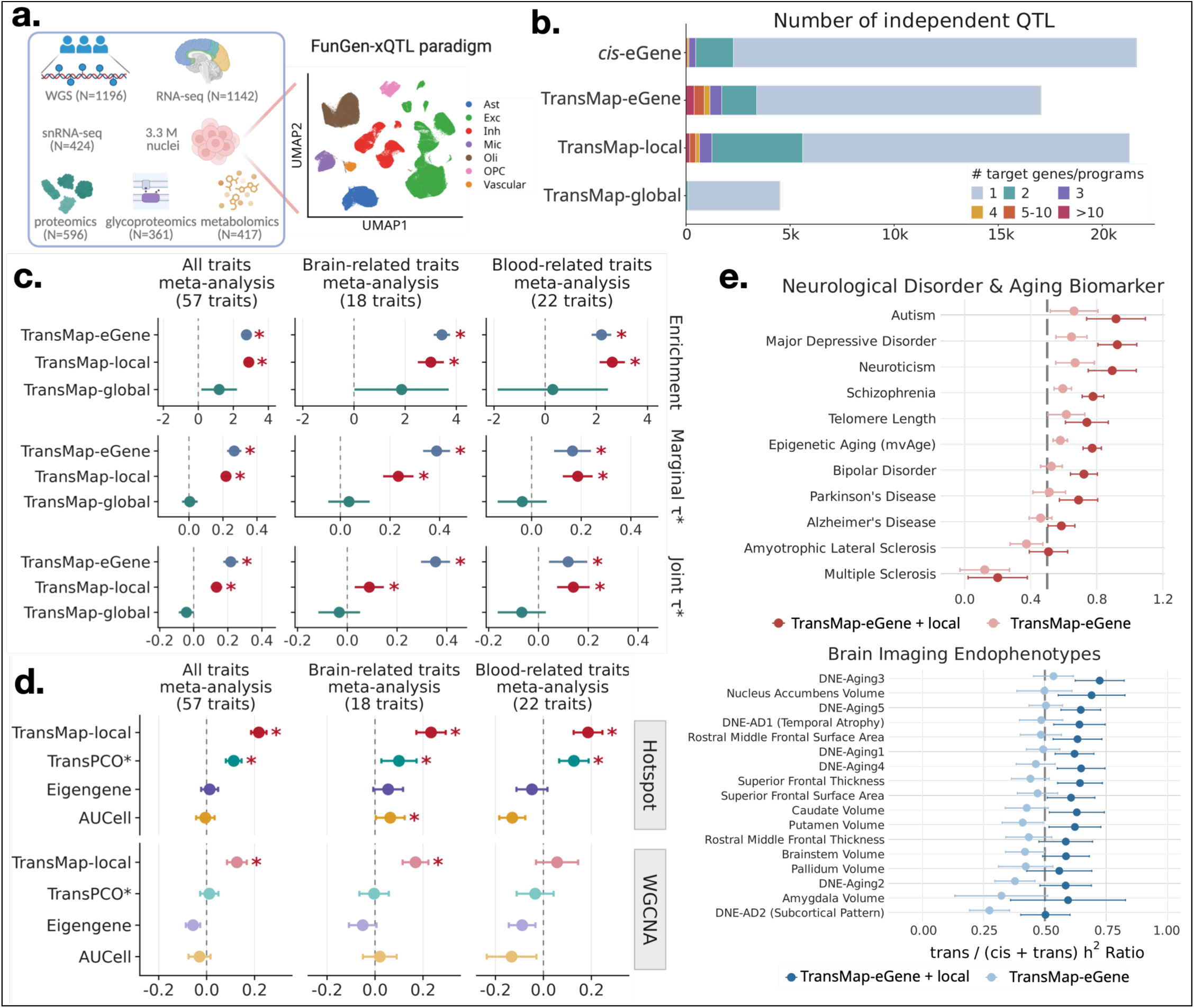
Genome-wide *trans*-eQTL fine-mapping and disease heritability analyses across three *trans*-regulatory scales from *TransMap*. **a.** Overview of ROSMAP multi-omics data, including whole-genome sequencing, bulk and single-nucleus RNA-seq, proteomics, glycoproteomics, and metabolomics. UMAP shows nuclei colored by cell types. **b.** Number of significant QTL at FDR<5% for *cis-*eGene and for *TransMap-eGene*, *TransMap-local* and *TransMap-global*, colored by the number of target genes or programs per QTL. **c.** Heritability enrichment, marginal *τ*\*, and joint *τ*\* of *trans*-MaxPIP annotations from three *trans*-regulatory scales defined in *TransMap*. **d.** Marginal *τ*\* of *trans-*MaxPIP annotations from *TransMap-local*, TransPCO, Eigengene, and AUCell representations built on Hotspot (top) and WGCNA (bottom) modules. **e.** Proportion of total explained heritability attributable to *trans*-eQTL annotations, *trans/(cis+trans)*, for 11 neurological disorders and aging biomarkers (top) and 17 brain imaging-derived endophenotypes (bottom), using *TransMap-eGene* alone (light) or combined with *TransMap-local* (dark). Dashed line denotes equal *cis* and *trans* contributions. In **c-d**, results are meta-analyzed across 57 complex traits and subsets of 18 brain-related and 22 blood-related traits. Asterisks indicate statistically significant at FDR<5%. Error bars indicate 95% confidence intervals. Numerical results are reported in Supplementary Data.

### Multi-scale *trans*-regulatory mapping reveals complementary disease genetic architectures

We next applied *TransMap* to bulk and single-nucleus RNA-sequencing profiles across 6 brain cell types and 3 brain cortical regions from the aging brain cortex in the ROSMAP cohort (average *N* = 481, **Figure 2a**, **Supplementary Table 1,** see **Supplementary Figure 3a** and **Supplementary Notes S1-S2** for technical choices). Across cell types and contexts, we identified 17,054 significant *TransMap-eGene* QTL associations across 16,901 genes, 21,315 significant *TransMap-local* QTL associations across 6,453 Hotspot co-expression programs and 4,507 significant *TransMap-global* QTL associations across 2,700 global programs (FDR < 5%, **Methods**, **Figure 2b**). Among significant *TransMap-eGene* QTL associations, 19.9% involved genetic signals associated with multiple distal genes, compared with 10.5% of *cis*-eQTL associations involving multiple proximal genes. Similarly, 26.4% of *TransMap-local* QTL associations involved multiple programs. Given the large number of correlated molecular phenotypes and genome-wide genetic associations evaluated in *trans*-QTL mapping, we independently assessed the empirical calibration of the nominal FDR-controlled discoveries using genotype-label permutations. The empirical FDR ranged 4.8%-9.2% in bulk contexts and ranged 7.7% to 14.9% across six cell types (**Supplementary Figure 3b**), indicating that conventional significance thresholds did not uniformly achieve nominal FDR control for single-phenotype *trans*-QTL mapping. These findings motivate the integration of complementary molecular-QTL evidence through multi-phenotype colocalization to provide additional statistical support for weak distal genetic associations, as evaluated in subsequent sections.

We generated 3 genome-wide variant-level *trans*-MaxPIP annotations separately for individual-gene (*TransMap-eGene*-MaxPIP), local-program (*TransMap-local*-MaxPIP) and global-program (*TransMap-global*-MaxPIP) phenotypes by aggregating fine-mapping posterior inclusion probabilities across phenotypes and cell types (**Methods**). We evaluated their disease relevance using two S-LDSC metrics: heritability enrichment and standardized effect size (*τ*\*), the latter quantifying annotation informativeness conditional on other annotations in the model. In the meta-analysis across 57 traits, 2 out of 3 annotations (*TransMap-eGene*-MaxPIP and *TransMap-local*-MaxPIP) showed high heritability enrichment (2.75x-2.88x) (p-value ranging from 2.9 × 10^−19^ to 4.3 × 10^−16^) and *τ*\* (0.22-0.27) (p-value ranging from 6.5 × 10^−38^ to 3.5 × 10^−33^) conditional on the baseline annotations (**Figure 2c**). Both annotations showed significant heritability enrichment and *τ*\* when meta-analyzed across 18 brain-related traits and 22 blood-related traits, and remained significant when modeled jointly and consistent results were observed in brain– and blood-related trait meta-analyses (**Figure 2c**). These results indicate that individual distal genes and compact local programs capture complementary components of complex-trait genetic architecture, whereas broad global programs contribute no detectable heritability beyond baseline annotations.

We next benchmarked the disease relevance of *TransMap* annotations against existing approaches. At the local program scale, we compared *TransMap-local*-MaxPIP annotation against analogous *trans*-MaxPIP annotations generated using Eigengene, TransPCO and AUCell approaches (**Methods**). Because TransPCO combines multiple principal components (PC)-level association statistics using an ACAT omnibus test, we adapted it for fine-mapping by separately fine-mapping retained PCs and aggregating their PIPs into a MaxPIP annotation (*TransPCO*-*MaxPIP; **Methods**). *TransMap-local*-MaxPIP consistently exhibited 1.6x-3.0x higher heritability enrichment than the competing annotations across all-trait, blood-related and brain-related trait meta-analyses (**Figure 2d**, **Supplementary Figure 4a)**. Both *TransMap-local*-MaxPIP and TransPCO*-MaxPIP showed significant heritability information (*τ*\*) in meta-analyses across these trait categories. Notably, *TransMap-local*-MaxPIP exhibited 1.5x-2.3x higher *τ*\* compared to TransPCO*-MaxPIP, and was the only annotation retaining significant positive conditional *τ*\* when the two annotations were jointly included in the heritability model (**Figure 2d, Supplementary Figure 4a).** Besides the fine-mapping based comparison, we generated native TransPCO variant-level associations for 43,090 xQTL-informed independent variants (*r*^2^<0.1, **Supplementary Note S3**), and compared heritability information against *TransMap-local*-MaxPIP restricted to these variants. *TransMap-local*-MaxPIP retained its improved performance in this comparison (**Supplementary Figure 4b).** When WGCNA programs were used instead of Hotspot, *TransMap-local*-MaxPIP showed 13.9% lower heritability enrichment and 42.1% lower marginal *τ*\* across the meta-analyses (**Figure 2d, Supplementary Figure 4a**). Nevertheless, WPC-based *TransMap-local* annotations remained more disease-informative than competing program-level approaches when applied to the same WGCNA programs, indicating that the advantage of background-whitened program representations is not restricted to Hotspot-derived programs. At global program scale, we benchmarked *TransMap-global* against ARCHIE^17^. Neither *TransMap-global*-MaxPIP or ARCHIE-MaxPIP showed significant heritability signals across all three meta-analyses (**Supplementary Figure 4c**). Therefore, we restricted all downstream program analyses to local Hotspot programs. At individual gene scale, *TransMap-eGene*-MaxPIP annotation showed 1.95x higher *τ*\* in all traits meta-analysis compared to fine-mapping centered around significant *trans*-eQTL at FDR<5% as performed in eQTLGen Phase II^9^, while marginally associated *trans*-eQTL (p-value< 10^−5^) did not show a significant *τ*\* (**Supplementary Figure 4d**). In a joint S-LDSC model, *TransMap-eGene*-MaxPIP remained the only annotation with a significant positive *τ*\* (*τ*\*=0.35, p-value=6.6 × 10^−44^) (**Supplementary Figure 4d**). Thus, retaining genome-wide fine-mapping posterior evidence captured distributed disease-relevant *trans*-regulatory information that was lost when downstream analyses were restricted to conventionally significant *trans*-eQTL; suggesting hidden heritability information.

We compared the proportion of trait heritability captured by *cis*-MaxPIP annotations, defined as the maximum fine-mapping PIP for each variant across *cis*-eGenes and cell types^25^, with that captured by individual-gene (*TransMap-eGene*-MaxPIP) and local-program (*TransMap-local*-MaxPIP) *trans*-QTL annotations. Across 28 brain-related traits^26,41^ (**Supplementary Table 4**), individual-gene *trans*-MaxPIP annotations captured a greater proportion of heritability than *cis*-MaxPIP annotations for 9 of 28 traits. When individual-gene and local-program *trans* annotations were combined, this increased to 26 of 28 traits (**Figure 2e; Supplementary Figure 4e**), highlighting the additional disease-heritability information captured when local-program annotations are combined with individual-gene *trans*-QTL annotations. This pattern was particularly pronounced for Autism and Schizophrenia, whereas AD and multiple sclerosis showed more comparable proportions of heritability captured by *cis*– and *trans*-MaxPIP annotations.

We performed two secondary analyses. First, we assessed whether *TransMap* yielded reproducible variant-level *trans*-MaxPIP annotations across independent datasets. For each expression profile, we randomly split donors 50:50, independently reconstructed the three scales of *trans* phenotypes, and repeated genome-wide fine-mapping in both donor subsets (**Methods**). The resulting split-level *trans*-MaxPIP annotations showed Pearson’s correlations ranging from 0.45 to 0.79 across regulatory scales (**Supplementary Figure 5a**). Despite the reduced sample sizes, the individual-gene and local-program annotations showed consistent disease-heritability signals, with significant enrichment and positive marginal *τ*^∗^ estimates in both splits (**Supplementary Figure 5b**). Second, we evaluated whether the framework could extend beyond data-derived local co-expression programs to two prespecified functional classes: 50 curated MSigDB Hallmark gene sets^16,42,43^ and cell-type-resolved SCENIC regulons^44^ (**Methods**). WPC-derived *TransMap* annotations from both program classes showed stronger disease-heritability informativeness than competing program-scoring approaches (**Supplementary Figures 5c–d**). When jointly modeled with our primary individual-gene and local-program *TransMap* annotations, most of their disease-relevant information was explained away, although Hallmark annotations retained additional conditional informativeness for blood-related traits (**Supplementary Figure 5e**).

We conclude that genome-wide *trans*-QTL fine-mapping yields reproducible and disease-informative variant annotations, with individual distal genes and compact local programs capturing complementary components of complex-trait genetic architecture.

### *Trans*-regulatory loci from *TransMap* colocalize with proximal gene regulation and downstream biomarker-QTL and exhibit distinct functional architectures

Having established the disease relevance of individual-gene and local-program *trans*-QTL annotations, we next investigated whether distal genetic associations could be linked to proximal gene regulation and downstream bulk-level biomarker molecular phenotypes (proteins, glycoproteins and metabolites) through shared genetic signals. We performed a genome-wide multi-phenotype colocalization using ColocBoost^25^ (**Methods**), representing each shared causal variant by a colocalization confidence set (CoS) if it contained at least one variant at p-value<10^−5^ and its associated configuration of molecular phenotypes sharing the genetic signal. Across the genome, we retained 49,532 colocalization events involving *trans*-eGenes or *trans*-ePrograms with support from at least one proximal *cis*-eQTL or additional biomarker-QTL (**Figure 3a, Supplementary Figure 6a**). To assess the empirical reliability of these shared-signal associations, we performed two complementary permutation analyses, yielding empirical FDR estimates of 0.4%–1.34% for genome-wide *trans*-phenotype colocalization and 0.52%–0.95% for matched *cis–trans* configurations (**Methods**, **Supplementary Figure 3b**). When evaluated against independent cell-type resolved *trans*-eQTL maps in PsychAD^13^ (*N=*1,494 samples), Colocalization-supported *trans*-eQTL associations showed 11.1– to 21.9– fold higher recall across cell types for similar precision compared to fine-mapped *trans*-eQTL hits (**Figure 3b**). Colocalization-supported *trans*-eQTL associations aggregated across cell types also showed higher recall with respect to the MetaBrain bulk *trans*-eQTL^10^ (*N=*2,683 samples) compared to fine-mapped *trans*-eQTL hits (**Supplementary Figure 6b**). The lower empirical FDR estimates and stronger external concordance support the increased reliability of colocalization-supported distal genetic associations compared to standard single-phenotype *trans*-QTL maps. Among all *cis*-eQTL signals, 26.7% were linked to at least one distal *trans*-regulatory phenotype, including 17.3% linked to both individual-gene and program-level *trans* phenotypes (**Supplementary Figure 6c**), revealing shared genetic associations across distinct scales of distal expression regulation.

**Figure 3.**
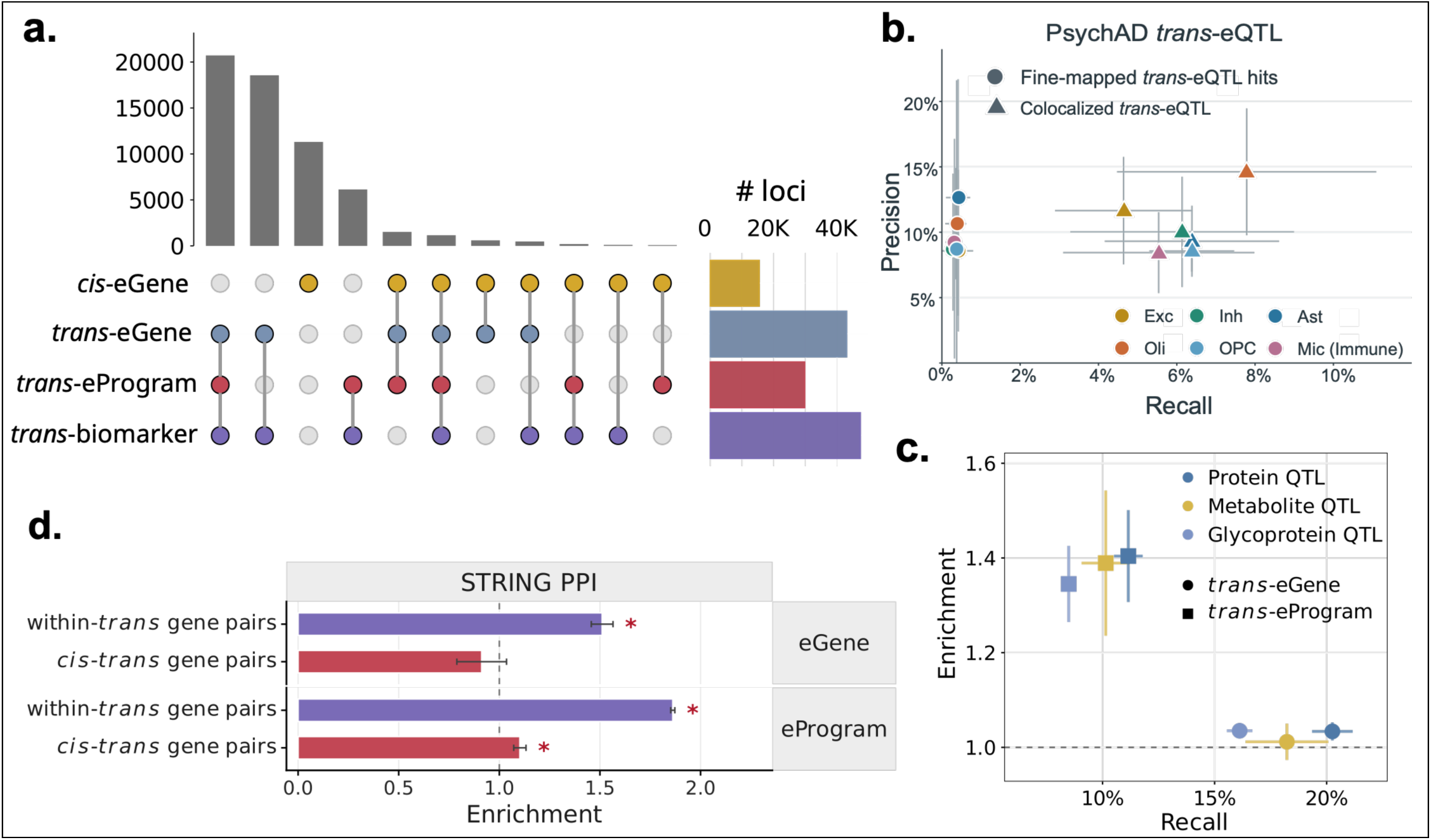
Multi-layer colocalization analysis of *trans*-eQTL with proximal *cis*-eQTL and downstream biomarker QTL. **a.** UpSet plot of colocalization events across *cis*-eGene, *trans*-eGene, *trans*-eProgram and *trans*-biomarker, restricted to events involving *trans*-QTL identified by *TransMap* supported by at least one *cis*-eQTL or biomarker-QTL. **b.** Precision and recall of *trans*-eQTL from fine-mapping hits and colocalization analyses among external PsychAD cell-type-resolved *trans*-eQTL associations with 95% confidence intervals. **c.** Enrichment and recall of *trans*-eGene QTL (circle) and *trans*-eProgram QTL (square) for overlap with each biomarker-QTL modality, relative to the genome-wide background of *trans*-QTL loci. **d.** Enrichment for functional gene properties of gene pairs within *trans*-eGenes or *trans*-eProgram (within-*trans*) and *cis*-eGene-*trans*-eGene pairs (*cis-trans*) for known protein-protein interactions in STRING database. Asterisks indicate statistical significance at FDR<5%. Error bars indicate 95% confidence intervals calculated by leave-one-chromosome-out jackknife. Numerical results are reported in Supplementary Data.

We designated colocalization events containing at least one *cis*-eGene and one *trans*-eGene or *trans*-eProgram as candidate *cis–trans* regulatory configurations (**Methods**). These included 2,198 *cis–trans* configurations and 1,825 *cis–trans–biomarker* configurations. We next examined whether program-level *trans*-QTL showed stronger genetic convergence with biomarker-QTL than individual-gene *trans-*QTL. Among *trans*-QTL loci with biomarker-QTL support, 86.4% were connected to individual *trans*-eGenes and 59.6% to *trans*-ePrograms, with substantial overlap of 46.2% across two *trans*-regulatory scales (**Supplementary Figure 6d**). Despite their lower representation among biomarker-supported loci, *trans*-eProgram QTL showed 1.35–1.40-fold enrichment for biomarker-QTL convergence compared to *trans*-eGene QTL (**Figure 3c**), indicating that program-level phenotypes preferentially identify *trans*-regulatory effects with measurable downstream molecular consequences. *Trans*-eGene and *trans*-eProgram QTL identified in glial cell types showed stronger enrichment (1.27x-1.41x, p-value < 10^−5^), whereas excitatory-neuron *trans*-QTL showed no significant enrichment across the three biomarker modalities (p-value=0.15; **Supplementary Figure 6e**).

We next examined the functional properties of genes associated with these colocalization events. We classified the colocalization events as *cis*-*only*, *cis–trans* or *trans-only* based on the representation of *cis* and *trans* phenotypes among colocalized phenotypes (**Methods**). Pairs of distal *trans*-eGenes implicated by the same colocalization event showed 1.5-fold enrichment (p-value=0.009) for known protein–protein interactions in the STRING database^45^, whereas pairs connecting proximal *cis*-eGenes and distal *trans*-eGenes showed no detectable enrichment (**Figure 3f**). Importantly, pairs of highly weighted genes within the same program exhibited an even higher 1.86-fold enrichment in protein-protein interactions. These results indicate that program-level *trans*-QTL preferentially implicate groups of functionally connected genes, complementing their stronger convergence with biomarker-QTL. We further examined whether *cis*-only, *cis–trans* and *trans*-only colocalization events exhibited distinct genomic and evolutionary properties. Genes nearest to *trans-only* QTL were significantly enriched for a curated set of 1,639 human transcription factors^46^ (1.2x-1.3x with p-value < 0.001) (**Supplementary Figure 7a**). *Cis*-only and *cis–trans* variants were located closer to transcription start sites and showed stronger evolutionary conservation^47,48^ than *trans*-only variants (**Supplementary Figures 7b–c**). However, genes nearest to *trans*-only loci showed greater enrichment for loss-of-function constraint, with 1.64-fold and 1.58-fold enrichment for loci involving *trans*-eGenes and *trans*-ePrograms, respectively (p-value < 0.001; **Supplementary Figure 7d**). These findings reveal distinct patterns of evolutionary constraint at *trans*-only loci, with weaker variant-level conservation but greater loss-of-function intolerance among nearby genes. This observation is consistent with recent reports that *trans*-regulatory loci can occur near highly constrained genes^46^, although the functional relationship between these genes and the associated distal molecular phenotypes remains unresolved.

Overall, these analyses establish that multi-phenotype colocalization provides additional statistical support for distal genetic associations and reveals complementary molecular and functional properties of individual-gene and program-level *trans*-QTL. In particular, local-program associations show stronger convergence with biomarker variation and functionally connected gene networks, supporting their utility for interpreting distal regulatory effects across molecular layers.

### Neuronal *cis* effects converge on coordinated glial *trans* responses

Having identified candidate *cis–trans* regulatory configurations, we next investigated whether shared genetic signals revealed systematic relationships between proximal and distal regulatory effects across brain cell types. Out of 4,023 *cis-trans* colocalization events, 32.3% involved distinct initiating and recipient cell types (**Figure 4a**). *Trans*-QTL represented in these colocalization events showed greater cell type specificity compared to *cis*-eQTL (**Figure 4a**). *Cis*-eQTL were distributed similarly between neuronal and glial populations, whereas *trans*-QTLs were enriched in glial cell types and relatively depleted in neurons (**Figure 4b, Supplementary Figure 8a**). Permutation tests revealed significant enrichment of neuronal-initiated, glial-recipient configurations among the initiating and recipient cell type pairs underlying the colocalization events **(Methods**, FDR<5%; **Figure 4c, Supplementary Figure 8b).** Conversely, excitatory neurons showed depleted representations among recipient cell types (0.34x-0.86x), with the strongest depletion observed for inhibitory-neuron-initiated configurations (**Figure 4d**, **Supplementary Figure 8c**). A similar pattern was observed when *trans*-eGene and *trans*-eProgram QTL were considered separately, supporting consistency across regulatory scales (**Supplementary Figures 8d-e**). We assessed whether these patterns could be explained by differences in cell-type representation as excitatory neurons comprised a large 39.8% of the 630K cells studied. We therefore performed donor-stratified subsampling of excitatory neurons, independently reconstructing their *cis*– and *trans*-eQTL maps while leaving other cell types unchanged (**Methods**). Enrichment of excitatory-neuron-initiated, glial-recipient configurations and the greater representation of excitatory neurons among initiating cell types persisted across the subsampled datasets (**Supplementary Figures 8f–g**). Fewer than 0.5% of the candidate *cis–trans* configurations overlapped detectable cell type fraction QTL, suggesting that the observed asymmetry was not predominantly explained by the cell-composition QTL identified in this cohort (**Supplementary Figure 8h-i**).

**Figure 4.**
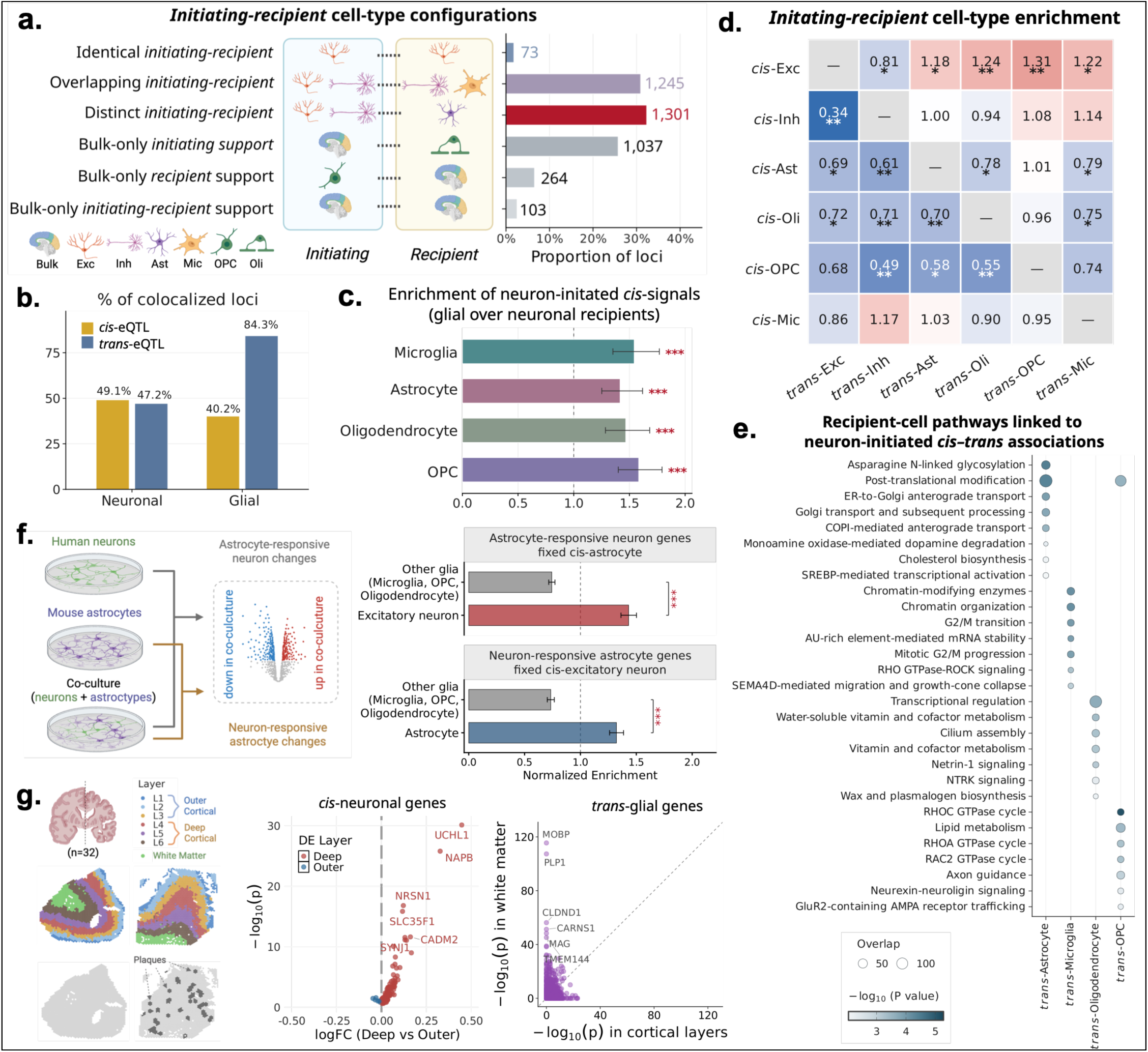
Neuron-initiated cis effects preferentially target glial cell types across genetic, pathway, and experimental evidence. **a.** Distribution of six initiating-recipient configurations of 4,023 *cis-trans* colocalization loci, defined by whether *cis*-eGene and *trans*-eGene or *trans*-eProgram cell-types are identical, overlapping or distinct, or resolved only bulk tissue on one or both sides. Numbers denote loci per configuration. **b.** Percentage of colocalized loci with *cis*-eQTL and *trans*-QTL support in neuronal (excitatory and inhibitory neurons) and glial (astrocyte, microglia, oligodendrocyte, OPC) cell types. **c.** Enrichment of glial relative to neuronal recipients among loci with neuron-initiated signals. **d.** Enrichment of each initiating (rows) and recipient (columns) cell type pair relative to shuffled null. Diagonal pairs are excluded by construction of distinct initiating-recipient configuration. **e.** Enriched reactome pathways among recipient-cell *trans*-eGenes at neuron-initiated loci. Dot size denotes the number of overlapping genes and color denotes − log_10_(*p*). **f.** Schematic of neuron-astrocyte co-culture design (left) and enrichment of recipient *trans*-eGenes among astrocyte-responsive neuronal genes for astrocyte-initiated loci and among neuron-responsive astrocytic genes for excitatory-neuron-initiated loci (right). **g.** 10X Visium sections from 32 ROSMAP DLPFC samples annotated by cortical layer, white matter and amyloid plaques (left), and spatial differential expression for *cis*-neuronal genes between deep (L4-L6) and outer (L1-L3) cortical layers (middle), and of their recipient glial *trans*-glial genes between white matter and cortical layers (right). Error bars are 95% CIs and asterisks are significance level (* FDR<5%,** FDR<1%,*** FDR<0.1%). Numerical results are reported in Supplementary Data.

We next evaluated whether neuronal-initiated, glial-recipient configurations were associated with coherent biological pathways and molecular functions. Among colocalization events involving excitatory neuron *cis* effects, astrocyte recipient *trans*-eGenes were enriched for asparagine N-linked glycosylation and post-translational protein modification (FDR<5%; **Figure 4e**). We additionally identified 26 colocalization events involving 53 glycoproteins, with enrichment for Neurofascin– and L1CAM-related interaction pathways, involving genes such as *NRCAM*, *CNTNAP1* and *NFASC* (FDR<5%, **Supplementary Figure 9a**). Of these glycoproteins, 52.5% showed STRING protein–protein interaction connections with colocalized astrocyte *trans*-eGenes (score>0.4^45^; **Supplementary Figure 9b**). Microglial *trans*-eGenes linked to excitatory-neuron *cis* effects were strongly enriched for RHO-ROCK signaling and a *SEMA4D*-associated cell-migration pathway (FDR<5%, **Figure 4e**). These pathways shared genes including *RHOB*, *ROCK2*, *LIMK1* and *MYL12B*, consistent with previously reported roles of Rho-family signaling and *SEMA4D* in microglial responses^49,50^. These findings indicate that candidate cross-cell-type configurations implicate distinct, functionally coherent glial programs associated with proximal neuronal genetic signals.

We next sought orthogonal support for the inferred neuronal–glial configurations using independent neuron– astrocyte co-culture transcriptomic data and spatial transcriptomic measurements from the ROSMAP cortex. First, in the co-culture experiment, human *Ngn2*-induced excitatory neurons were cultured with and without mouse cortical astrocytes and profiled by single-cell RNA sequencing at five time points^51^. Differentially expressed genes were pooled across time points for downstream enrichment analyses (**Methods, Figure 4f**). Excitatory-neuron *trans*-eGenes from astrocyte-initiated configurations were specifically 1.43-fold enriched among neuronal genes responsive to astrocyte co-culture **(Figure 4f).** Similarly, astrocyte *trans*-eGenes from excitatory-neuron-initiated configurations were 1.32-fold enriched among astrocytic genes responsive to neuronal co-culture. This directional correspondence was observed for both *trans*-eGenes and key driver genes of *trans-*ePrograms (**Supplementary Figure 9c**). Second, we examined the anatomical organization of neuronal–glial configurations using 10x Visium spatial transcriptomic data from 32 ROSMAP cortical tissue sections spanning different stages of AD pathology^52^ (**Methods, Figure 4g**). *Cis*-regulated neuronal genes participating in neuronal-glial configurations showed preferential expression in deep cortical layers (L4-L6) proximal to the white matter compared to the outer layers (L1-L3), whereas their associated glial *trans*-regulated genes were enriched in adjacent white matter regions (**Methods, Figure 4g, Supplementary Figure 9d**). The pattern was preserved across pathological stages and across strata of amyloid-plaque burden (**Supplementary Figure 9e**). Although spot-level resolution precludes direct inference of cell–cell contact, these results place candidate initiating neuronal genes and recipient glial responses in anatomically adjacent cortical compartments.

To illustrate how *TransMap* resolves candidate cross-cell-type regulatory configurations at individual genetic loci, we first examined an excitatory neuron-initiated (*cis*) and microglial-recipient (*trans*) colocalization event in the *SEC61G* gene locus (CoS spanning chr7:54723624-54780677). The *SEC61G cis*-eQTL in excitatory neurons colocalized with 23 microglial *trans*-eGene QTL, as well as 2 microglial *trans*-eProgram QTL (Program 31 and Program 100) (**Figure 5a**). To investigate whether variation in neuronal *SEC61G* expression statistically mediated the corresponding microglial *trans* associations, we applied structural equation model^53^ (SEM) separately for each colocalized microglial *trans*-eGene and *trans*-eProgram (**Methods**). 10 gene-level and 2 program-level associations showed significant indirect effects (p-value < 0.05 based on donor bootstrap analysis) (**Figures 5b**). Both colocalized *trans*-ePrograms also contained multiple genes with marginal associations at this locus: 2 of 49 genes in Program 31 and 2 of 18 in Program 100 had p-value< 10^−5^. Pathway analysis linked Program 31 to the TYROBP causal network in microglia (FDR=0.006) and erythropoietin-activated PI3K signaling (FDR=0.01; **Figure 5c**), whereas Program 100 was enriched for nephrin-family interactions (FDR=0.0034) and RHO GTPase-related pathways (FDR=0.004; **Supplementary Figure 10a**). To investigate the anatomical distribution of the genes implicated by this candidate configuration, we examined *SEC61G* and the associated microglial *trans*-gene *MAP4K4* in ROSMAP Visium spatial transcriptomic sections spanning different stages of AD pathology. *SEC61G* expression was more prominent across cortical layers, whereas *MAP4K4* expression was stronger in white-matter regions (**Figure 5d**), consistent with the broader spatial organization observed for neuronal *cis*– and glial *trans*-associated genes (**Figure 4g**). We next examined a complementary configuration linking neuronal *cis* regulation to astrocytic transcriptional programs and glycoprotein variation. Consistent with the enrichment of astrocytic *trans*-eGenes for N-linked glycosylation pathways, an excitatory-neuron *GDA cis*-eQTL (CoS spanning chr9:72,219,468–72,236,972) colocalized with QTL for 9 glycoproteins, including *NRCAM*, *CNTNAP1*, *NFASC* and *CNTN2*, and a 97-gene astrocytic *trans*– eProgram (**Supplementary Figure 10b**). These 9 glycoproteins were consistently enriched for neurofascin interactions and L1CAM interactions with FDR < 0.01 (**Supplementary Figure 10c**). The astrocytic *trans*-eProgram was included heavily prioritized genes *SLC3A2*, *ALG3, KDELR1* and *SEC24C*, that overlapped genes responsive to neuron-responsive astrocyte genes in the co-culture experiment (**Supplementary Figure 10d**). Although SEM found no significant indirect association through *GDA* excitatory neuron expression or astrocyte *trans*-eProgram to glycoproteins, the STRING protein-protein interaction network linked the glycoproteins to genes in astrocyte *trans*-eProgram (**Supplementary Figure 10e**).

**Figure 5.**
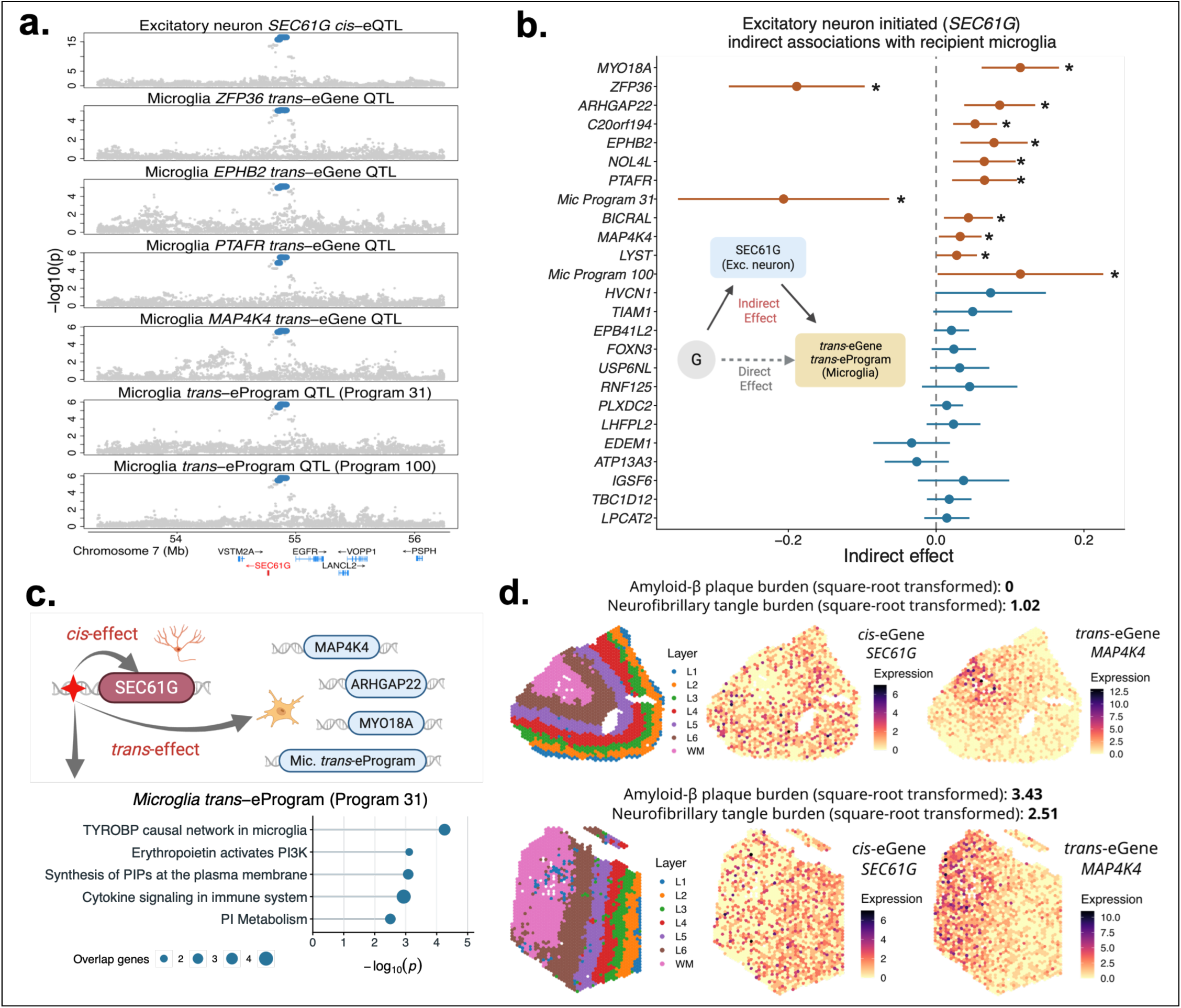
Cross-cell-type molecular associations at the *SEC61G* loci. **a.** Colocalization plot for *SEC61G cis*-eQTL, selected microglial *trans*-eGene QTL and two microglial *trans*-eProgram QTL. **b.** Structural equation model estimates of indirect effects from genotype dosage of colocalized loci through excitatory neuron *SEC61G* expression to 23 microglial *trans*-eGenes and two *trans*-ePrograms. Points show estimated indirect effects and 95% confidence intervals. Asterisks indicatxe indirect effects with p-value<0.05. **c.** Schematic of the excitatory neuron *SEC61G cis*-effect and microglial *trans*-eGenes and *trans*-ePrograms, along with the Reactome pathway enrichment for colocalized microglia *trans*-eProgram (Program 31). **d.** Cortical layer annotations and spatial expression of *SEC61G* and *MAP4K4* in two ROSMAP Visium sections with largest expressed spots.

These analyses reveal an asymmetric organization of cell-type-resolved *cis–trans* genetic associations, characterized by preferential convergence of proximal neuronal signals with functionally coherent glial distal responses. Concordance with co-culture transcriptional responses and spatial organization provides complementary biological support for these candidate cross-cell-type regulatory configurations.

### Multiscale *trans*-regulatory maps resolve the functional architecture of Alzheimer’s disease loci

We evaluated how *TransMap trans*-QTL resolve the regulatory architecture of Alzheimer’s disease. We performed AD-centered multi-phenotype colocalization analysis integrating *cis*-eGene, *trans*-eGene, *trans*-eProgram and biomarker-QTL maps with a recent AD GWAS meta-analysis^54^ (cases=128,681, controls=849,833) (**Methods**). Overall, we identified 118 colocalization events involving AD and at least one molecular*-*QTL (**Figure 6a**). For 51.7% of these events, the corresponding AD GWAS locus lacked a well-resolved 95% credible set from GWAS-only SuSiE fine-mapping, illustrating how molecular-QTL integration can nominate shared regulatory signals at loci that remain poorly resolved by GWAS fine-mapping alone. (**Supplementary Figure 11a**). 84 of these 118 colocalization events showed both *cis*– and *trans*-QTL evidence, 3 showed *cis*-only evidence and 31 showed *trans*-only evidence; 70.4% of the events involving *trans*-QTL had support across multiple *trans*-regulatory layers. *Trans*-eProgram QTL showed 1.43-fold enrichment among AD colocalization events relative to *trans*-eGene QTL (p-value= 5.4 × 10^−12^; **Figure 6b**). Among AD colocalization events involving *trans*-eGene QTL and *trans*-eProgram QTL, 74.8% also showed biomarker-QTL evidence (**Figure 6a**), revealing frequent convergence of disease association signals with distal expression phenotypes and additional molecular traits. No AD–molecular-QTL colocalization events were recovered under the genotype-permutation null, providing additional empirical support for the observed associations (**Methods**). These results highlight the utility of *trans* regulatory information specifically in the context of understanding AD disease genetics. Bulk tissue *cis*-eQTL were more frequently represented among AD colocalization events than bulk tissue *trans*-QTL (20.7% versus 4.3%; **Supplementary Figure 11b**). AD colocalization events frequently involved distinct *trans* phenotypes across multiple cellular contexts; 59 loci implicated more than three recipient cell types (**Figure 6b**). This extends prior observations that a single AD locus may engage distinct distal genes or programs across several brain cell populations^55^. Microglia was the most frequently represented initiating cell types, and astrocytes and OPC were significantly enriched among recipient cell types (**Supplementary Figures 11c-d**).

**Figure 6.**
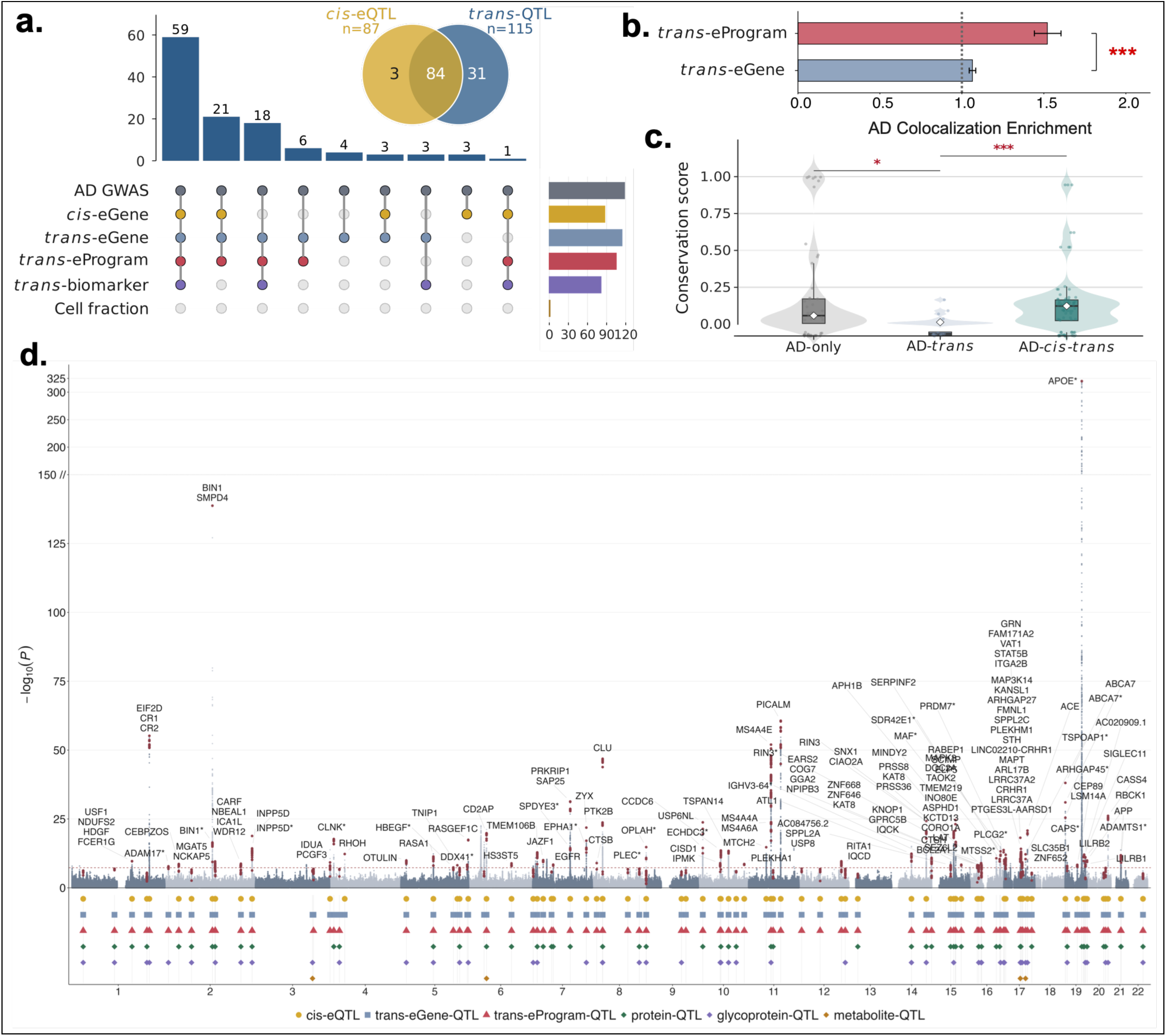
AD-centered *TransMap* colocalization identifies *cis*, *trans* and biomarker regulatory layers at AD GWAS loci. **a.** UpSet plot of 118 distinct colocalization events from AD-centered analysis, showing colocalization patterns across different modalities. **b.** Enrichment of AD-colocalization loci among *trans*-eGene and *trans*-eProgram QTL relative to the genome-wide background with 95% confidence interval. **c.** Variants conservation based on Zoonomia phyloP scores of the lead variant at each AD locus (highest MaxVCP for AD colocalized loci and MaxPIP for only AD fine-mapped loci). *AD-cis* was removed since there is only three *AD-cis* only colocalized loci. Asterisks denote significant differences between group (* FDR<5%, *** FDR<0.1%). **d.** Manhattan plot of the AD GWAS meta-analysis with colocalized loci labeled by *cis*-eGene or nearest gene at *AD-trans* only colocalized loci, and tracks below making the molecular QTL classes colocalized at each locus. Numerical results are reported in Supplementary Data.

We next investigated whether AD-colocalized variants exhibited distinct evolutionary properties according to their molecular-QTL configurations. For each AD–*cis*, AD–*trans* or combined AD–*cis–trans* colocalization event, we selected the variant with the highest variant colocalization probability (VCP) and compared its evolutionary conservation score^47,48^ (**Methods**). Variants supported by both *cis*– and *trans*-QTL colocalization showed the highest conservation scores, significantly exceeding those with *trans*-only support (p-value = 0.0007; Figure 6c). As a GWAS-only reference, we selected the highest-PIP variant from AD fine-mapping at loci without detectable *cis*– or *trans*-QTL colocalization. These AD-only variants also showed significantly greater conservation than AD– *trans*-only variants (p-value = 0.04; **Figure 6c**). To explore whether variant conservation differed across cellular configurations, we regressed conservation scores against indicators of candidate initiating cell types, recipient cell types and initiating–recipient pairs (**Methods**). No initiating cell type was significantly associated with greater conservation. Among recipient cell types, microglia and oligodendrocytes were associated with higher conservation (p-value = 0.01 for each), whereas astrocytes and inhibitory neurons were associated with lower conservation (p-value = 0.02 for each; **Supplementary Figure 11e**). Exploratory pair-level analyses also identified nominally higher conservation for selected excitatory-neuron-initiated configurations involving OPC, oligodendrocyte and microglial recipient phenotypes (**Supplementary Figure 11e**). These findings suggest that evolutionary conservation varies across AD-associated molecular QTL configurations and their implicated cell types, complementing the neuronal–glial asymmetry observed genome-wide.

We visualized the genome-wide distribution of AD-associated molecular-QTL evidence by annotating the AD GWAS Manhattan plot with colocalized *cis*– and *trans*-regulatory phenotypes (**Figure 6d**; **Supplementary Figure 11f**). This revealed substantial heterogeneity in the regulatory evidence associated with different AD loci. Some loci were predominantly supported by multiple distal regulatory phenotypes, including the *APP* region (highest VCP variant *rs432766*), whereas others showed convergence of proximal and distal associations, such as the *BIN1* locus (*rs6733839*), linking *BIN1 cis*-eQTL with an *AP4M1 trans*-eQTL, and the *APH1B* locus (*rs117618017*), linking *APH1B cis*-eQTL with 11 *trans*-eGenes, including *IKBKE*, and two *trans*-ePrograms. Several loci with comparatively modest AD association statistics also showed colocalization with multiple distal or biomarker phenotypes, illustrating the additional molecular context provided by integrated genetic associations. To assess the reproducibility of these findings across AD genetic studies, we repeated AD-focused *TransMap* colocalization using a previous GWAS meta-analysis^56^ with a smaller effective sample size (111,326 cases and 677,663 controls). This analysis replicated 69 out of the 118 colocalization events, including 44 with replicated *cis*– and *trans*-QTL evidence (**Supplementary Figure 12**).

These analyses demonstrate how multiscale, cell-type-resolved *trans*-QTL maps enhance the discovery and functional characterization of AD-associated regulatory signals by prioritizing candidate regulatory genes, coordinated expression programs, cellular contexts and biomarker-associated molecular phenotypes at AD susceptibility loci.

### Multiscale *cis–trans* regulatory maps connect genes across independent AD susceptibility loci

We next examined two case studies illustrating how multiscale *trans*-regulatory mapping can connect molecular associations across independent AD susceptibility loci and nominate candidates for experimental follow-up.

We first examined an AD-associated locus near *KNOP1* on chromosome 16^57,58^, where an astrocytic *KNOP1 cis*-eQTL colocalized with a microglial *trans*-eProgram (Program 27) containing *MS4A4E*, *MS4A6A*, *APOE* and *RIN3* (**Figure 7a**). Among the distal molecular phenotypes implicated at this locus, microglial Program 27 showed the strongest statistical support for an indirect association in SEM analysis (**Supplementary Figure 13a**). Astrocytic *KNOP1* expression exhibited the largest estimated indirect effect among the evaluated proximal candidates (**Figure 7b**). The 87-gene Program 27 included *MS4A4E*, which ranked fourth among constituent genes by marginal *trans*-association significance, and STRING analysis placed *MS4A4E* in a functionally connected component with *APOE*, *RIN3* and *MS4A6A* (**Figure 7c**). Interestingly, *MS4A4E* was also implicated as a proximal *cis*-regulated gene at the independent chromosome 11 *MS4A*-family AD locus^59^. AD-centered colocalization at this locus identified 2 distinct colocalization confidence sets with different molecular-QTL configurations (minimum between-set purity = 0.44; **Figure 7d**), consistent with distinct signals also identified by AD GWAS fine-mapping. One colocalization event included microglia-specific *cis*-effects at *MS4A4A* and *MS4A6A*, the other one included *MS4A4E cis*-eQTL in DLPFC and 9 *trans*-eGene-QTLs and 3 *trans*-eProgram– QTL in inhibitory neurons (52.2% of all colocalized *trans* phenotypes; **Supplementary Figure 13b**). STRING protein–protein interaction analysis revealed functional connections among the implicated inhibitory-neuron *trans*-eGenes and colocalized protein phenotypes (**Supplementary Figure 13c**). One of the colocalized trans– ePrograms (program 59) was enriched in *MECP2*-related transcription and protein-folding pathways, (FDR < 5%; **Supplementary Figures 13d-e**). These observations connect two independent AD susceptibility loci through *MS4A4E*, which appears as a distal member of a microglial program associated with the *KNOP1* locus and as a proximal *cis*-regulated gene at the *MS4A* locus, where the same genetic signal also implicates an inhibitory-neuron program enriched for *MECP2*-related transcription^60,61^. This cross-locus convergence implicated by *TransMap* nominates *MS4A4E* for AD susceptibility, but whether it play the key role of AD risk at *MS4A*-family locus remains to be established experimentally.

**Figure 7.**
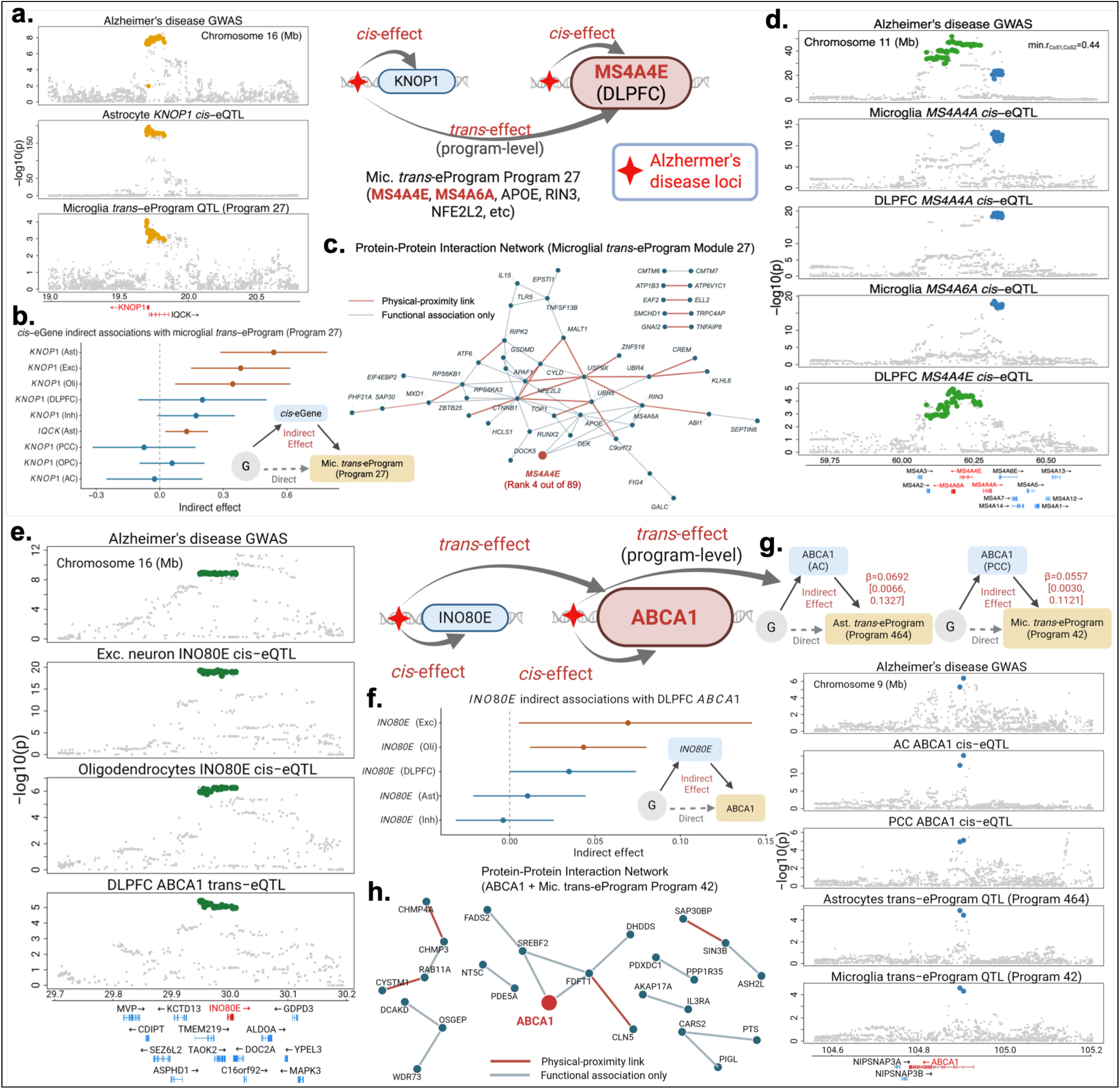
Candidate *cis-trans* regulatory paths involving *MS4A*-familiy genes and *ABCA1* at distinct AD loci. **a.** Colocalization plot at the chromosome 16 *KNOP1* locus for AD GWAS, the astrocyte *KNOP1 cis*-eQTL and the microglia *trans*-eProgram QTL. **b.** Schematic of mediation analysis model and results from genotype of colocalized locus through candidate *cis*-eGene expression to microglia *trans*-eProgram (Program 27), which includes *MS4A4E* and *MS4A6A*. Points show estimated indirect effects with 95% confidence intervals. **c.** STRING network of genes in microglia *trans*-eProgram module 27, with *MS4A4E* highlighted. **d.** Colocalization plot at the chromosome 11 *MS4A*-family gene locus for AD GWAS, including two 95% colocalization confidence sets. **e.** Colocalization plot at the chromosome 16 *INO80E* locus for AD GWAS. **f.** Schematic of mediation analysis model and results from genotype of colocalized locus through *INO80E* expression to *ABCA1* expression in DLPFC. Points show estimated indirect effects with 95% confidence intervals. **g.** Colocalization plot at the chromosome 9 *ABCA1* locus for AD GWAS. **h.** STRING PPI network of *ABCA1* (red highlighted) and colocalized microglia *trans*-eProgram.

We next examined a complementary cross-locus configuration beginning with *INO80E* at the chromosome 16p11.2 BP4–BP5 region^62,63^, a structurally complex genomic locus. At this locus, an AD-associated signal colocalized with *cis*-eQTL for ten genes across expression contexts and a DLPFC *trans*-eQTL for *ABCA1*, an established AD-associated gene at an independent chromosome 9 locus^64,65^ (**Figure 7e; Supplementary Table 7**). To prioritize candidate proximal regulators of the distal *ABCA1* association, we fitted structural equation models with *ABCA1* expression as the distal outcome (**Methods**). Among the ten proximal *cis*-eGenes, *INO80E* expression in excitatory neurons and oligodendrocytes showed nominally significant indirect associations with distal *ABCA1* expression (p-value < 0.05; **Figure 7f**). *INO80E* encodes a subunit of the INO80 chromatin remodeling complex^66,67^ and belonged to a functionally connected group of these local *cis*-eGenes in the STRING network, while they did not connect with *ABCA1* (**Supplementary Figure 14a**). We then examined indirect associations among the local *cis*-eGenes with *INO80E* by fitting additional mediations with *INO80E* alternately as mediator and outcome. These models yielded 14 and 15 indirect associations, respectively (p-value < 0.05; **Supplementary Figures 14b-c**). With *INO80E* modeled as the mediator, *MAPK3* and *KCTD13* showed the strongest indirect effects among the local genes. Although ERK1/2 and RhoA signaling have been implicated in *ABCA1* regulation in other systems^68–71^, whether *INO80E* contributes to the distal *ABCA1* association through either gene remains to be tested experimentally. At the independent chromosome 9 *ABCA1* locus, *ABCA1* instead appeared as a proximal *cis*-regulated gene, with bulk-tissue *ABCA1 cis*-eQTL colocalizing with astrocytic and microglial *trans*-eProgram QTL (**Figure 7g**). These configurations also showed nominal support for indirect associations in SEM analyses. Within the 79-gene microglial program, *FDFT1* ranked sixth by marginal *trans*-association significance and was functionally connected to *ABCA1* in the STRING interaction network (**Figure 7h**). These results link independent AD-associated loci through *ABCA1*, which appears as a distal *trans*-regulated gene associated with the *INO80E* locus and as a proximal *cis*-regulated gene connected to glial regulatory programs at its own locus.

## Discussion

*TransMap* provides a generalizable framework for performing (i) multiscale cell-type-resolved *trans*-QTL mapping, involving individual genes and gene programs, and (ii) multi-phenotype colocalization of these *trans*-QTL with cell-type-resolved *cis*-eQTL and biomarker-QTL. Applied to the aging human brain through the ADSP Functional Genomics Consortium (FunGen-AD), *TransMap* provides a cell-type-resolved atlas of *trans*-regulatory variation and their relationships with AD-associated genetic variation. The resulting maps reveal (i) complementary contributions of individual-gene and local program-level QTL annotations to complex trait heritability, (ii) stronger correspondence of program-level genetic signals with downstream biomarker-QTL and functional networks, (iii) candidate *cis–trans* regulatory configurations spanning distinct brain cell types, and (iv) expanded interpretation of AD susceptibility loci through shared genetic associations with distal genes, regulatory programs and molecular biomarkers.

A central insight from *TransMap* is that distal genetic regulation can be more sensitively detected and functionally characterized through coordinated molecular responses than through individual gene associations alone. Although a regulatory variant may exert only weak effects on individual distal genes, its effects can converge on groups of co-regulated genes, producing a more detectable program-level genetic signal. *TransMap*’s whitened PCA approach improves the detection of coordinated trans effects while accounting for background expression covariance and capturing multiple axes of regulatory variation within a program. Notably, individual-gene and local-program *trans*-QTL annotations capture complementary components of complex-trait heritability despite being derived from the same transcriptomic measurements, highlighting the value of representing distal genetic regulation at multiple molecular scales. Complementing this program-based strategy, multi-phenotype colocalization leverages shared genetic signals across molecular phenotypes to support weak *trans* associations that may not achieve conventional significance thresholds individually. More broadly, the integration of multiple molecular modalities provides complementary views of the consequences of genetic variation, allowing evidence from proximal gene regulation, distal expression programs and molecular biomarkers to jointly inform trans-regulatory inference. By connecting these complementary molecular phenotypes through shared genetic signals, *TransMap* can pinpoint candidate regulatory relationships underlying an association, providing a more detailed basis for downstream functional investigation. The stronger convergence of program-level *trans*-QTL with biomarker-QTL and functional protein networks further suggests that coordinated expression programs provide informative representations of distal genetic effects across molecular layers. Moreover, the enrichment of neuronal *cis*-regulatory signals among loci associated with glial *trans*-regulatory responses, supported by co-culture and spatial transcriptomic analyses, suggests that distal genetic regulation may involve relationships across cellular contexts rather than being restricted to the cell type containing the proximal genetic effect. This has broader implications for disease-variant interpretation, as the cellular context of proximal gene regulation may differ from those exhibiting associated distal molecular responses, as also highlighted in recent mouse models^72,73^. These findings support a multiscale and multimodal view of *trans* regulation in which genetic effects are distributed across coordinated molecular programs, molecular layers and cellular contexts.

*TransMap* provides several reusable outputs and opportunities for downstream applications in human genetics. First, its modular end-to-end pipeline for trans-acting phenotype construction, genome-wide QTL fine-mapping, and multi-phenotype colocalization, can be deployed across diverse tissues, cell types and molecular QTL cohorts. This framework may be particularly useful in tissue-specific or cell-type-resolved QTL datasets with modest sample sizes, where conventional individual-gene *trans*-QTL discovery remains underpowered. Moreover, as molecular QTL resources expand across transcriptomic, proteomic, metabolomic and other molecular measurements, integrating these complementary genetic views offers opportunities to improve the detection and functional interpretation of distal regulatory effects. Second, although the gene programs examined in this study were primarily derived from the analyzed tissues, *TransMap* can also incorporate externally defined gene programs, including those identified from mouse models or genetic perturbation experiments, to investigate their regulation by human genetic variation. Third, genome-wide fine-mapped *trans*-eGene and *trans*-eProgram QTL provide variant-level annotations that can complement *cis*-eQTL annotations in functional annotation-based heritability analyses^32,34^ and potentially inform polygenic risk prediction^74,75^ and polygenic localization methods^30^ across a broad range of complex diseases and traits. Fourth, *cis*-regulated genes and distal *trans*-eGenes or *trans*-ePrograms implicated through colocalization with disease GWAS provide specific molecular and cellular hypotheses for targeted functional validation. In particular, cell-type-resolved *cis–trans* regulatory configurations can guide the design of cell-type-specific CRISPR perturbation experiments in co-culture systems, where candidate *cis*-regulated genes are perturbed in putative initiating cell types and their predicted distal transcriptional responses are measured in recipient cell types^76^. Finally, while the present study primarily focuses on molecular assays with single readouts, we are currently extending *TransMap* to 10X scRNA+ATAC multiome datasets as part of the FunGen-AD effort to characterize distal genetic regulation across coupled molecular layers.

Our work has several limitations, representing important directions for future research. First, the substantial differences in effect sizes between proximal *cis*-eQTL and weaker distal *trans*-QTL create statistical challenges for performing untargeted multi-phenotype colocalization jointly across *cis* and *trans* phenotypes, motivating our strategy of performing separate *cis*– and *trans*-QTL colocalization analyses, followed by integration of colocalization events sharing genetic signals. Additionally, weak individual-gene cell-type-resolved *trans*-QTL associations exhibited elevated empirical false discovery rates despite nominal FDR control. In contrast, multi-phenotype colocalization yielded *trans*-regulatory associations with lower empirical false discovery rates in the evaluated permutation analyses and stronger concordance with independent *trans*-QTL resources (**Supplementary Figure 4c**). Further methodological development is needed to accommodate differences in statistical power across molecular phenotypes and enable more comprehensive joint *cis–trans* colocalization. Second, although *TransMap* identifies candidate configurations through multi-phenotype colocalization, shared genetic associations do not establish that a *cis*-regulated gene causally mediates a downstream *trans* effect or that cross-cell-type associations reflect direct intercellular signaling. Although statistical mediation, co-culture and spatial transcriptomic analyses provide complementary support for selected configurations, targeted genetic perturbation experiments will be important for establishing the causal architecture of these regulatory relationships. Third, the performance of background-whitened PCA depends on accurately estimating the background expression covariance structure. Limited sample sizes, expression sparsity and heterogeneous cellular states may compromise this estimation and affect the sensitivity and interpretability of derived programs. Fourth, strong residual correlations among molecular phenotypes can result in loss of power for large-scale multi-phenotype colocalization, as highlighted in the ColocBoost paper^25^. The residual correlations estimated in the present datasets fall within the range where statistical power and false discovery rates were generally well maintained^25^, and empirical permutation analyses further support favorable null calibration in the current application. Fifth, the present study does not systematically address cross-cohort harmonization of expression measurements, program definitions and genetic association signals, which will be necessary for integrating independently generated *trans*-QTL maps and improving portability across datasets and populations.

## Methods

### *TransMap*: an end-to-end framework for distal genetic regulation across cell types

*TransMap* is proposed as an end-to-end computational framework that represents *trans* regulation at three molecular resolutions from gene expression, including individual distal genes (*trans*-eGene) and gene programs (*trans*-eProgram). A gene program may be represented by a set of small number of genes (typically ranging from tens to hundreds of genes) or a continuous weighting of all genes genome-wide (**Figure 1a**). Each *trans*-acting gene or gene program within a cell type is encoded as a donor-level quantitative activity phenotype and carried through genome-wide LD-aware *trans*-QTL fine-mapping. These cell-type resolved *trans*-QTL are then analyzed together with cell-type-resolved *cis*-eQTL and downstream bulk tissue biomarker-QTL using multi-phenotype colocalization, which tests whether proximal *cis* signals, distal *trans* signals and downstream biomarker signals are consistent with a shared underlying genetic locus. Colocalization indicates compatibility with a shared causal variant across a set of molecular phenotypes but does not establish mediation or direct intercellular signaling. *TransMap* nominated candidate regulatory configurations represented by (*cis*-eGene, *cis*-effect cell type, *trans*-eGene/eProgram, *trans*-effect cell type, *trans*-biomarker). We term the *cis-effect* cell types as ‘*initiating cell types*’ and the distal *trans*-effect cell types as ‘*recipient cell types*’. This design allows *TransMap* to compare regulatory evidence across scales and cell types for biological characterization.

### Construction of multi-scale, cell-type-resolved *trans*-acting phenotypes

*TransMap* constructs cell-type-resolved, donor-level trans-acting molecular phenotypes from gene expression data at three resolutions: i) individual distal genes provide the most specific measured targets (*TransMap-eGene*), ii) local programs extract co-regulatory axes from compact co-expression neighborhoods (*TransMap-local*), and iii) global programs capture broader axes of genetically coordinated expression learned from genome-wide trans-association architecture (*TransMap-global*).

***TransMap-eGene.*** Let ***E*** ∈ ℝ*^n^*^×*q*^ denote the donor-level pseudo-bulk whole-genome mean-centered expression matrix for *q* genes of *n* individuals in each cell type or bulk context, after removal of relevance covariates considered in *trans*-eQTL analysis^10^ (see **Data resources and preprocessing** below). Let 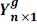 denote the standardized gene-expression vector across *n* donors for gene *g*. These phenotypes represent the conventional unit of *trans*-eQTL mapping and provide the single-gene baseline for program-level analyses.

***TransMap-local.*** *TransMap-local* converts compact co-expression genes into donor-level program activity phenotypes through two steps: (i) co-expression program detection and (ii) the program-level activity characterization across donors. To detect compact co-expression modules in each cell type or bulk context, we implement Hotspot^28^ on the expression matrix ***E***, with donor similarity defined from the entire expression space. Gene modules were identified based on gene-level local autocorrelation on this donor-neighborhood graph (n_neighbors=30) and graph-smoothed local correlations among informative genes^28^. We considered minimum module-size thresholds of 10, 30, and 50 genes to evaluate local program definitions across module granularity, from fine-grained co-expression substructures to broader co-expression genes (**Supplementary Note S4.1**).

**Let** 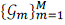 denote the *M* co-expression programs identified by Hotspot in each cell type or bulk context, where *g_m_* represents a vector of genes in program *m*. We first create 22 leave-one-chromosome-out (LOCO) programs, 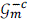, by leaving out all genes from chromosome *c*. Let 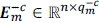 denote the donor-by-gene expression matrix for program *m*, where 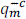 is the number of genes outside chromosome *c*. *TransMap-local* extracts donor-level co-regulatory activity from 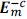 using the whitened principal component (WPC) approach^23^. Let 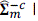 be the program-level covariance matrix for 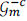, and 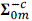 be the background residual covariance matrix matched with 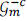. WPC estimates loading directions by maximizing module variance contrastive to the background, which maximizes signal-to-noise ratio (SNR)

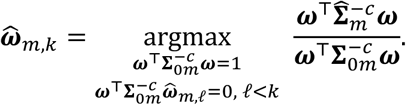

The denominator normalizes each projected score by the background covariance, whereas the numerator measures module variance along the same direction. WPC as a method parallels the ContraPC contrastive PCA approach^77^. Therefore, WPC prioritizes axes whose variation exceeds the residual background rather than axes with the largest raw expression variance. The *k*-th WPC loading vector and corresponding eigenvalue are obtained from the generalized eigenvalue problem 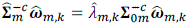. If the module covariance is fully explained by the residual background, then 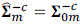 and 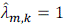 for all *k*. Thus, eigenvalues greater than 1 indicate module variance in excess of the residual co-expression background. We defined *K_m_* as the number of WPCs with 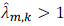, and retained these *K_m_* components for module *m*. Details on estimating 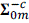, constructing empirical backgrounds, and retaining WPC components were provided in **Supplementary Note S4.2**. The corresponding activity score for *k*-th WPC in module *m* is defined as

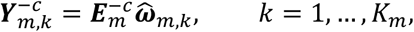

where each 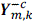 was standardized and used as a continuous *trans*-acting molecular phenotype for downstream association testing, fine-mapping, and colocalization analysis.

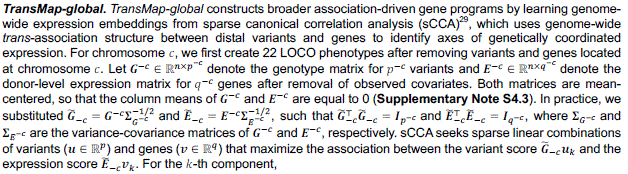

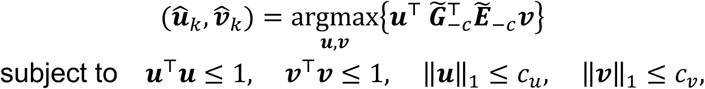

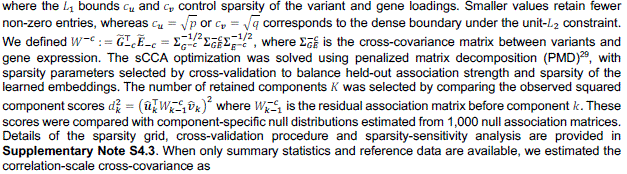

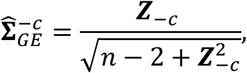

where ***Z***_−*c*_ is the *Z*-score matrix of *trans*-associations between variants and genes, and the square, division and product are elementwise. **Σ***_G_*−*c* and **Σ***_E_*−*c* can be approximated by an empirical LD matrix and a co-expression matrix from external reference panels. The derivation of 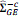 is provided in **Supplementary Note S4.3**.

The corresponding donor-level activity score for the *k*-th sCCA component is constructed by directly projecting expression onto the retained gene loading

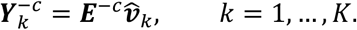

To avoid double usage of *trans* association evidence and ensure genuine *trans* effects, we implemented a LOCO strategy for chromosome-specific testing. For variants tested on chromosome *c*, only variants and genes outside chromosome *c* were used to construct *TransMap*-*global* phenotypes. In addition, when constructing 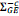, variant-gene entries from the same chromosome were set to zero according to the *trans* definition. Each 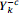 was standardized and used as a continuous *trans*-acting molecular phenotype for downstream association testing, fine-mapping and colocalization analysis.

### Genome-wide *trans*-QTL characterization through statistical fine-mapping

Statistical fine-mapping analysis provides an LD-aware approach to pinpoint putative causal variants underlying regional association signals. Each independent signal is summarized by an *α*-level credible set (CS) and variant-level posterior inclusion probabilities (PIP)^24^. *TransMap* defines an independent *trans*-QTL as a distal genetic signal represented by a 95% CS for a standardized, cell-type-resolved, donor-level *trans* molecular phenotype ***Y***^−*c*^ from statistical fine-mapping analysis. Each ***Y***^−*c*^ can represent an individual distal-gene expression trait (*TransMap-eGene*), a local WPC program score (*TransMap-local*), or a global sCCA program score (*TransMap-global*). We summarized the resulting *trans*-QTL and PIP as genome-wide variant-level *trans*-MaxPIP annotations to evaluate their enrichment for complex-trait heritability (**Methods**).

Genome-wide fine-mapping is organized across 1,361 approximately independent autosomal LD blocks^26^. Each LD block is extended by 50 kb on both sides to define a fine-mapping region and reduce boundary effects for signals near block edges. For each region on chromosome *c*, we first assembled the corresponding *trans* phenotype set across three scales by excluding genes on chromosome *c*. We then performed fine-mapping using the SuSiE (Sum of Single Effects) model^24^ with a maximum of 10 causal signals per locus and purity= 0.5. We defined a significant *trans*-QTL association as a 95% credible set (CS) from SuSIE containing at least one variant with a marginal association p-value below the method-specific threshold corresponding to FDR < 5%.

To assess the expected false discovery burden, we estimated the effective number of independent common variants (MAF > 5%) in the ROSMAP WGS reference panel as 619,380 (PLINK ––indep-pairwise 50 1 0.2)^9^. Across ∼108K individual *trans*-genes (average 12,444 genes, **Supplementary Table 1**) and ∼9K *trans*-programs (average 1,017 programs, **Supplementary Figure 1a**) genome-wide scans spanning 3 cortical regions and 6 major brain cell types, we identified 91,019 and 42,030 independent 95% credible sets whose lead variant reached p-value < 5 × 10^−8^ and p-value < 5 × 10^−K^, respectively. Under a calibrated null, we would expect approximately 3,345 (619,380 × 108K × 5 × 10^−8^) and 2,787 (619,380 × 9K × 5 × 10^−K^) false-positive credible sets at these thresholds, corresponding to approximate FDRs of 3.7% and 6.6%^9^. Because correlation among phenotypes does not alter the expected number of false positives, these estimates are approximately unbiased but do not account for residual p-value miscalibration, which we assess empirically using genotype-label permutations.

### Genome-wide QTL discovery for curated and data-driven gene programs

To enable program-level QTL discovery beyond co-expression modules, we extended *TransMap* to analyze any prespecified functional gene set. For each gene set and expression context, *TransMap* applies the WPC procedure used for *TransMap-local* to derive donor-level co-regulatory activity scores. Each retained WPC is treated as a quantitative molecular phenotype and subjected to genome-wide fine-mapping across 1,361 LD blocks^26^. For *trans*-specific mapping of variants on chromosome *c*, genes located on this chromosome are excluded before phenotype construction; complementary network-wide mapping retain all member genes to capture genetic effects on the complete program. To evaluate curated biological programs, we analyzed the 50 Hallmark gene sets from the Molecular Signatures Database (MSigDB) previously examined using TransPCO^16^, comprising 32-200 genes and provide coherent and relatively nonredundant representation of biological states and processes. To evaluate data-driven regulatory programs, we inferred context-specific TF-target regulons separately for six major brain cell types and three bulk cortical regions using SCENIC^44^: GRNBoost2^78^ was first used to estimate weighted regulatory association between human TFs and all expressed genes, after which the top highest-ranked targets per TF were partitioned according to their correlation direction and pruned using RcisTarge motif enrichment (normalized enrichment score > 3.0)^79^. The resulting motif-supported regulons and Hallmark gene sets were summarized using WPC and mapped using the same genome-wide framework. These applications establish *TransMap* as a general-purpose for converting curated or data-driven gene sets into quantitative phenotype for program-level QTL discovery.

### Multi-phenotype colocalization of cross-cell-type *cis*–*trans* regulatory paths

A *trans*-QTL can represent a downstream molecular consequence of a proximal *cis* effect, in which local genetic regulation propagates to distal molecular phenotypes within or across cell types. These effects may further converge with downstream biomarker signals measured through proteomic, glycoproteomic and metabolomic assays. Proximal *cis*-regulatory genes are relatively few and typically have stronger *cis*-QTL effects, whereas distal *trans*-regulatory phenotypes are far more numerous and individually weaker^7^. This imbalance in dimensionality and association strength can substantially limit power for a single joint colocalization analysis. We therefore implemented a two-stage genome-wide multi-phenotype colocalization strategy using ColocBoost^25^.

In the first stage, we identified colocalization among distal molecular phenotypes within the same 1,361 extended LD block regions. Candidate phenotypes comprised cell-type-resolved *trans* genes and *trans* programs together with *trans* biomarkers when available. Within each region, we retained phenotypes for which fine-mapping identified at least one 50% CS overlapping the region^26^. To reduce redundancy, highly correlated phenotypes were pruned within each phenotype class at an absolute pairwise correlation of ∣ *r* ∣≥ 0.7. ColocBoost was then applied jointly to the retained distal phenotypes. Each *trans*-only colocalization event was summarized by a 95% colocalization confidence set (CoS), variant-level variant colocalization probabilities (VCP), and a trait configuration specifying the subset of phenotypes compatible with the shared genetic signal. We retained 95% CoS as an independent locus for colocalization events when it contains at least one variant with marginal p-value < 10^−5^ for the traits in configurations.

In the second stage, we matched these distal colocalization events to events from a separate genome-wide gene-level colocalization analysis of cell-type-resolved *cis*-eQTL^26^. A distal *trans*-only colocalization event and a proximal *cis*-only colocalization event were integrated when their 95% CoS shared the same causal variants. Therefore, we define a ‘*Colocalization Event’* as this integrated colocalization confidence set and an associated configuration of molecular phenotypes sharing the genetic signal. Configurations that link one or more proximal *cis*-eGenes to one or more distal *trans*-eGenes or *trans*-ePrograms were defined as ‘*Candidate Regulatory Configurations’*, indicating the core *cis*–*trans* link and may further connect to a downstream *trans*-biomarker. The *cis*-effect cell types define ‘*initiating cell types*’, whereas the distal *trans*-effect cell types define ‘*recipient cell types*’ (**Figure 1a**). Because the two stages were connected through overlap between independently estimated colocalization events, these configurations do not represent formal joint *cis-trans* colocalization and cannot establish mediation or direct intercellular signaling. Nevertheless, this design allows *TransMap* to organize shared regulatory evidence across molecular scales and cell types for subsequent biological characterization.

### AD-prioritized multi-phenotype colocalization of *cis*– and *trans*-regulatory signals

In contrast to LD-block-based genome-wide fine-mapping and colocalization analyses, AD-prioritized colocalization was organized around topologically associating domain (TAD)-based regions containing candidate AD loci. We extracted candidate AD loci curated by the FunGen-xQTL project^26^ and mapped their constituent variants by genomic position to a genome-wide reference set of 1,328 TAD regions^27^. Regions containing at least one candidate AD variant were retained, and candidate variants mapping to the same TAD region were analyzed jointly. Because a TAD region may contain multiple proximal genes, each TAD region was therefore extended to encompass the *cis* windows of all candidate genes assigned to that region, with each window defined as ±1.5 Mb from the transcription start site^27^.

For each tested TAD region on chromosome *c*, we performed targeted QTL mapping of all candidate molecular phenotypes at the AD variants assigned to this region. Candidate proximal phenotypes comprised cell-type-solved expression traits for genes within the TAD region; candidate distal phenotypes comprised cell-type-resolved *TransMap* phenotypes constructed from genes outside chromosome *c*. Proteomic, glycoproteomic and metabolomic phenotypes were additionally evaluated as potential downstream *trans*-biomarkers. A molecular phenotype was retained for colocalization analysis if at least one candidate AD variant in this TAD region showed a marginal QTL association at p-value< 10^−4^ as suggested in ColocBoost^25^. We jointly analyzed all retained molecular phenotypes with an AD GWAS meta-analysis using the disease-prioritized mode of ColocBoost in each TAD region, with AD designed as the focal trait (**Data resources and preprocessing** below). This mode uses expedited single-effect coupling to prioritize colocalization signals involving AD while allowing multiple distinct signals within the region. Each colocalization event was summarized by a 95% CoS, where its corresponding trait configuration specified the molecular phenotypes sharing the AD-associated signal. We retained colocalization events linking AD to at least one molecular phenotype and containing at least one variant with marginal p-value < 10^−4^ for AD colocalized molecular traits in configuration^25^. We then classified retained colocalization events as *AD-cis*, *AD-trans* or *AD-cis-trans* according to whether the configuration contained proximal *cis*-eGenes, distal *trans* phenotypes or both. *AD-cis-trans* configurations were interpreted as AD-supported candidate *cis-trans* regulatory paths. For comparison, we additionally fine-mapped the AD GWAS using SuSiE and classified an AD locus as *AD-only* when its 95% CS shared no variants with any AD-centered 95% CoS containing at least one molecular effect.

### Empirical assessment of statistical calibration and reproducibility

To assess the calibration and robustness of *TransMap*, we performed two complementary empirical analyses: (i) 50:50 split-sample analyses to assess reproducibility of *trans*-MaxPIP annotation from genome-wide fine-mapping analysis, and (ii) genome-wide permutation analyses to evaluate the null calibration of fine-mapping and multi-phenotype colocalization for downstream *trans*-QTL discovery. These analyses evaluated two distinct properties of the framework: control of spurious discoveries under the null and generalizability of the inferred programs and genetic associations across donor subsets.

i. Split-sample assessment of *TransMap trans*-MaxPIP annotation reproducibility. We randomly divided donors into two non-overlapping halves (50:50 donor-level split) for each expression profile. We ran the full workflow independently in each split, including *trans* phenotype construction and genome-wide fine-mapping at three *trans*-regulatory scales. Thus, Hotspot modules, WPC co-regulatory activity phenotypes and sCCA-derived global embeddings were estimated independently in the two splits, without transferring program definitions or model parameters between two splits. We evaluated reproducibility using two complementary metrics. First, we calculate the Pearson correlation between split-specific *trans*-MaxPIP annotations, separately for three *trans*-regulatory scales. Second, we evaluated whether the independently derived annotations captured consistent disease-relevance genetic information. We applied the same S-LDSC models to each split-specific *trans*-MaxPIP annotation and compared heritability enrichment and marginal *τ*^∗^ estimates (**Methods** below).
ii. Permutation-based assessment of null calibration. We assessed the calibration of fine-mapping using SuSiE for individual *trans* genes and multi-phenotype colocalization using ColocBoost when jointly analyzing thousands of *trans*-eGene, *trans*-eProgram, and *trans*-biomarker phenotypes. We performed genome-wide permutations across 1,361 LD blocks used in the primary analyses. For statistical fine-mapping, we randomly shuffled donor labels in the genotype matrix while molecular phenotypes was unchanged; then we applied SuSiE to each permuted block and defined the genome-wide empirical false discovery rate (FDR) as the number of fine-mapped *trans*-eQTL under permutation divided by the number of *trans*-eQTL detected in the original data. For multi-phenotype colocalization, two permutation schemes separated the effect of analyzing thousands of phenotypes from that of their observed correlation structure. First, to assess null colocalization events when cross-phenotype correlations were disrupted, we independently permuted donor labels for each *trans*-phenotype within each block while leaving the genotype matrix unchanged. This preserved LD among variants but disrupted genotype-phenotype associations and the observed correlations among *trans*-phenotypes. Second, to assess null colocalization events under the phenotype covariance present in the primary analysis, we applied a single donor-label permutation to the genotype matrix within each block while leaving all *trans*-phenotypes unchanged. This procedure preserved both LD among variants and covariance among *trans*-phenotypes while disrupting genotype-phenotype associations. We reran ColocBoost under each permutation scheme and estimated a genome-wide empirical FDR as the number of colocalization events detected under permutation divided by the number of events detected in the original data. To account for differences in the number of phenotypes tested across LD blocks, we also calculated a weighted FDR. Each detected event was weighted by the fraction of phenotype in its trait configuration among all phenotypes tested in that block, with identical weights applied to permuted and observed events. We then matched this permuted *trans*-only colocalization events to genome-wide *cis*-gene events identified from the unpermuted data. A false *cis-trans* match required at least one shared variant between their 95% CoS. The genome-wide empirical *cis-trans* FDR was calculated as the number of *cis-trans* matches obtained under permutation divided by the number of *cis-trans* matches obtained in the original analysis.

### Multi-omics and GWAS data sources and preprocessing

The primary empirical analyses were applied to matched whole-genome sequencing (WGS) and molecular phenotype resources from ROSMAP donors^80,81^ (average *N* = 481, **Supplementary Table 1**). Genotype data were derived from quality-controlled WGS dosages, and molecular phenotypes were measured in the same donor cohort and matched at the donor level. The primary molecular resources included bulk and single-nucleus RNA-sequencing profiles across 6 cell types and 3 brain regions, and bulk proteomic, glycoproteomic and metabolomic measurements from the aging brain cortex. Bulk RNA-seq data were generated from postmortem tissues including dorsolateral prefrontal cortex (DLPFC), posterior cingulate cortex (PCC) and anterior cingulate cortex (AC). Standard QC and adapter trimming was performed on raw reads, followed by alignment via STAR and gene expression quantification via RNASeqQC^82,83^. Genes were removed if over 20% samples have TPM expression level of 10% or less. Single-nucleus RNA-seq data were aggregated into donor-level pseudobulk expression profiles, quantified as counts per million (CPM), for major brain cell classes, including excitatory neurons (Exc), inhibitory neurons (Inh), microglia (Mic), astrocytes (Ast), oligodendrocytes (Oli) and oligodendrocyte precursor cells (OPC). For downstream biomarkers, Selected Reaction Monitoring (SRM) proteomics was performed on DLPFC samples, and the peptide relative abundances were log2 transformed after quality control and filtering for proteins quantified in at lease 50% of samples. Glycoproteome was also perform on DLPFC samples using an improved glycopeptide-enrichment workflow followed by liquid chromatography-tandem mass spectrometry^84^. The 9,044 detected glycopetiforms originated from 1,217 distinct glycoproteins, which are log-transformed after excluding glycopeptiforms with fewer than two-thirds samples with valid measurements. Missing values for protein and glycoprotein were imputed using the grouped Empirical Bayesian Matrix Factorization method^85^. The 656 brain metabolites were measured in DLPFC samples and profiled using untargeted mass spectrometry, where metabolite abundances were log-transformed^86^. For all *trans*-acting molecular phenotypes, we removed the technical factors (batch, RNA integrity number, post-mortem interval), biological covariates (sex, age at death), and the top 15 genotype PC^26^. Details on the consideration of additional hidden factor derived from phenotype matrix were described in **Supplementary Note S1**. We restricted the QTL analysis to autosomal variants with minor allele frequency (MAF) > 0.05 and minor allele count (MAC) > 5.

For AD-centered *TransMap* analysis, we constructed a custom LD reference panel from the Alzheimer’s Disease Sequencing Project (ADSP), consisting of approximately 17,000 whole-genome sequenced (WGS) individuals of European ancestry. We primarily used a recent AD GWAS meta-analysis summary statistics (cases=128,681, controls=849,833)^54^ after performing quality control against this reference panel, including allele harmonization and LD mismatch detection using SLALOM, with flagged suspicious variants removed^87^. As a secondary AD GWAS, we considered meta-analysis summary statistics with a smaller effective sample size (cases=111,326, controls=677,663)^56^ used in our previous analysis^25^, followed by the same allele harmonization and quality control steps above.

### Benchmarks of local program *trans*-regulatory analysis

For *TransMap-local*, we benchmarked two complementary components: the regulatory coherence of local co-expression modules and its utility for *trans*-QTL mapping. First, using independent empirical *trans-*eQTL datasets, we tested whether the co-expression modules defining local programs grouped genes with coordinated *trans*-regulatory effects. We used empirical evidence because simulations do not readily capture the complex covariance structure of gene expression in human tissues. Second, we used simulations to assess whether WPC-based program phenotypes improved *trans*-QTL detection for a specified local program.

We first benchmarked Hotspot against WGCNA, a widely used method for detecting co-expression modules, using external and internal *trans*-eQTL resources. Hotspot and WGCNA modules were constructed from the same donor-level expression resource, with method-specific parameters described in **Methods**. Variant-program pairs were considered active when the program-level association passed the Bonferroni-corrected threshold of 10^−5^. For an active pair between variant *j* and module *G_m_*, *trans*-eQTL coherence was defined as

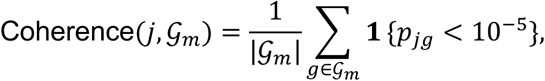

where, 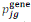 denotes the individual-gene *trans*-eQTL p-value for variant *j* and gene *g* in module G*_m_*. Larger coherence indicates that a program-level association is supported by multiple genes in the module rather than by a single outlier gene. We implemented a match internal benchmark evaluated ROSMAP cell-type-resolved co-expression modules using matched ROSMAP *trans*-eQTL association, In addition to ROSMAP benchmark, we also used two external resources to benchmark program-level coherence for co-expression module detection methods. For blood-based benchmarking, GTEx whole-blood expression profiles (*N* = 673)^7^ were paired with eQTLGen *trans*-eQTL associations for 10,317 tested trait-associated variants^4^, with 3,961 independent variants after LD pruning (*r*^2^ < 0.1) used in the program-level coherence evaluation. For brain-based benchmarking, GTEx brain cortex expression profiles (*N* = 208)^7^ were paired with MetaBrain *trans*-eQTL associations for 228,819 tested variants^10^, with 25,035 independent variants after LD pruning (*r*^2^ < 0.1) used for the same evaluation.

We next evaluated program-level *trans*-QTL testing for a specified gene program containing *M* genes using simulation studies. We first considered a single variant affecting the gene program through a *cis*-regulated gene. Let ***G*** ∈ ℝ*^n^*^×1^ denote the genotype vector for the tested variant, sampled from Binomial(2, *f*), where *f* was drawn from the empirical MAF distribution in ROSMAP donors (**Supplementary Figure S2a**). Let *β*_cis_ denote the effect size from the variant to the *cis*-gene, and let 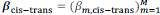 denote the vector of effects from the *cis*-gene to the *M trans*-genes. We generated expression for the *cis*-gene, ***Y***_cis_, and each *trans*-gene, ***Y****_m_*_,trans_, as

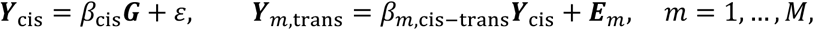

where *ε* ∼ N(0, *I_n_*) denotes residual noise for the *cis*-gene. The residual expression matrix ***E*** = [***E***_1_, …, ***E****_M_*] was generated with row-wise covariance **Σ***_E_*, using an AR(1) structure 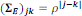. We assigned *β*_cis_ such that the per-SNP *cis* heritability of 0.04 and varied *β_m_*_,cis−trans_ to control the per-SNP *cis* to *trans* heritability^25,39^, defined as per-SNP *trans* 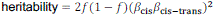.

The primary numerical study considered six broad settings: (i) program size (*M* = 20,50,100), matched to the median size of modules in real data analyses; (ii) sample size, varied from 400 to 1,000; (iii) the proportion of causal *trans*-genes affected by the variant, varied from 0% to 100%, with 0% used to evaluate type-I error (T1E); (iv) residual expression correlation structure **Σ***_E_*, with *ρ* = 0.1,0.3,0.5; (v) per-SNP *trans* heritability values corresponding to low-(0.0025), intermediate-(0.004) and high-power (0.01) regimes^39^; and (vi) sign heterogeneity of *trans*-QTL effects, where homogeneous effects assign all causal *trans*-effects positive signs and heterogeneous effects assign balanced positive and negative signs. In each replicate, we randomly sampled 1-3 independent causal variants with probabilities of 85.9%, 10.6% and 3.6%, respectively, based on their observed frequencies on ROSMAP (**Figure 2b**). When multiple variants were simulated, each variant affected a disjoint subset of genes within the same module. *TransMap-local*, with Bonferroni correction across retained WPCs (*λ* > 1), was compared with standard single-gene *trans*-eQTL testing using Bonferroni correction across genes in the module, Eigengene testing using only the first expression PC in a program^40^, and TransPCO, an omnibus test that combines multiple PC-based association tests^16^. Details of each competing method are described in **Methods**. Performance was evaluated at the level of *trans*-QTL discovery using a significance threshold of p-value < 10^−5^. We used two principal metrics: statistical power and T1E. Power was defined as the proportion of true module-level *trans*-QTL correctly identified across replicates, and T1E was defined as the proportion of null replicates in which a *trans*-QTL was detected. For each simulation setting, 1,000 replicates were generated to evaluate statistical power and 10^8^ replicates were generated to evaluate T1E at nominal p-value of 10^−5^.

### Benchmarks of global program *trans*-regulatory analysis

For *TransMap-global*, we benchmarked whether sparse association-driven embeddings recovered the underlying variant-gene *trans*-effect structure. Unlike the local benchmark, which evaluates *trans*-QTL detection for a specified gene module, the global benchmark evaluates recovery of low-dimensional regulatory structure at the variant-gene pair level. We used a structured causal-network simulation following the benchmark developed for ARCHIE^17^, in which groups of variants act through upstream regulated genes and induce coordinated but indirect distal expression responses. The simulation contains 40 *trans*-active causal variants and nine distal genes, with 20 additional non-*trans* variants included to evaluate false discovery rate (FDR). In this structured network, *trans*-active variants are organized into five-SNP blocks, and each block affects the expression of one proximal *cis*-gene mediator. The proximal mediators then propagate effects through three downstream *trans*-gene layers (**Figure 1f**). Same as in local-program simulation, genotype vectors were simulated independently for 60 variants. For each variant *j*, ***G***_j_ ∼ Binomial(2, *f*_j_), where *f*_j_ was drawn from the empirical MAF distribution in ROSMAP donors (**Supplementary Figure S2a**). Let *β*_0_ denote the effect size from a variant to its proximal *cis*-gene mediator, and let ***Y***^0^ denote expression of the proximal *cis*-gene mediators. Let *β_P_* denote the effect size in layer *P*, for *P* = 1,2,3, where *P* = 1 represents the propagation from *cis*-gene mediators to the first layer of *trans*-genes. Let ***Y****^P^* denote expression of *trans*-genes in layer *P*. The layered graph can be summarized schematically as

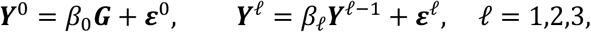

where ***ε****^P^* ∼ N(0, *I_n_*) denotes residual noise for gene expression. We assigned *β*_0_ such that the per-SNP *cis* heritability of 0.04 and varied *β_P_* to control the path-level *trans* heritability for different causal layers, defined as 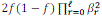 for a representative path at layer *P* = 1,2,3.

The primary numerical study considered four broad settings: (i) *trans* heritability, varied by changing the downstream layer-specific effect sizes to span low-(0.0025), intermediate-(0.004) and high-power (0.01) regimes for the weakest layer, and (ii) embedding sparsity, varied by changing the target proportion of non-zero loadings under sparse (0.4), dense (0.8) and cross-validation-optimized settings. We additionally generated an independent held-out sample of 1,000 individuals for methods requiring an expression covariance estimate. *TransMap-global* was compared with ARCHIE, with variant-gene association *Z*-scores were calculated from the simulated genotypes and distal gene expression. Details of the competing-method implementation are provided in **Methods**. Performance was evaluated at the level of variant-gene *trans*-effect recovery from paired variant and gene embeddings using precision-recall curves (PRC). Let 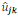 and 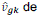 denote the variant loading and gene loading for component *k*. Pair-level evidence was summarized as 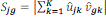, where larger *S_jg_* indicates stronger alignment between variant *j* and gene *g* across the first *K* paired components, providing stronger evidence that the pair belongs to a shared *trans*-effect structure. PRC was obtained by ranking variant-gene pairs according to *S*_jg_, with true positives defined as simulated *trans* variant-gene pairs connected by a non-zero directed path from a variant to a distal gene. Precision was defined as the proportion of selected pairs in true *trans* variant-gene pairs, recall was defined as the proportion of true pairs recovered.

### Benchmarking *TransMap* against existing *trans*-QTL and *trans*-program methods

In simulations, we compared *TransMap-local* with conventional single-gene *trans*-eQTL testing (*trans*-eGene) and two local program-level *trans*-QTL calling methods, Eigengene^40^ and TransPCO^16^. For a module containing *M* genes, *trans*-eGene tested each gene separately and assigned the module a Bonferroni-corrected p-value. For Eigengene, we computed the first principal component (PC) of the donor-level expression matrix using ‘prcomp’ in R and tested its association with each variant by linear regression. We ran TransPCO using the code provided in https://github.com/liliw-w/Trans which retained components with eigenvalues *λ* > 0.1 and combined the six PC-based association test p-values by ACAT^88,89^. For global *trans*-program discovery, we compared *TransMap-global* with ARCHIE^17^. The cross-covariance matrix is defined by 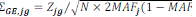, where *Z*_jg_ is z-score between variant *j* and gene *g* from marginal association, *N* is sample size and *MAF*_j_ is minor allele frequency of variant *j*; then we fitted the sparse components with the same PMD implementation used for *TransMap-global*.

For the real data heritability comparisons, we converted competing methods into donor-level phenotypes wherever the original method did not provide phenotypes suitable for statistical fine-mapping analysis. Eigengene was fine-mapped using the same chromosome-exclusion strategy to construct phenotype from the first PC of the donor-level expression matrix. Because the native TransPCO test produces a variant-level combined p-value rather than a program activity phenotype, we defined TransPCO* by retaining expression PCs with eigenvalues *λ* > 0.1 and fine-mapping each PC separately; TransPCO* is therefore an adaptation for fine-mapping, not the original ACAT test by combining p-values from six PC-based association tests. We additionally used AUCell^44^ to calculate a continuous, ranked-based activity score (aucMaxRank=0.2) for each donor and local program as the phenotype, then fine-mapped those phenotypes. Details of co-expression program detection are provided in **Supplementary Note S4.1** for Hotspot and **Supplementary Note S5** for WGCNA. For the global comparison, we projected donor expression onto ARCHIE-derived global gene embeddings using the same leave-one-chromosome-out phenotype construction applied to *TransMap-gobal*. After performed genome-wide fine-mapping, each method was summarized as variant-level *trans*-MaxPIP annotations and evaluated using the same S-LDSC analysis.

### MaxPIP-based *trans*-acting annotation and heritability analysis

To evaluate whether *trans*-QTL implicated by different scales of *TransMap* inform disease risk, we constructed variant-level *trans*-acting annotation based on the *maximum posterior inclusion probability* (*trans*-MaxPIP) from genome-wide *trans*-QTL fine-mapping. For each *trans*-acting phenotype *y*, such as individual *trans*-eGenes, local WPC-based program scores or global sCCA-based program scores, let 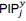 denote the PIP from fine-mapping of variant *j* to *trans*-phenotype *y*. Let ℋ denote the set of *trans*-phenotypes whose 95% CS contains variant *j*, we then defined

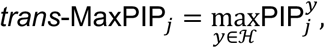

with variants do not present in any credible set assigned a score of zero. Separate *trans*-MaxPIP annotations were generated for *TransMap-eGene*, *TransMap-local* and *TransMap-global*, and analogous annotations were generated for competing methods where appropriate. Except for the cell-type-resolved variant annotation from each scale, we also generated cross-cell-types annotations by taking maximum value of *trans*-MaxPIP_j_ for variant *j*. We then applied S-LDSC to quantify whether *trans*-acting annotations captured complex disease heritability across 57 GWAS summary statistics datasets^25^, conditioning on 53 baseline annotations comprising coding, conserved and tissue-aggregated epigenomic annotations^25,34^ (**Supplementary Table 1**). We considered two metrics, enrichment and standardized effect size, across all 57 traits, as wells as separately across the blood and brain-related traits. Heritability enrichment quantified the concentration of trait heritability in annotated variants, whereas the standardized effect size *τ*\* estimated the conditional informativeness of an annotation after accounting for baseline annotations and other annotations in the same model. Joint S-LDSC models were used to test whether single-gene, local-program and global-program *trans*-QTL annotations carried nonredundant heritability information. Both enrichment and *τ*\* were estimated via a genomic block-jackknife procedure with 200 blocks that also provides standard errors. More detailed decription of metrics can be found in **Supplementary Note S6**.

### xQTL-informed independent and background variants selection

We constructed an xQTL-informed set of independent variants to represent common variants with prior evidence for molecular regulatory activity. These variants were used for *TransMap-global* program learning and for benchmarking against competing methods, including evaluation of *trans*-eQTL coherence and *trans*-eQTL calling. Candidate variants were assembled from multiple resources, including eQTLGen *cis*– and *trans*-eQTL^4^, MetaBrain *cis*– and *trans*-eQTL^10^, cerebrospinal fluid (CSF) protein QTL^90,91^, and FunGen-xQTL *cis*-QTL across four cohorts and 77 molecular datasets^26^. Candidate variants were harmonized to ROSMAP whole-genome sequencing genotypes. We then removed duplicate variants, retained common variants with MAF ≥ 0.05, and performed LD pruning using the ROSMAP genotype reference with a pairwise *r*^2^ < 0.1 threshold. This procedure yielded 43,090 approximately independent xQTL-informed variants (**Supplementary Figure S1b**. Resource-specific extraction rules and pipeline implementation details are provided in **Supplementary Note S3**.

For WPC background construction in *TransMap-local*, null variants were selected from genome-wide common variants, after excluding the xQTL-informed variants described above and any variants in LD with these variants (*r*^2^ > 0.1). The remaining variants were then LD-pruned to obtain 26,363 approximately independent null variants. This design reduces enrichment of known *cis*-, *trans*-QTL and other xQTL signals in the empirical background used by WPC, while limiting redundancy among nearby correlated variants. The resulting null set provides a regulatory-signal-depleted common variant background for estimating residual co-expression structure and calibrating WPC-based program activity comparisons.

### Quantifying downstream biomarker-QTL convergence of *trans*-regulatory signals

To evaluate whether *trans*-eGenes or *trans*-ePrograms converge with downstream biomarkers through the shared genetic signals, we defined the biomarker convergence *Enrichment* and *Recall* at the colocalization event level, where each event is represented by a 95% CoS and its corresponding trait configuration. Let C_0_ denote all retained 95% colocalization events considered in the analysis, C*_A_* denote colocalization events for *trans*-eGenes or *trans*-ePrograms, and C*_B_* denote colocalization events for downstream biomarkers. Enrichment quantifies whether colocalization events involving *trans*-eGenes or *trans*-ePrograms are more likely to also involve downstream biomarker-QTL signals than expected from all colocalization events:

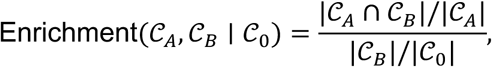

where Enrichment > 1 indicates that downstream biomarker-QTL events occur more frequently within the specified *trans*-regulatory event set than expected from all retained colocalization events. Recall quantifies the fraction of downstream biomarker-QTL colocalization events that are also captured by the given *trans*-regulatory signal set:

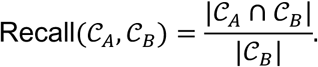

For biomarker-category analyses, C*_A_* was defined separately for *trans*-eGene and *trans*-eProgram, whereas C*_B_* was restricted to events involving protein-, glycoprotein– or metabolite-QTL signals. For cell-type analyses, C*_A_* and C_0_ were restricted to events containing the given cell type. Statistically significant and 95% confidence intervals for *Enrichment* and *Recall* were estimated using a leave-one-chromosome-out jackknife.

### Functional characterization of QTL variants and target genes

To characterize the genomic context and biological interpretation of *cis* and *trans* genetic signals identified by *TransMap* colocalization analysis, we classified colocalization events into three regulatory categories according to their trait configurations. A ‘*cis*-only’ colocalization event contained one or more proximal *cis*-eGenes without a distal *trans*-eGene or *trans*-eProgram. A ‘*cis–trans*’ colocalization event contained at least one proximal *cis*-eGene and at least one distal *trans*-eGene or *trans*-eProgram. A ‘*trans*-only’ colocalization event contained at least one distal *trans*-eGene or *trans*-eProgram without a proximal *cis*-eGene. We then tested whether these categories showed distinct genomic and functional architectures by characterizing variant-level conservation^48^, gene-level loss-of-function (LoF) constraint^92^, human transcription factor (TF)^93^, and protein–protein interaction (PPI)^46^. We used the same *Enrichment* across variant-, gene– and gene-gene interaction-level analyses after redefining the analysis unit and background universe. Depending on the analysis, a unit represented a colocalized variant in 95% CoS, the gene nearest to a QTL variant, a molecular target gene, or a gene-gene interaction. Let U denote the background set of units, and let A, ℬ ⊆ U denote two sets of interest. Enrichment is defined as Enrichment(A, ℬ ∣ U) = (|A ∩ ℬ|/|A|)/(|ℬ|/|U|).

i. For variant-level characterization, distance to transcription start site (TSS) was calculated from the corresponding each CoS to the nearest protein-coding gene TSS. Variant conservation was evaluated using Zoonomia primate PhastCons scores^48^, with conserved variants defined as PhastCons > 0.9.
ii. For gene-level characterization, we distinguished two target-gene definitions. First, nearest genes were defined as protein-coding genes closest to representative CoS and were used to characterize locus-proximal gene properties. Second, molecular target genes were defined from the phenotypes included in the trait configuration, including proximal *cis*-eGenes, distal *trans*-eGenes and genes contributing to *trans*-ePrograms. Loss-of-function-constrained genes were defined as genes with pLI > 0.9. Human transcription factors were defined using a curated 1,631 human transcription-factor catalogue^46^.
iii. For gene-gene interaction characterization, the unit of analysis was a gene connection. For *cis–trans* configurations, observed gene pairs were defined between proximal *cis*-eGenes and distal *trans*-eGenes or genes assigned to *trans*-ePrograms in the same colocalization event. For within-*trans* analyses, observed gene pairs were defined among distal genes linked to the same *trans*-regulatory event or program. Protein–protein interaction support was evaluated using STRINGdb^45^ after mapping genes to protein-coding identifiers.

For AD-centered colocalization, we compared lead-variant conservation across *AD-only*, *AD*-*cis*, *AD*-*trans* and *AD-cis-trans* events using two-sided Wilcoxon rank-sum tests^94^, where the lead variant was the highest-VCP variant in each 95% CoS (or the highest-PIP variant from AD fine-mapping for *AD-only* loci). To test whether conservation depended on cellular context, we fitted a linear model regressing the Zoonomia mammalian phyloP score of the lead variant on a binary indicator denoting the presence of a given cell type among the initiating (*cis*) cell types, the recipient (*trans*) cell types, or a specific initiating–recipient cell-type pair, adjusting for the lead-variant VCP and the number of distinct cell types in the colocalization event.

### Cross-cell-type enrichment of neuron-initiated *cis–trans* regulatory configurations

We evaluated whether candidate *cis–trans* regulatory configurations preferentially connected specific initiating and recipient cell types. Each candidate path was represented by a proximal *cis*-eGene, its initiating cell type, a distal *trans*-eGene or *trans*-eProgram, and the recipient cell type in which the distal signal was observed. Cell-type-pair enrichment was calculated for each initiating-recipient cell-type pair by comparing the observed pairs to a randomized background from the null distribution. To generate the null distribution, we randomly permuted the cell-type labels separately among *cis*-signals and among *trans*-signals, while preserving the colocalization event, the number of *cis* and *trans* signals within each event, and the marginal cell-type composition of each signal type. This preserves the overall colocalization-level signal structure but disrupts the observed pairing between initiating and recipient cell types. Statistical significance was calculated as the fraction of shuffled datasets with enrichment at least as large as the observed enrichment.

### Co-culture validation of neuron–astrocyte regulatory configurations

To obtain orthogonal support for neuron–astrocyte-specific *cis*–*trans* regulatory configurations, we compared the genes implicated by *TransMap* candidate paths with a published neuron–astrocyte co-culture transcriptomic dataset^51^, in which human iPSC-derived *Ngn2*-induced glutamatergic neurons were cultured either alone or with primary mouse cortical astrocytes. This study defined 5,330 astrocyte-responsive neuronal genes as human genes differentially expressed between co-cultured and mono-cultured neurons, and 3,587 neuron-responsive astrocytic genes as mouse genes differentially expressed between co-cultured and mono-cultured astrocytes at an FDR level of 0.01. For validation of *TransMap* candidate regulatory configurations, recipient-astrocytic *trans*-eGenes or genes assigned to recipient-astrocytic *trans*-ePrograms from neuron-initiated configurations were compared with neuron-responsive astrocytic genes, whereas recipient-neuronal *trans*-eGenes or genes assigned to recipient-neuronal *trans*-ePrograms from astrocyte-initiated configurations were compared with astrocyte-responsive neuronal genes. *Enrichment* quantified whether *TransMap* recipient *trans*-eGenes or ePrograms were more concentrated in the corresponding co-culture-responsive gene set than expected for protein coding genes. Statistical significance for enrichment was evaluated using a leave-one-chromosome-out jackknife.

### Spatial proximity validation of initiated-neuronal-recipient-glial regulatory configurations

To evaluate whether *cis*-neuronal and *trans*-glial genes from candidate *cis*–*trans* configurations showed compatible spatial proximity in human AD brain, we analyzed a published ROSMAP 10x Visium spatial transcriptomic dataset from DLPFC^52^. The dataset contains 32 spatial transcriptomic sections, including 4 control samples from 2 healthy donors and 28 samples from 15 donors with AD pathology. Spots were assigned to *cortical layers* L1–L6 or *white matter* using the layer annotations provided by the original study^95^. We grouped L1–L3 as *outer cortical layer*, L4–L6 as *deep cortical layer* and retained white matter as a separate anatomical compartment. Gene-level spatial differential expression (DE) analysis was performed using the zero-inflated hurdle model implemented as zlm function^96^. For initiated-neuronal-recipient-glial stratum, we compared spatial expression of initiated-neuronal genes in deep cortical layer against outer cortical layer, and compared expression of recipient-glial genes in white matter against all cortical-layer spots. Following the original study, AD samples were stratified into lower, middle and higher pathology groups, with 10, 9 and 9 samples, respectively. DE analysis was evaluated using the full dataset and within each control or AD-pathology group.

### Donor-stratified subsampling of excitatory neurons

To assess whether the greater representation of excitatory neurons influenced the inferred cross-cell-type regulatory architecture, we applied donor-stratified subsampling to generate excitatory neuron subsets containing approximately *M* = 100,000, *M* = 200,000and *M* = 300,000 nuclei. The complete excitatory neuron comprised 630,488 nuclei from 419 donors, accounting for 39.8% of all profiled nuclei. For each target size *M*, nuclei were randomly sampled within each donor using a common sampling probability of *M*/630,448, subject to retaining at least 10 nuclei per donor. This procedure maintained representation of all donors while reducing cell numbers proportionally across donors. For each subsample, we reconstructed donor-level excitatory pseudobulk expression profiles and repeated expression preprocessing, covariate adjustment, *cis*-eQTL and individual gene *trans*-eQTL fine-mapping analysis across 1,361 LD blocks, and the corresponding *cis-trans* colocalization analysis using the parameters applied in the primary analysis. Data for all other cell types were held fixed, thereby isolating the effect of excitatory neuron sampling depth.

### Genome-wide fine-mapping of cell-type composition QTL

To characterize genetic effects on brain cellular composition, we generated donor-level phenotypes for cell-type proportions in ROSMAP. For each donor, cell-type proportions were calculated as the fraction of annotated nuclei assigned to each of six major cell types. Each proportion phenotype was adjusted for age, sex, postmortem interval and the first 15 genotype PCs. For each phenotype, we performed genome-wide QTL fine-mapping across 1,361 LD blocks used in the primary *TransMap* analyses. Given the generally modest genetic effects on cell-type composition, candidate QTL were represented by 50% CS for downstream integration. All association and fine-mapping settings matched those used in corresponding primary analyses.

### Mediation analysis of candidate *cis-trans* regulatory configurations

To assess candidate molecular mediation at colocalized loci, we fitted a structural equation model (SEM) for each variant–mediator–outcome triplet, specifying genotype dosage at the lead variant with highest VCP from the corresponding 95% colocalization confidence set as the exposure, proximal *cis*-eGene expression as the mediator, and a distal *trans*-eGene or *trans*-eProgram phenotype as the outcome. Molecular phenotypes were adjusted for age, sex, pmi and the top 15 genotype PC before model fitting. We estimated the indirect association as the product of the genotype–mediator and mediator–outcome coefficients. For assays with partially overlapping donor samples, SEM was fitted with ‘lavaan’ using full-information maximum likelihood under the missing-at-random assumption^53^. We assessed uncertainty in the indirect association using 1,000 donor-level bootstrap resamples and percentile 95% confidence intervals. Selected triplets were further examined using Bayesian SEM and missing-data sensitivity analyses.

### Software and reporting

The *TransMap* workflow is implemented as a modular command-line pipeline with components for phenotype construction, fine-mapping, annotation construction and colocalization. Analyses used custom code together with established tools for sparse CCA, fine-mapping, S-LDSC and multi-phenotype colocalization. Software versions, command-line parameters, random seeds and data-access restrictions are reported in the **Code Availability**, **Data Availability** and **Reporting Summary** materials.

## Data Availability

All data produced in the present work are contained in the manuscript.

## Acknowledgements

We thank members of the Wang and Dey labs for discussions and support. This study was coordinated through the Alzheimer’s Disease Sequencing Project Functional Genomics Consortium (FunGen-AD), with contributions from consortium principal investigators. We thank the National Institute on Aging Alzheimer’s Disease Data Storage Site (NIAGADS) at the University for data management and resource distribution. We acknowledge the Genome Center for Alzheimer’s Disease (GCAD) for processing and harmonizing ADSP whole-genome sequencing data used to construct the LD reference. We would like to acknowledge Harm-Jan Westra, Ellen A. Tsai and Lude Franke for sharing unpublished *trans*-eQTL results from the MetaBrain dataset (de Klein et al 2023). This work was supported in part by NIH grants R01AG076901 (G.W.), R01AG086467 (G.W.), R00HG012203 (K.K.D.), R01AG091534 (M.S., A.B., H.K.), and P30CA008748 (K.K.D.). This work was also supported in part by Alzheimer’s Association Grant ADSF-21-816675 (H.K., M.T.) and by a grant from The Urbut Family Foundation (G.W.). NIAGADS and GCAD were supported by U24AG041689 and U54AG052427, respectively.

## Author Contributions

G.W., K.K.D. jointly supervised research. X.C. conceived and designed the experiments, performed analyses, interpreted the results and implemented to *transmap* pipeline with documentation and example vignettes. Y.W. contributed to global program level simulation studies and xQTL-informed variants extraction. H.S., H.Z., H.L. contributed to spatial transcriptomics data analysis. R.F. and H.K. contributed to glycoprotein data analysis and interpretation. Y.L. contributed to mediation analysis and interpretation. B.K., M.T., H.K. contributed to spatial transcriptomics data generation. B.L., C.G., G.C. contributed to neuron-astrocyte co-culture experiments and data generation. H.W., E.T., L.F. contributed to MetaBrain data generation. M.S., A.B., H.K. contributed to glycoprotein data generation. P.L.D., D.B., G.W. contributed and supervised other molecular QTL data generation. X.C., G.W., K.K.D. wrote and revised the manuscript.

## Supplementary Figures Legend

**Supplementary Figure 1.** Supporting analyses for local program benchmarking. **a.** Number of gene modules identified by Hotspot and WGCNA across GTEx and ROSMAP expression contexts. Colors indicate different co-expression program detection methods. **b.** Selection of 43,090 xQTL-informed independent variants from FunGen-xQTL resources after duplicate removal, minor allele frequency filtering and LD pruning. **c.** Schematic overview for simulation studies. **d.** Power of *TransMap-local*, TransPCO, Eigengene and conventional single-gene *trans*-eQTL testing at intermediate SNR across varying proportions of causal *trans*-genes, sample sizes, module sizes, residual correlations and sign heterogeneity. Error bars indicate 95% confidence intervals. Colors indicate different *trans*-QTL calling methods. **e.** Type I error (T1E) estimated across 10^8^null simulation replicates with 0% causal *trans*-genes, using a nominal significant threshold of 10^−5^ for all methods. Error bars indicate 95% confidence intervals. Numerical results are reported in Supplementary Data.

**Supplementary Figure 2.** Supporting analyses for global program benchmarking. **a.** Minor allele frequency (MAF) distribution of variants from ROSMAP WGS data that used in the simulations. Dashed line indicates MAF=0.05. **b.** Area under the precision-recall curves (AUPRC) of recovering true variant-gene pairs from *TransMap-global* versus ARCHIE at per-SNP *trans* heritability levels and sparsity-parameter grid. Colors indicate per-SNP *trans* heritability and shapes indicate sparsity. **d.** AUPRC of recovering true variant-gene pairs from *TransMap-global* versus ARCHIE at per-SNP *trans* heritability levels and MAF intervals. Colors indicate per-SNP *trans* heritability and shapes indicate MAF intervals. In **c** and **d**, the dashed line denotes equal performance. Numerical results are reported in Supplementary Data.

**Supplementary Figure 3.** Supporting analyses for genome-wide *trans*-eQTL discovery and *TransMap-eGene* annotation. **a.** Number of *trans*-eQTL identified across ROSMAP expression contexts after adjustment for different numbers of phenotype principal components (PCs) along with biological and genotype PC covariates. Red dashed lines indicate the corresponding analysis without any covariate adjustment. See Supplementary Note for details of covariates consideration. **b.** Genome-wide empirical FDR estimates for SuSiE fine-mapping and ColocBoost multi-phenotype colocalization under phenotype-wise permutation, which disrupted covariance among *trans* phenotypes, and genotype-label permutation, which preserved the observed phenotype covariance while disrupting genotype–phenotype associations. Results are shown for the overall empirical FDR, the empirical FDR weighted by the number of phenotypes tested and the empirical *cis–trans* FDR obtained by matching permuted distal events to observed *cis*-eQTL colocalization events. Numerical results are reported in Supplementary Data.

**Supplementary Figure 4.** Disease-heritability analyses of local and global *trans*-regulatory program annotations. **a.** Heritability enrichment and conditional standardized effect size (joint *τ*^∗^) of *TransMap-local*, TransPCO, Eigengene and AUCell *trans-*MaxPIP annotations derived from Hotspot or WGCNA modules. **b.** Heritability enrichment, marginal *τ*^∗^ and joint *τ*^∗^ of *TransMap-local*, native TransPCO and TransPCO* annotations, restricted to the 43,090 xQTL-informed variants. TransPCO* denotes annotation obtained by genome-wide fine-mapping of TransPCO-selected principal components (Methods). **c.** Marginal *τ*^∗^, heritability enrichment and joint *τ*^∗^ of *TransMap-global* and ARCHIE annotations. **d.** Heritability enrichment and marginal standardized effect size (*τ*^∗^) of *TransMap-eGene* annotations derived from genome-wide fine-mapping, hit-based fine-mapping or marginal association testing (left) and number of *trans*-eQTL retained by the genome-wide *TransMap-eGene* fine-mapping compared with *trans*-eQTL selected by hit-based fine-mapping or marginal association testing across expression contexts (right). Dashed line denotes equality. **e.** Heritability explained by fine-mapped *cis*-MaxPIP versus the union of *trans*-MaxPIP combined from *TransMap-eGene* and *TransMap-local* across 28 brain-related traits, dashed line denotes equality. In **a–d**, results are meta-analyzed across all 57 traits, 18 brain-related traits and 22 blood-related traits. S-LDSC models were conditioned on 53 baseline-LD annotations, with joint models additionally including all annotations shown in the corresponding panel. Error bars indicate 95% confidence intervals, and asterisks indicate statistical significance at FDR<5%. Numerical results are reported in Supplementary Data.

**Supplementary Figure 5.** Split-sample reproducibility of *TransMap* annotations and empirical null calibration of multi-trait colocalization. **a.** Pearson correlations between variant-level *trans*-MaxPIP annotations independently derived from two non-overlapping 50:50 donor splits for the *TransMap-eGene*, *TransMap-local* and *TransMap-global* scales. **b.** Heritability enrichment (top) and marginal standardized effect size (*τ*^∗^; bottom) of split-specific *trans*-MaxPIP annotations. Heritability enrichment, marginal standardized effect size (*τ*^∗^) and conditional *τ*^∗^ from joint S-LDSC models for *trans-*MaxPIP annotations derived from **c.** 50 MSigDB Hallmark gene sets and **d.** cell-type-resolved, data-driven SCENIC regulons using *TransMap*, TransPCO*, AUCell or Eigengene representations. Corresponding analyses. **e.** Heritability enrichment, marginal *τ*^∗^ and joint *τ*^∗^ of the Hallmark and SCENIC *TransMap* annotations modeled together with *TransMap-eGene*-MaxPIP and *TransMap-local*-MaxPIP annotations. S-LDSC results are meta-analyzed across all 57 traits, 18 brain-related traits and 22 blood-related traits, conditional on 53 annotations; joint models additionally included all annotations shown in the corresponding panel. Error bars indicate 95% confidence intervals, and asterisks indicate statistical significance. Numerical results are reported in Supplementary Data.

**Supplementary Figure 6.** Genome-wide colocalization patterns and functional characterization of *TransMap trans*-regulatory loci. **a.** UpSet plot of genome-wide colocalization events involving *cis*-eQTL, *trans*-eGene, *trans*-eProgram, protein-QTL, glycoprotein-QTL and metabolite-QTL. **b.** Log2 odds ratios of *trans*-eQTL from fine-mapping and colocalization analyses among external PsychAD and MetaBrain *trans*-eQTL associations with 95% confidence intervals. **c.** Proportion of *cis*-eQTL supported by different number of downstream *trans*-regulatory layers. **d.** Sankey plot linking trans-eQTL loci supported by *trans*-eGene or *trans*-eProgram to three modalities of biomarker-QTL. **e.** Enrichment and recall of cell-type-resolved *trans*-eQTL for overlap with protein, metabolite and glycoprotein QTL. Numerical results are reported in Supplementary Data.

**Supplementary Figure 7.** Genome-wide colocalization patterns and functional characterization of *TransMap trans*-regulatory loci. **a.** Enrichment for functional gene properties of a curated set of 1,639 human transcription factors compared with: genes nearest *cis-only*, *cis-trans*, *trans-only* loci in term of transcription start site (TSS) and *cis-*eGenes for *cis-only* and *cis-trans* loci. **b.** Distribution of distance to the TSS of nearest gene for *cis-only*, *trans-only* and *cis-trans* loci. **c-d.** Enrichment for evolutionary constraint of three classes’ **c.** variants for highly conserved sequence (PhastCons>0.9), and **h.** genes nearest to each loci (nearest TSS genes) and *cis*-eGenes at *cis-only* and *cis-trans* loci for loss-of-function constrained genes (pLI>0.9). In **a,c,**d, results are shown separately for *trans*-eGene (top) and *trans*-eProgram (bottom). Asterisks indicate statistical significance at FDR<5%. Error bars indicate 95% confidence intervales calculated by leave-one-chromosome-out jackknife. Numerical results are reported in Supplementary Data.

**Supplementary Figure 8.** Cell-type architecture and robustness of neuron-initiated glial *trans*-regulatory paths. **a.** Percentage of colocalized loci involving each brain cell type through a *cis*-eQTL or *trans*-eQTL signal. **b.** Enrichment of glial recipient cell types among candidate regulatory paths initiated by excitatory-neuron (left) or inhibitory-neuron (right) *cis* signals. **c.** Number of candidate cross-cell-type *cis–trans* loci for each initiating and recipient cell-type pair. **d.** Corresponding locus counts shown separately for *trans*-eGene and *trans*-eProgram supported paths. **e.** Enrichment of initiating–recipient cell-type pairs relative to a null distribution preserving cell-type representation and locus-level phenotype multiplicity, shown separately for *trans*-eGenes and *trans*-ePrograms. **f.** Numbers of excitatory-neuron *cis*-eGene and *trans*-eGene credible sets identified using the complete dataset and subsets containing 100,000, 200,000 or 300,000 excitatory-neuron nuclei. **g.** Numbers of candidate paths connecting excitatory-neuron *cis* signals to glial *trans* signals (heatmaps) and numbers of *cis*-eGene and *trans*-eGene loci identified in each cell type (bar plots) after excitatory-neuron subsampling. **h.** Distribution of donor-level cell fractions for the six major brain cell types used in cell-proportion-QTL mapping. **i.** Proportion of candidate neuronal-*cis*–glial-*trans* colocalization events overlapping cell-proportion QTL in the complete dataset and excitatory-neuron subsamples. Error bars indicate 95% confidence intervals, and asterisks indicate statistical significance. Numerical results are reported in Supplementary Data.

**Supplementary Figure 9.** **Pathway enrichment and orthogonal validation of candidate neuron–glia regulatory paths**. **a**. Reactome pathway enrichment of glycoproteins colocalized with excitatory neuron and astrocyte imitating-recipient events with FDR < 1%. **b**. STRING protein-protein interaction network of astrocyte trans-eGenes and glycoproteins from excitatory neuron and astrocyte imitating-recipient events. **c**. STRING network among all 25 genes annotated to RHO-ROCK and SEMA4D-induced cell migration pathways. In **b** and **c**, only proteins with association score > 0.4 are retained. **d.** Spatial differential expression patterns of neuronal *cis*-regulated genes between deep (L4–L6) and outer (L1–L3) cortical layers (left) and of their associated glial *trans*-regulated genes in white matter and cortical layers (right), stratified by AD pathological stage. **e.** Spatial organization of genes implicated through *trans*-ePrograms. The left panel shows differential expression of neuronal *cis*-regulated genes between deep and outer cortical layers; the right panel compares spatial differential expression evidence for recipient glial genes in white matter and cortical layers. Error bars indicate 95% confidence intervals, and asterisks indicate statistical significance. CTRL, cognitively normal control.

**Supplementary Figure 10.** Supporting analyses at the *SEC61G* and *GDA* loci. **a.** Reactome pathway enrichment for the microglial *trans*-eProgram (module 100) colocalized at the *SEC61G* locus. **b.** Colocalization plots at the *GDA* locus for the excitatory neuron *GDA cis*-eQTL, astrocyte *trans*-eProgram QTL, and 11 *trans*-glycosylation traits representing nine glycoproteins. Orange points mark variants in the reported 95% colocalization confident set. **c.** Pathway enrichment for the nine glycoproteins shown in panel b. **d.** Neuron-responsive astrocyte genes identified in co-culture experiment. Red points denote genes in *GDA* colocalized astrocyte program genes that overlap the co-culture response, blue points denote other neuron-responsive astrocyte genes (FDR < 0.01). The axes show log₂ fold change and −log₁₀(adjusted *P*). **e.** STRING PPI network of colocalized *trans*-glycoproteins and member genes in astrocyte *trans*-eProgram (Program 80). Orange nodes denote glycoproteins, blue nodes denote program genes also overlapped with neuron-responsive astrocyte genes in co-culture experiment, and grey nodes denote other program genes. Numerical results are reported in Supplementary Data.

**Supplementary Figure 11.** Multilayer and cell-type-resolved architecture of AD-associated colocalization events. **a.** UpSet plot of 112 AD-centered colocalization events across AD GWAS fine-mapping, *cis*-eGene, *trans*-eGene, *trans*-eProgram, *trans*-biomarker, AD-pathology and cell-fraction QTL layers. **b.** Number of AD colocalized loci supported by *cis*-eQTL and *trans*-eQTL, colored coded by the number of snRNA-seq cell types implicated at each locus. **c.** Percentage of AD-colocalized loci involving each brain cell type through *cis*-eQTL or *trans*-eQTL evidence and **d.** separating *trans*-eGene and *trans*-eProgram evidence. **e.** Associations between Zoonomia phyloP conservation scores of lead colocalized variants and the presence of individual initiating cell types, recipient cell types or initiating–recipient cell-type pairs. Error bars indicate 95% confidence intervals. **f.** Manhattan plot of the AD GWAS meta-analysis with tracks indicating cell-type-resolved *cis*– and *trans*-eQTL evidence at each colocalized locus. Loci are labeled by the colocalized *cis*-eGene; asterisks indicate the nearest-gene label for *AD–trans-only* loci. Numerical results are reported in Supplementary Data.

**Supplementary Figure 12.** AD-centered *TransMap* colocalization identifies *cis*, *trans* and biomarker regulatory layers with AD GWAS loci shown in secondary AD GWAS meta-analysis. a. UpSet plot of 69 distinct colocalization events from AD-target analysis, showing colocalization patterns across different modalities. Venn diagram shows the number of colocalization events supported by *cis*-eQTL and *trans*-eQTL. b. Number of AD colocalized loci supported by *cis*-eQTL and *trans*-eQTL, colored coded by the number of snRNA-seq cell types implicated at each locus. c. Variants conservation based on Zoonomia phyloP scores of the lead variant at each AD locus (highest MaxVCP for AD colocalized loci and MaxPIP for only AD fine-mapped loci). Asterisks denote significant differences between group (* FDR<5%, ** FDR<1%). d. Manhattan plot of the AD GWAS meta-analysis with colocalized loci labeled by *cis*-eGene or nearest gene at *AD-trans* only colocalized loci, and tracks below making the molecular QTL classes colocalized at each locus. Numerical results are reported in Supplementary Data.

**Supplementary Figure 13.** Supporting analyses of AD colocalized *cis-trans* signals at the *KNOP1* and *MS4A*-family gene loci. **a.** Heatmap of indirect effect estimates within the chromosome 16 *KNOP1* colocalization set. Columns indicate proximal *cis*-eGenes by context and rows indicate distal *trans*-eGenes or *trans*-ePrograms by context. Color shows the signed indirect effects, asterisks mark nominal significant effects with p-value<0.05. **b.** Number of colocalized *trans* phenotypes by recipient context in the chromosome 11 *MS4A*-family gene loci colocalization. **c.** Pathway enrichment analysis for two colocalized inhibitory neuron *trans*-ePrograms with FDR<5%. **d-e.** STRING networks of inhibitory neuron *trans*-eProgram module 59 (**d**) and module 115 (**e**), respectively. In **d**-**e**, red edges denote physical proximity links, and gray edges denote functional associations only. Numerical results are reported in Supplementary Data.

**Supplementary Figure 14.** Supporting analyses of AD colocalized *cis-trans* signals at the 16p11.2 locus. **a.** STRING PPI network of proteins encoded by genes in the *INO80E cis* region and the distal target *ABCA1*. **b.** Indirect associations between genotype of colocalized loci and local *cis*-eGenes, with *INO80E* as the mediator. **c.** Indirect associations with local *cis*-eGene as mediator and *INO80E* as the outcome. In **b-c**, points show estimated indirect effects with 95% confidence intervals. Numerical results are reported in Supplementary Data.

