## Supplementary Figures for "A multi-scale cell-type resolved map of *trans*-regulatory architecture in the aging human brain"

**Supplementary Figure 1**. **Supporting analyses for local program benchmarking. a.** Number of gene modules identified by Hotspot and WGCNA across GTEx and ROSMAP expression contexts. Colors indicate different co-expression program detection methods. **b.** Selection of 43,090 xQTL-informed independent variants from FunGen-xQTL resources after duplicate removal, minor allele frequency filtering and LD pruning. **c.** Schematic overview for simulation studies. **d.** Power of *TransMap-local*, TransPCO, Eigengene and conventional single-gene *trans*-eQTL testing at intermediate SNR across varying proportions of causal *trans*-genes, sample sizes, module sizes, residual correlations and sign heterogeneity. Error bars indicate 95% confidence intervals. Colors indicate different *trans*-QTL calling methods. **e.** Type I error (T1E) estimated across $10^8$ null simulation replicates with 0% causal *trans*-genes, using a nominal significant threshold of $10^{-5}$ for all methods. Error bars indicate 95% confidence intervals. Numerical results are reported in Supplementary Data.

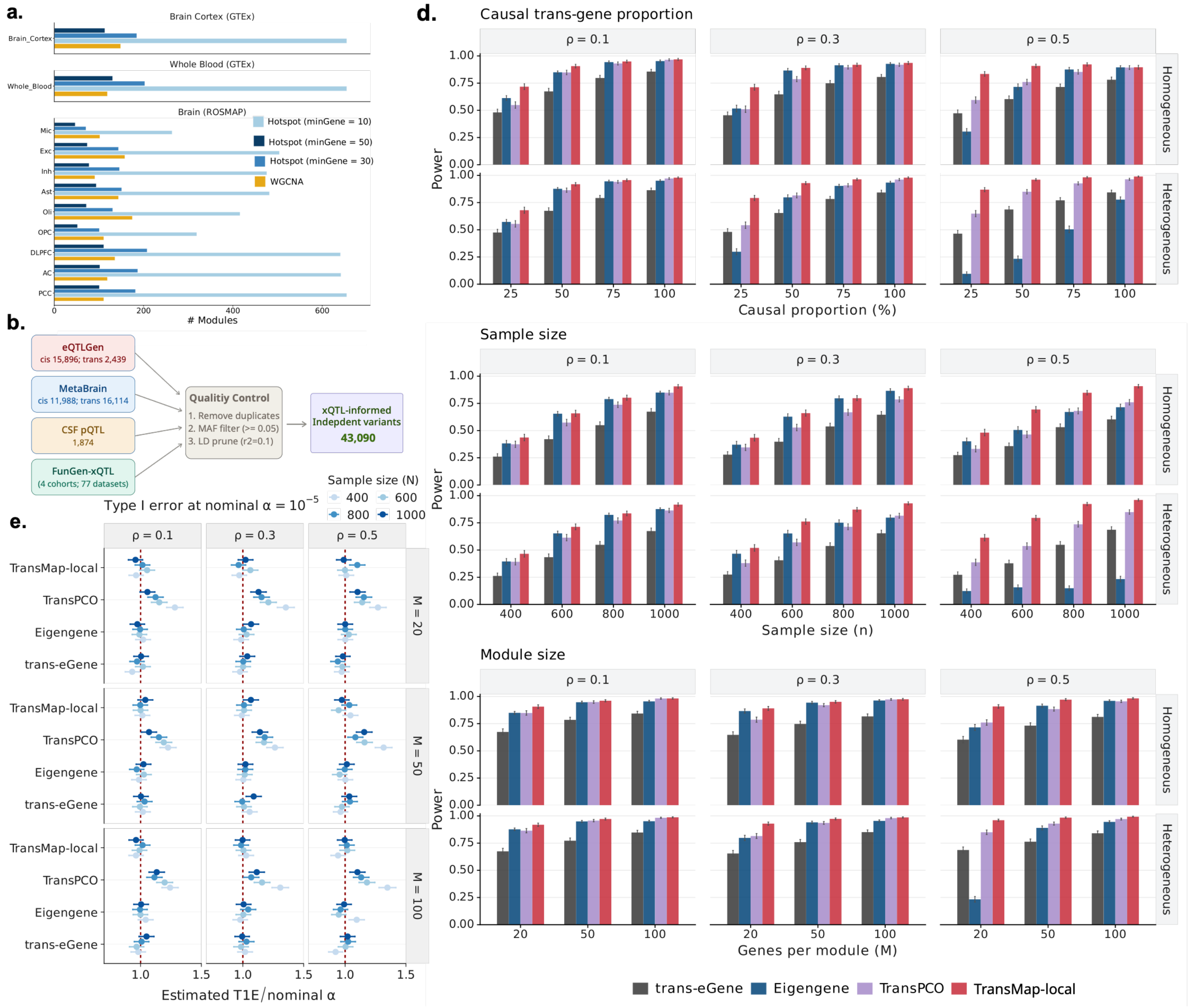

**Supplementary Figure 2**. **Supporting analyses for global program benchmarking. a.** Minor allele frequency (MAF) distribution of variants from ROSMAP WGS data that used in the simulations. Dashed line indicates MAF=0.05. **b.** Area under the precision-recall curves (AUPRC) of recovering true variant-gene pairs from *TransMap-global* versus ARCHIE at per-SNP *trans* heritability levels and sparsity-parameter grid. Colors indicate per-SNP *trans* heritability and shapes indicate sparsity. **d.** AUPRC of recovering true variant-gene pairs from *TransMap-global* versus ARCHIE at per-SNP *trans* heritability levels and MAF intervals. Colors indicate per-SNP *trans* heritability and shapes indicate MAF intervals. In **c** and **d**, the dashed line denotes equal performance. Numerical results are reported in Supplementary Data.

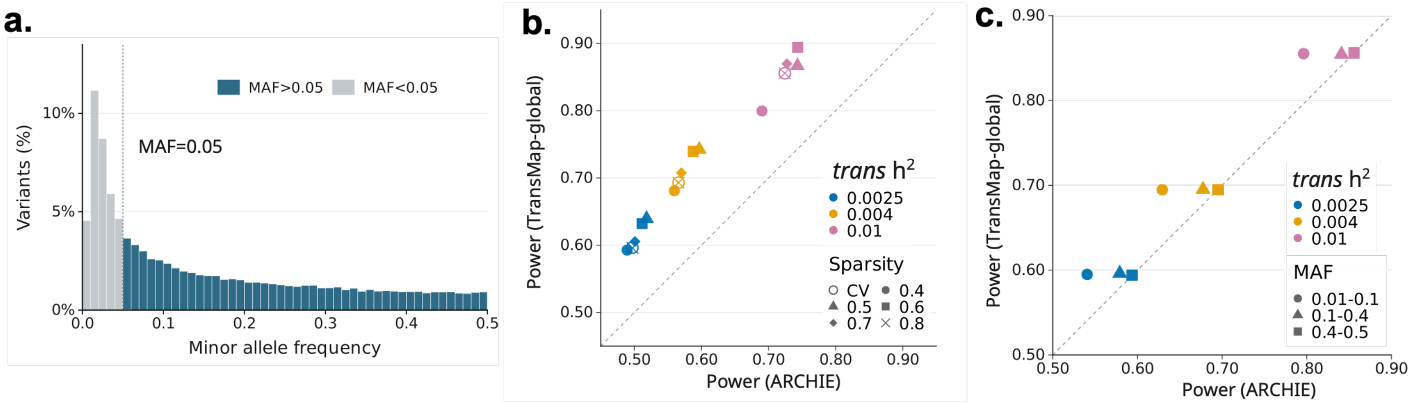

**Supplementary Figure 3. Supporting analyses for genome-wide *trans*-eQTL discovery and *TransMap-eGene* annotation. a.** Number of *trans*-eQTL identified across ROSMAP expression contexts after adjustment for different numbers of phenotype principal components (PCs) along with biological and genotype PC covariates. Red dashed lines indicate the corresponding analysis without any covariate adjustment. See Supplementary Note for details of covariates consideration. **b.** Genome-wide empirical FDR estimates for SuSiE fine-mapping and ColocBoost multi-phenotype colocalization under phenotype-wise permutation, which disrupted covariance among *trans* phenotypes, and genotype-label permutation, which preserved the observed phenotype covariance while disrupting genotype–phenotype associations. Results are shown for the overall empirical FDR, the empirical FDR weighted by the number of phenotypes tested and the empirical *cis*–*trans* FDR obtained by matching permuted distal events to observed *cis*-eQTL colocalization events. Numerical results are reported in Supplementary Data.

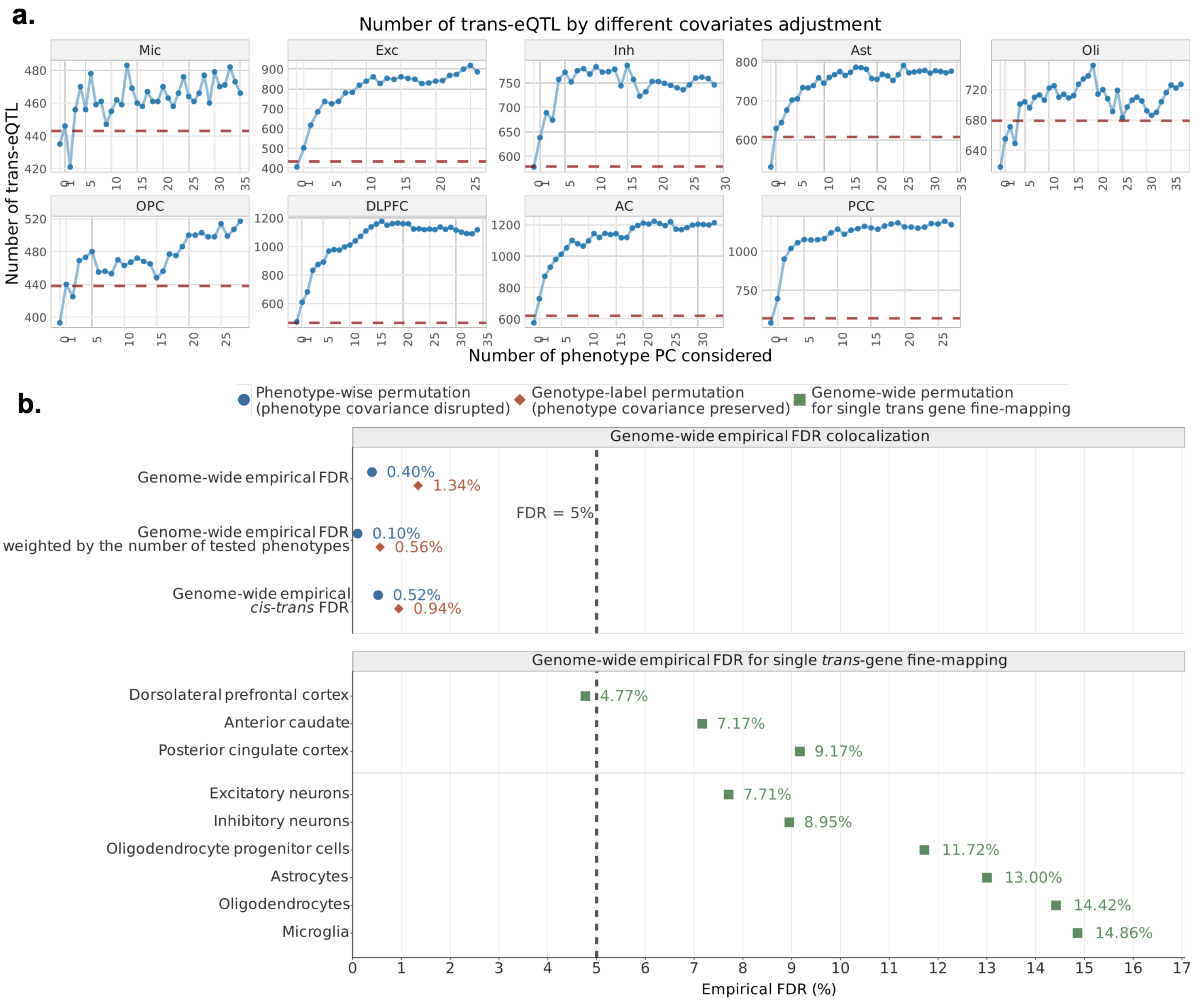

**Supplementary Figure 4. Disease-heritability analyses of local and global *trans*-regulatory program annotations. a.** Heritability enrichment and conditional standardized effect size (joint $\tau^*$) of *TransMap-local*, TransPCO, Eigengene and AUCell *trans*-MaxPIP annotations derived from Hotspot or WGCNA modules. **b.** Heritability enrichment, marginal $\tau^*$ and joint $\tau^*$ of *TransMap-local*, native TransPCO and TransPCO* annotations, restricted to the 43,090 xQTL-informed variants. TransPCO* denotes annotation obtained by genome-wide fine-mapping of TransPCO-selected principal components (Methods). **c.** Marginal $\tau^*$, heritability enrichment and joint $\tau^*$ of *TransMap-global* and ARCHIE annotations. **d.** Heritability enrichment and marginal standardized effect size ($\tau^*$) of *TransMap-eGene* annotations derived from genome-wide fine-mapping, hit-based fine-mapping or marginal association testing (left) and number of *trans*-eQTL retained by the genome-wide *TransMap-eGene* fine-mapping compared with *trans*-eQTL selected by hit-based fine-mapping or marginal association testing across expression contexts (right). Dashed line denotes equality. **e.** Heritability explained by fine-mapped *cis*-MaxPIP versus the union of *trans*-MaxPIP combined from *TransMap-eGene* and *TransMap-local* across 28 brain-related traits, dashed line denotes equality. In **a–d**, results are meta-analyzed across all 57 traits, 18 brain-related traits and 22 blood-related traits. S-LDSC models were conditioned on 53 baseline-LD annotations, with joint models additionally including all annotations shown in the corresponding panel. Error bars indicate 95% confidence intervals, and asterisks indicate statistical significance at FDR<5%. Numerical results are reported in Supplementary Data.

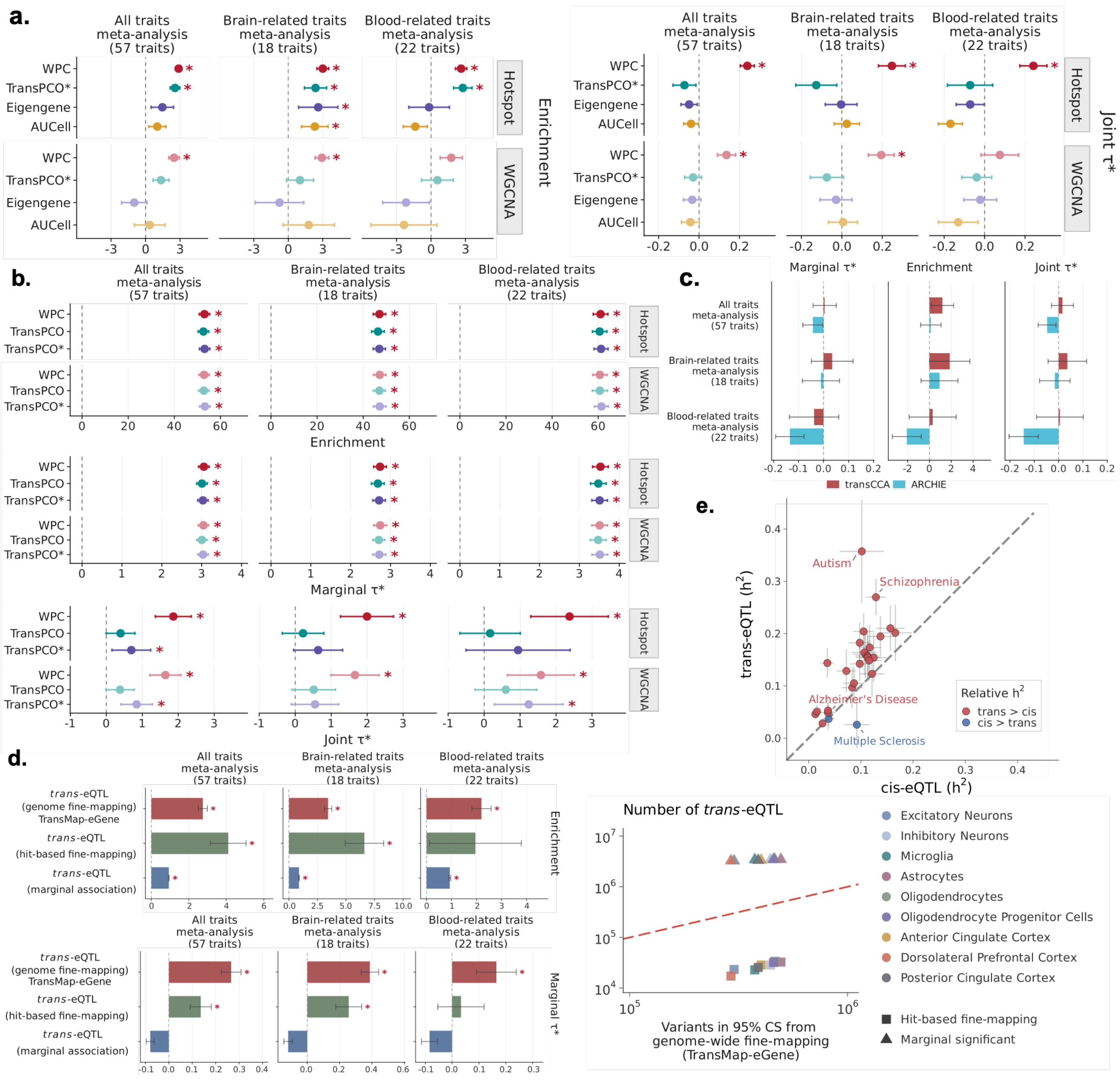

**Supplementary Figure 5. Split-sample reproducibility of *TransMap* annotations and empirical null calibration of multi-trait colocalization. a.** Pearson correlations between variant-level *trans*-MaxPIP annotations independently derived from two non-overlapping 50:50 donor splits for the *TransMap-eGene*, *TransMap-local* and *TransMap-global* scales. **b.** Heritability enrichment (top) and marginal standardized effect size ($\tau^*$; bottom) of split-specific *trans*-MaxPIP annotations. Heritability enrichment, marginal standardized effect size ($\tau^*$) and conditional $\tau^*$ from joint S-LDSC models for *trans*-MaxPIP annotations derived from **c.** 50 MSigDB Hallmark gene sets and **d.** cell-type-resolved, data-driven SCENIC regulons using *TransMap*, TransPCO*, AUCell or Eigengene representations. Corresponding analyses. **e.** Heritability enrichment, marginal $\tau^*$ and joint $\tau^*$ of the Hallmark and SCENIC *TransMap* annotations modeled together with *TransMap-eGene*-MaxPIP and *TransMap-local*-MaxPIP annotations. S-LDSC results are meta-analyzed across all 57 traits, 18 brain-related traits and 22 blood-related traits, conditional on 53 annotations; joint models additionally included all annotations shown in the corresponding panel. Error bars indicate 95% confidence intervals, and asterisks indicate statistical significance. Numerical results are reported in Supplementary Data.

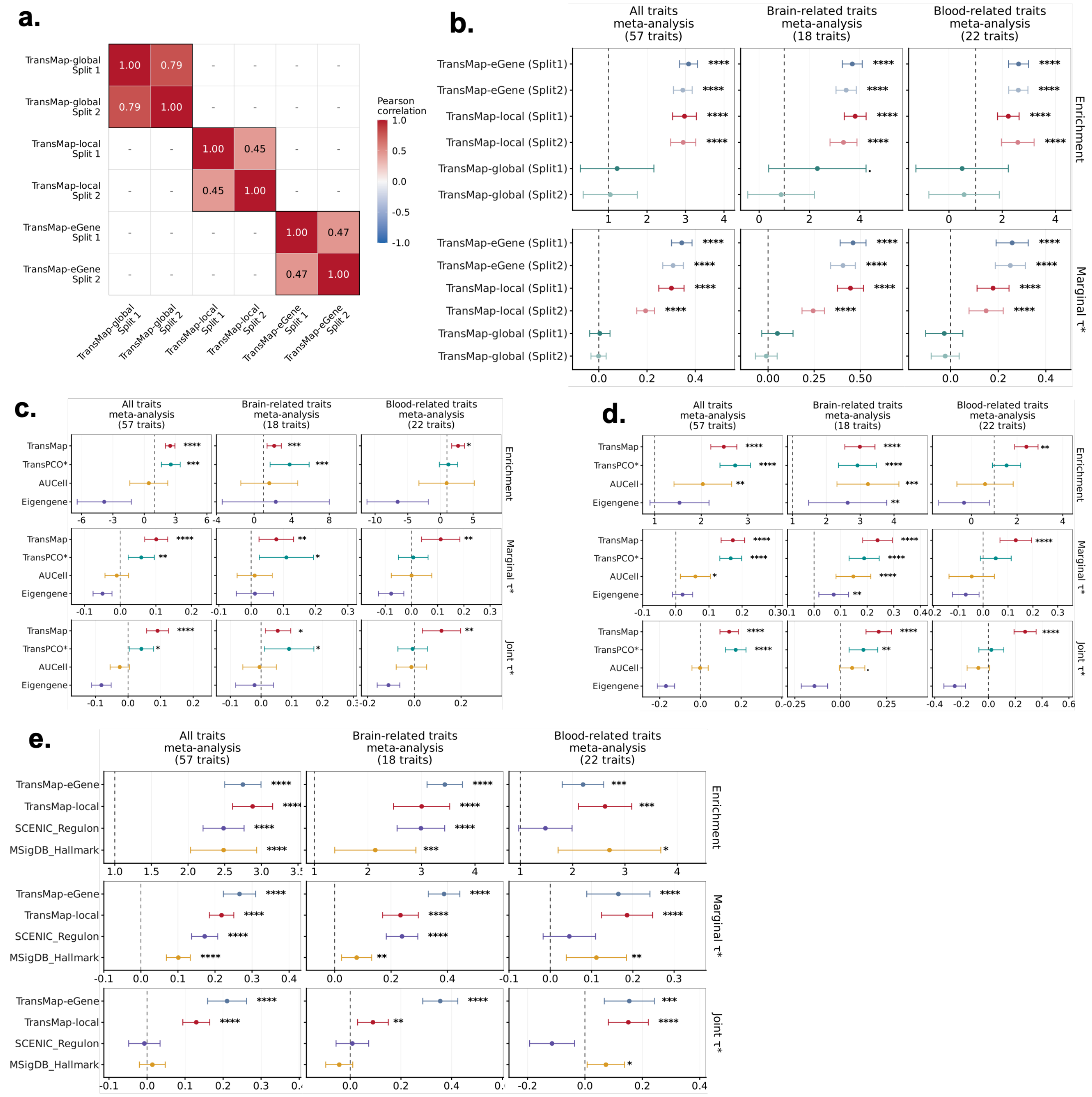

**Supplementary Figure 6. Genome-wide colocalization patterns and functional characterization of *TransMap trans*-regulatory loci. a.** UpSet plot of genome-wide colocalization events involving *cis*-eQTL, *trans*-eGene, *trans*-eProgram, protein-QTL, glycoprotein-QTL and metabolite-QTL. **b.** Log2 odds ratios of *trans*-eQTL from fine-mapping and colocalization analyses among external PsychAD and MetaBrain *trans*-eQTL associations with 95% confidence intervals. **c.** Proportion of *cis*-eQTL supported by different number of downstream *trans*-regulatory layers. **d.** Sankey plot linking trans-eQTL loci supported by *trans*-eGene or *trans*-eProgram to three modalities of biomarker-QTL. **e.** Enrichment and recall of cell-type-resolved *trans*-eQTL for overlap with protein, metabolite and glycoprotein QTL. Numerical results are reported in Supplementary Data.

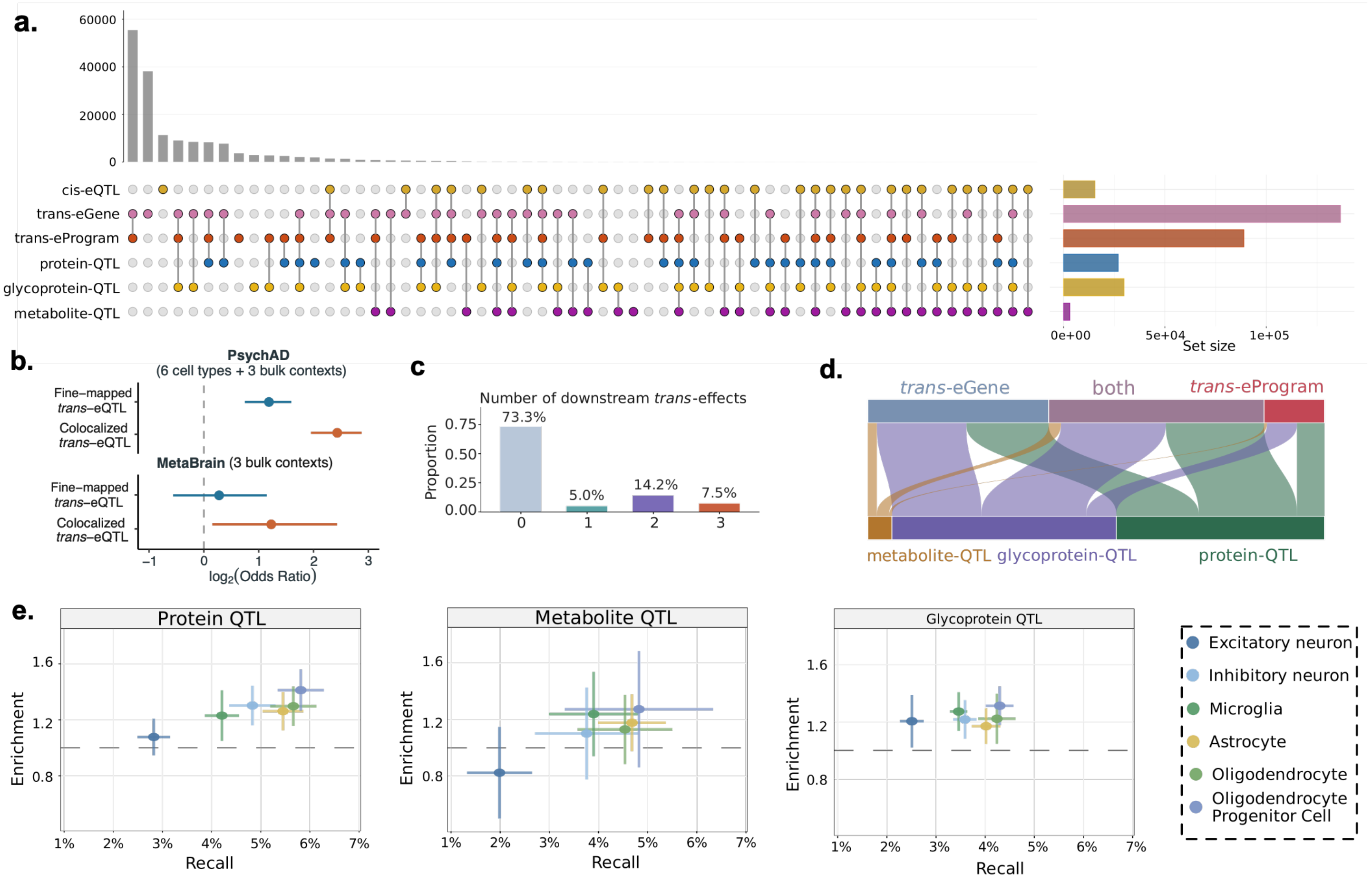

**Supplementary Figure 7. Genome-wide colocalization patterns and functional characterization of *TransMap trans*-regulatory loci. a.** Enrichment for functional gene properties of a curated set of 1,639 human transcription factors compared with: genes nearest *cis-only*, *cis-trans*, *trans-only* loci in term of transcription start site (TSS) and *cis*-eGenes for *cis-only* and *cis-trans* loci. **b.** Distribution of distance to the TSS of nearest gene for *cis-only*, *trans-only* and *cis-trans* loci. **c-d.** Enrichment for evolutionary constraint of three classes' **c.** variants for highly conserved sequence (PhastCons>0.9), and **h.** genes nearest to each loci (nearest TSS genes) and *cis*-eGenes at *cis-only* and *cis-trans* loci for loss-of-function constrained genes (pLI>0.9). In **a,c,**d, results are shown separately for *trans*-eGene (top) and *trans*-eProgram (bottom). Asterisks indicate statistical significance at FDR<5%. Error bars indicate 95% confidence intervales calculated by leave-one-chromosome-out jackknife. Numerical results are reported in Supplementary Data.

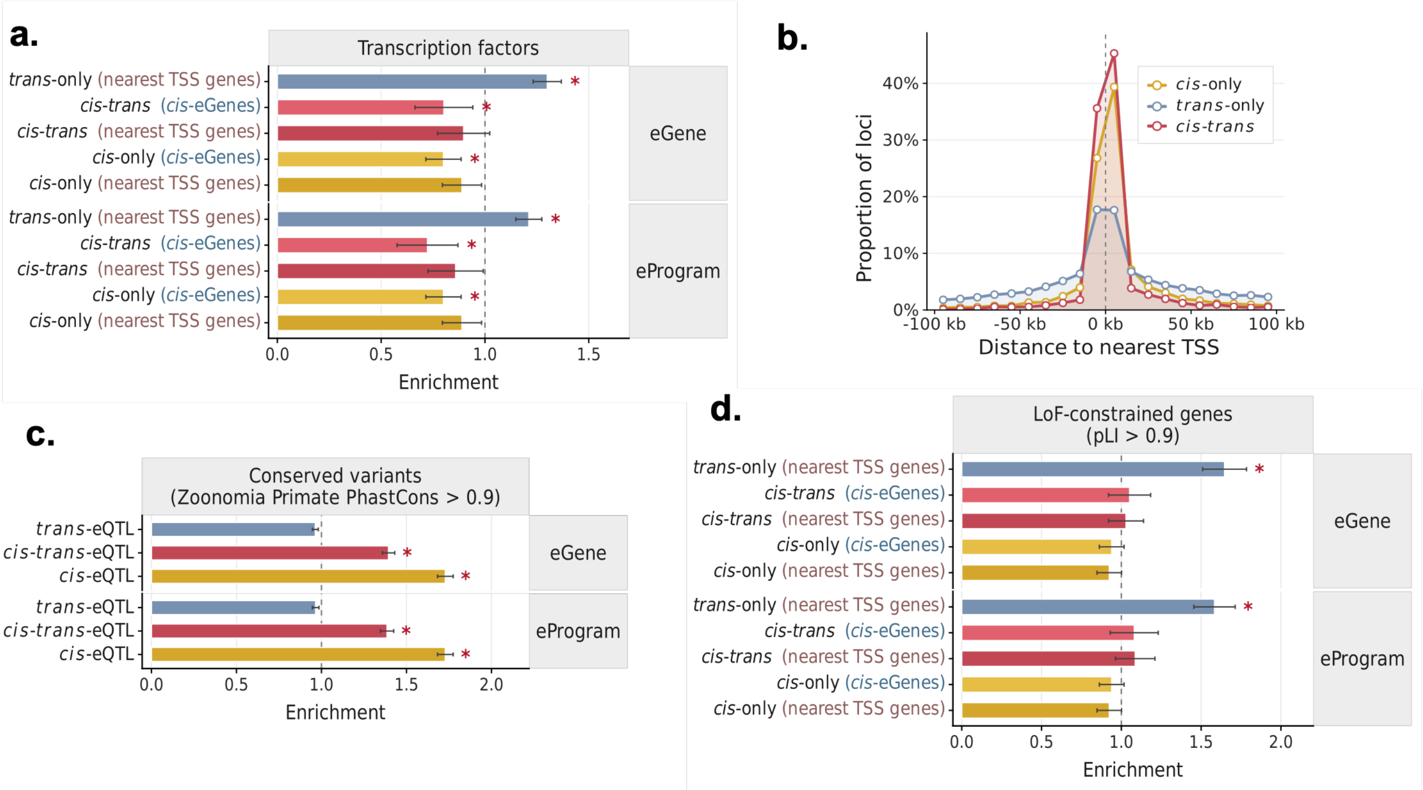

**Supplementary Figure 8. Cell-type architecture and robustness of neuron-initiated glial *trans*-regulatory paths. a.** Percentage of colocalized loci involving each brain cell type through a *cis*-eQTL or *trans*-eQTL signal. **b.** Enrichment of glial recipient cell types among candidate regulatory paths initiated by excitatory-neuron (left) or inhibitory-neuron (right) *cis* signals. **c.** Number of candidate cross-cell-type *cis*–*trans* loci for each initiating and recipient cell-type pair. **d.** Corresponding locus counts shown separately for *trans*-eGene and *trans*-eProgram supported paths. **e.** Enrichment of initiating–recipient cell-type pairs relative to a null distribution preserving cell-type representation and locus-level phenotype multiplicity, shown separately for *trans*-eGenes and *trans*-ePrograms. **f.** Numbers of excitatory-neuron *cis*-eGene and *trans*-eGene credible sets identified using the complete dataset and subsets containing 100,000, 200,000 or 300,000 excitatory-neuron nuclei. **g.** Numbers of candidate paths connecting excitatory-neuron *cis* signals to glial *trans* signals (heatmaps) and numbers of *cis*-eGene and *trans*-eGene loci identified in each cell type (bar plots) after excitatory-neuron subsampling. **h.** Distribution of donor-level cell fractions for the six major brain cell types used in cell-proportion-QTL mapping. **i.** Proportion of candidate neuronal-*cis*–glial-*trans* colocalization events overlapping cell-proportion QTL in the complete dataset and excitatory-neuron subsamples. Error bars indicate 95% confidence intervals, and asterisks indicate statistical significance. Numerical results are reported in Supplementary Data.

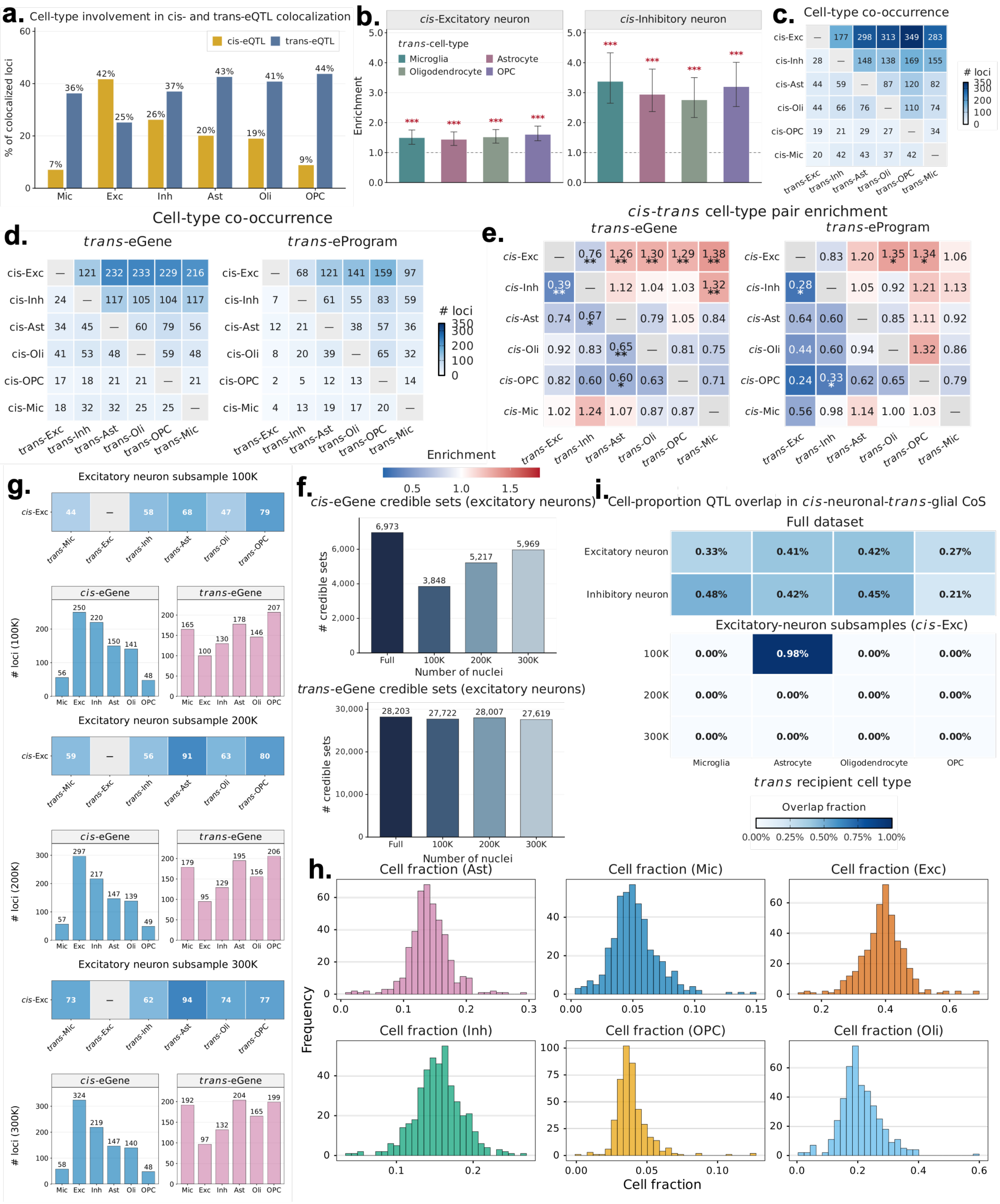
a.
Cell-type involvement in cis- and trans-eQTL colocalization
cis-eQTL
trans-eQTL
% of colocalized loci
7%
36%
42%
25%
26%
37%
20%
43%
19%
41%
9%
44%
Mic
Exc
Inh
Ast
Oli
OPC
b.
cis-Excitatory neuron
cis-Inhibitory neuron
trans-cell-type
Microglia
Astrocyte
Oligodendrocyte
OPC
Enrichment
c.
Cell-type co-occurrence
cis-Exc — 177 298 313 349 283
cis-Inh 28 — 148 138 169 155
cis-Ast 44 59 — 87 120 82
cis-Oli 44 66 76 — 110 74
cis-OPC 19 21 29 27 — 34
cis-Mic 20 42 43 37 42 —
trans-Exc
trans-Inh
trans-Ast
trans-Oli
trans-OPC
trans-Mic
### loci
d.
Cell-type co-occurrence
trans-eGene
cis-Exc — 121 232 233 229 216
cis-Inh 24 — 117 105 104 117
cis-Ast 34 45 — 60 79 56
cis-Oli 41 53 48 — 59 48
cis-OPC 17 18 21 21 — 21
cis-Mic 18 32 32 25 25 —
trans- eProgram
cis-Exc — 68 121 141 159 97
cis-Inh 7 — 61 55 83 59
cis-Ast 12 21 — 38 57 36
cis-Oli 8 20 39 — 65 32
cis-OPC 2 5 12 13 — 14
cis-Mic 4 13 19 17 20 —
trans-Exc
trans-Inh
trans-Ast
trans-Oli
trans-OPC
trans-Mic
### loci
e.
cis-trans cell-type pair enrichment
trans-eGene
cis-Exc — 0.76 1.26 1.30 1.29 1.38
cis-Inh 0.39 — 1.12 1.04 1.03 1.32
cis-Ast 0.74 0.67 — 0.79 1.05 0.84
cis-Oli 0.92 0.83 0.65 — 0.81 0.75
cis-OPC 0.82 0.60 0.60 0.63 — 0.71
cis-Mic 1.02 1.24 1.07 0.87 0.87 —
trans-eProgram
cis-Exc — 0.83 1.20 1.35 1.34 1.06
cis-Inh 0.28 — 1.05 0.92 1.21 1.13
cis-Ast 0.64 0.60 — 0.85 1.11 0.92
cis-Oli 0.44 0.60 0.94 — 1.32 0.86
cis-OPC 0.24 0.33 0.62 0.65 — 0.79
cis-Mic 0.56 0.98 1.14 1.00 1.03 —
trans-Exc
trans-Inh
trans-Ast
trans-Oli
trans-OPC
trans-Mic
Enrichment
g.
Excitatory neuron subsample 100K
cis-Exc 44 — 58 68 47 79
trans-Mic trans-Exc trans-Inh trans-Ast trans-Oli trans-OPC
cis-eGene
trans-eGene
### loci (100K)
56 250 220 150 141 48
165 100 130 178 146 207
Mic Exc Inh Ast Oli OPC
Excitatory neuron subsample 200K
cis-Exc 59 — 56 91 63 80
cis-eGene
trans-eGene
### loci (200K)
57 297 217 147 139 49
179 95 129 195 156 206
Excitatory neuron subsample 300K
cis-Exc 73 — 62 94 74 77
cis-eGene
trans-eGene
### loci (300K)
58 324 219 147 140 48
192 97 132 204 165 199
f.
cis-eGene credible sets (excitatory neurons)
### credible sets
6,973
3,848
5,217
5,969
Full
100K
200K
300K
Number of nuclei
trans-eGene credible sets (excitatory neurons)
28,203
27,722
28,007
27,619
i.
Cell-proportion QTL overlap in cis-neuronal-trans-glial CoS
Full dataset
Excitatory neuron 0.33% 0.41% 0.42% 0.27%
Inhibitory neuron 0.48% 0.42% 0.45% 0.21%
Excitatory-neuron subsamples (cis-Exc)
100K 0.00% 0.98% 0.00% 0.00%
200K 0.00% 0.00% 0.00% 0.00%
300K 0.00% 0.00% 0.00% 0.00%
Microglia
Astrocyte
Oligodendrocyte
OPC
trans recipient cell type
Overlap fraction
0.00% 0.25% 0.50% 0.75% 1.00%
h.
Cell fraction (Ast)
Cell fraction (Mic)
Cell fraction (Exc)
Cell fraction (Inh)
Cell fraction (OPC)
Cell fraction (Oli)
Frequency
Cell fraction

**Supplementary Figure 9. Pathway enrichment and orthogonal validation of candidate neuron–glia regulatory paths**. **a**. Reactome pathway enrichment of glycoproteins colocalized with excitatory neuron and astrocyte imitating-recipient events with FDR < 1%. **b**. STRING protein-protein interaction network of astrocyte trans-eGenes and glycoproteins from excitatory neuron and astrocyte imitating-recipient events. **c**. STRING network among all 25 genes annotated to RHO-ROCK and SEMA4D-induced cell migration pathways. In **b** and **c**, only proteins with association score > 0.4 are retained. **d.** Spatial differential expression patterns of neuronal *cis*-regulated genes between deep (L4–L6) and outer (L1–L3) cortical layers (left) and of their associated glial *trans*-regulated genes in white matter and cortical layers (right), stratified by AD pathological stage. **e.** Spatial organization of genes implicated through *trans*-ePrograms. The left panel shows differential expression of neuronal *cis*-regulated genes between deep and outer cortical layers; the right panel compares spatial differential expression evidence for recipient glial genes in white matter and cortical layers. Error bars indicate 95% confidence intervals, and asterisks indicate statistical significance. CTRL, cognitively normal control.

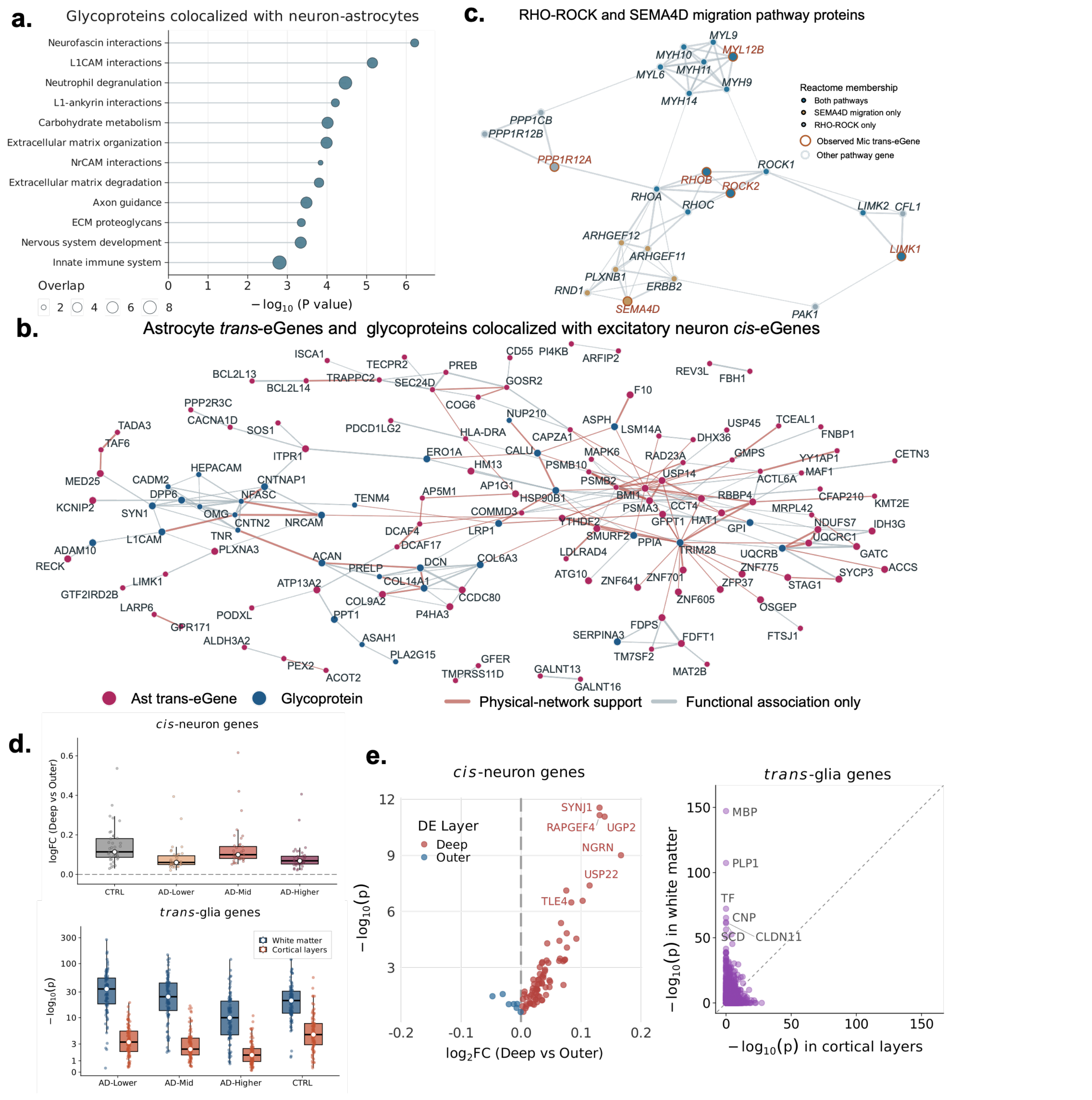

**Supplementary Figure 10. Supporting analyses at the *SEC61G* and *GDA* loci. a.** Reactome pathway enrichment for the microglial *trans*-eProgram (module 100) colocalized at the *SEC61G* locus. **b.** Colocalization plots at the *GDA* locus for the excitatory neuron *GDA cis*-eQTL, astrocyte *trans*-eProgram QTL, and 11 *trans*-glycosylation traits representing nine glycoproteins. Orange points mark variants in the reported 95% colocalization confident set. **c.** Pathway enrichment for the nine glycoproteins shown in panel b. **d.** Neuron-responsive astrocyte genes identified in co-culture experiment. Red points denote genes in *GDA* colocalized astrocyte program genes that overlap the co-culture response, blue points denote other neuron-responsive astrocyte genes (FDR < 0.01). The axes show $\log_2$ fold change and $-\log_{10}$(adjusted $P$). **e.** STRING PPI network of colocalized *trans*-glycoproteins and member genes in astrocyte *trans*-eProgram (Program 80). Orange nodes denote glycoproteins, blue nodes denote program genes also overlapped with neuron-responsive astrocyte genes in co-culture experiment, and grey nodes denote other program genes. Numerical results are reported in Supplementary Data.

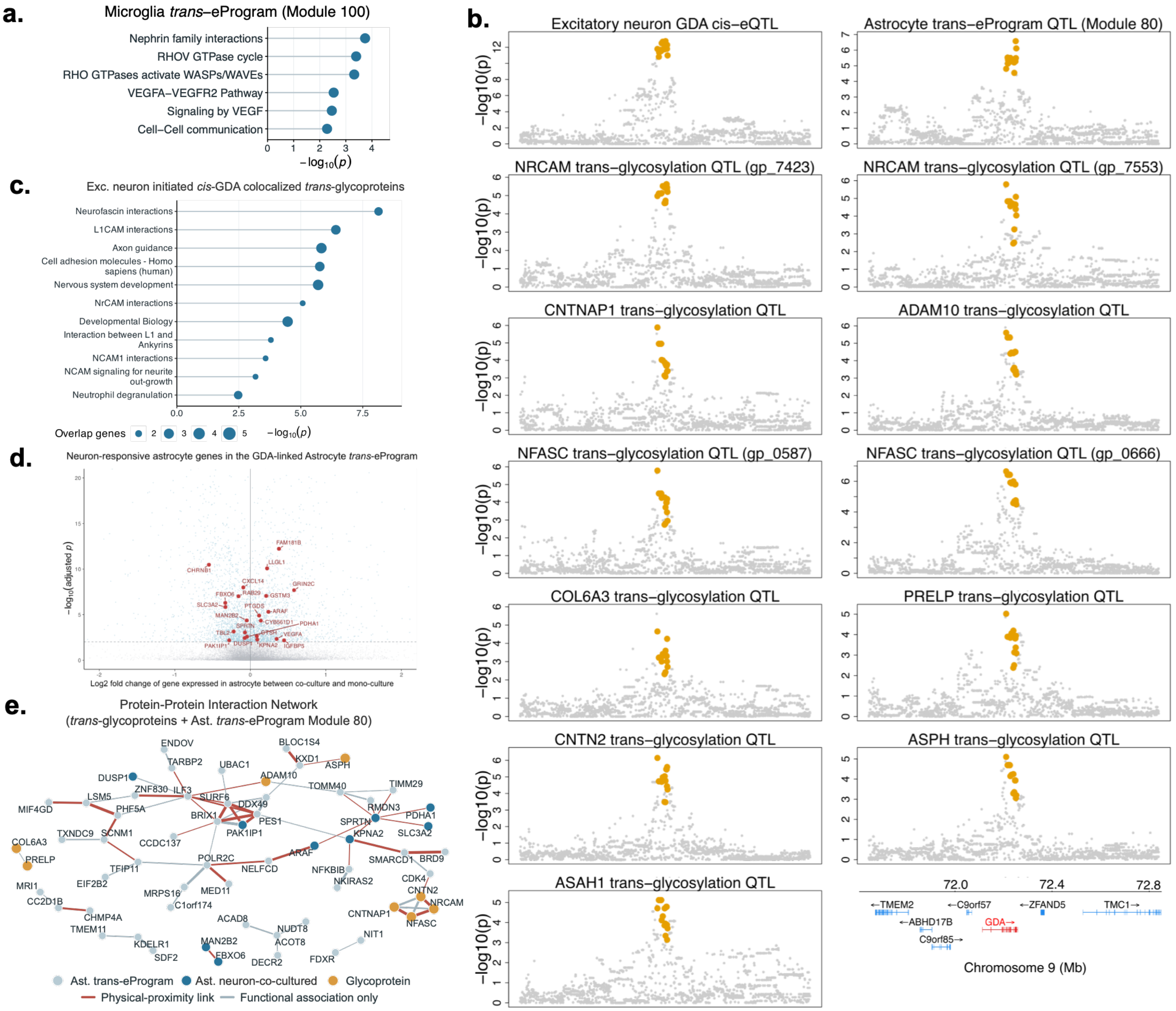

**Supplementary Figure 11. Multilayer and cell-type-resolved architecture of AD-associated colocalization events. a.** UpSet plot of 112 AD-centered colocalization events across AD GWAS fine-mapping, *cis*-eGene, *trans*-eGene, *trans*-eProgram, *trans*-biomarker, AD-pathology and cell-fraction QTL layers. **b.** Number of AD colocalized loci supported by *cis*-eQTL and *trans*-eQTL, colored coded by the number of snRNA-seq cell types implicated at each locus. **c.** Percentage of AD-colocalized loci involving each brain cell type through *cis*-eQTL or *trans*-eQTL evidence and **d.** separating *trans*-eGene and *trans*-eProgram evidence. **e.** Associations between Zoonomia phyloP conservation scores of lead colocalized variants and the presence of individual initiating cell types, recipient cell types or initiating–recipient cell-type pairs. Error bars indicate 95% confidence intervals. **f.** Manhattan plot of the AD GWAS meta-analysis with tracks indicating cell-type-resolved *cis*- and *trans*-eQTL evidence at each colocalized locus. Loci are labeled by the colocalized *cis*-eGene; asterisks indicate the nearest-gene label for *AD–trans-only* loci. Numerical results are reported in Supplementary Data.

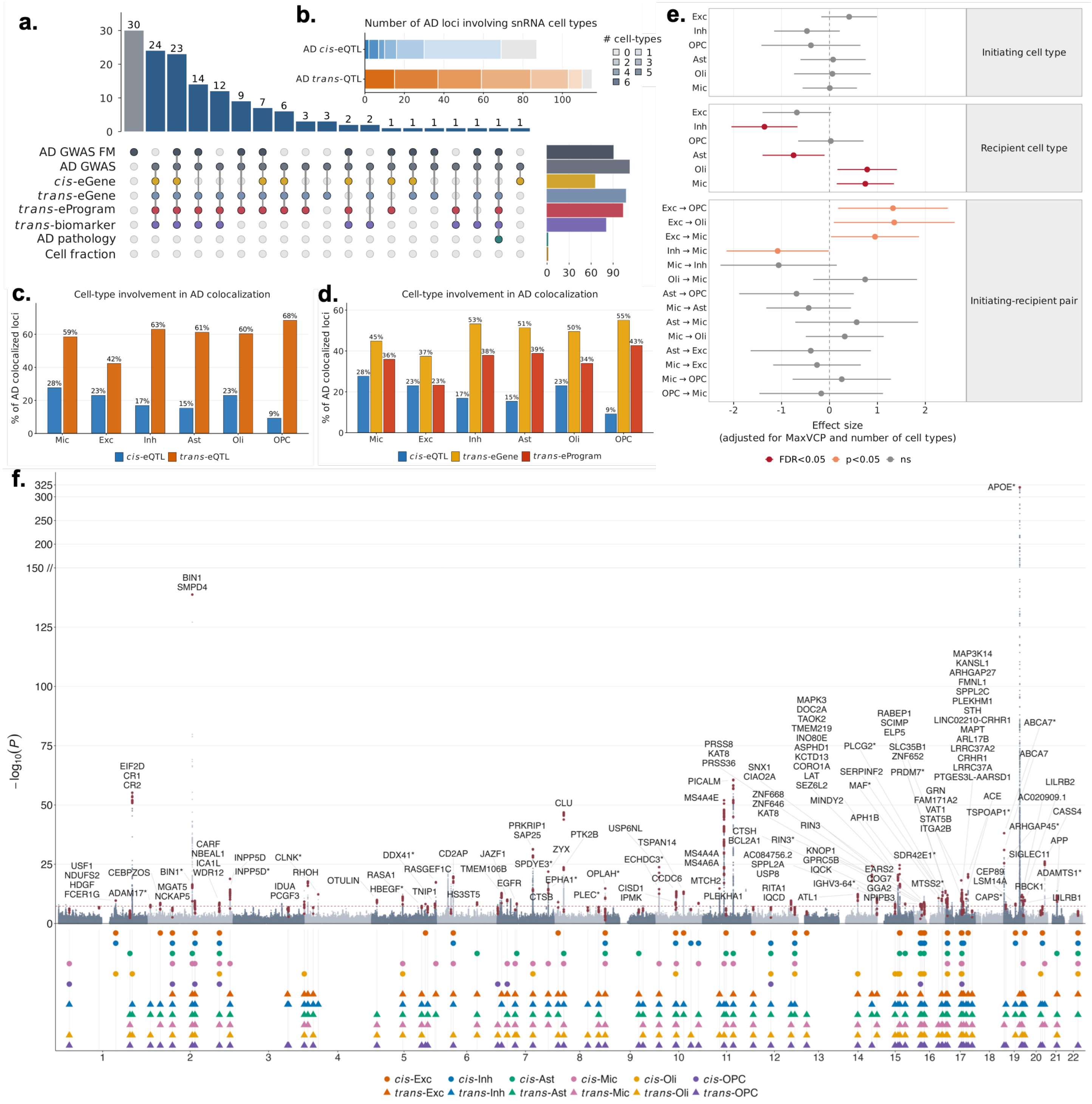

**Supplementary Figure 12. AD-centered *TransMap* colocalization identifies *cis*, *trans* and biomarker regulatory layers with AD GWAS loci shown in secondary AD GWAS meta-analysis. a.** UpSet plot of 69 distinct colocalization events from AD-target analysis, showing colocalization patterns across different modalities. Venn diagram shows the number of colocalization events supported by *cis*-eQTL and *trans*-eQTL. **b.** Number of AD colocalized loci supported by *cis*-eQTL and *trans*-eQTL, colored coded by the number of snRNA-seq cell types implicated at each locus. **c.** Variants conservation based on Zoonomia phyloP scores of the lead variant at each AD locus (highest MaxVCP for AD colocalized loci and MaxPIP for only AD fine-mapped loci). Asterisks denote significant differences between group (* FDR<5%, ** FDR<1%). **d.** Manhattan plot of the AD GWAS meta-analysis with colocalized loci labeled by *cis*-eGene or nearest gene at *AD-trans* only colocalized loci, and tracks below making the molecular QTL classes colocalized at each locus. Numerical results are reported in Supplementary Data.

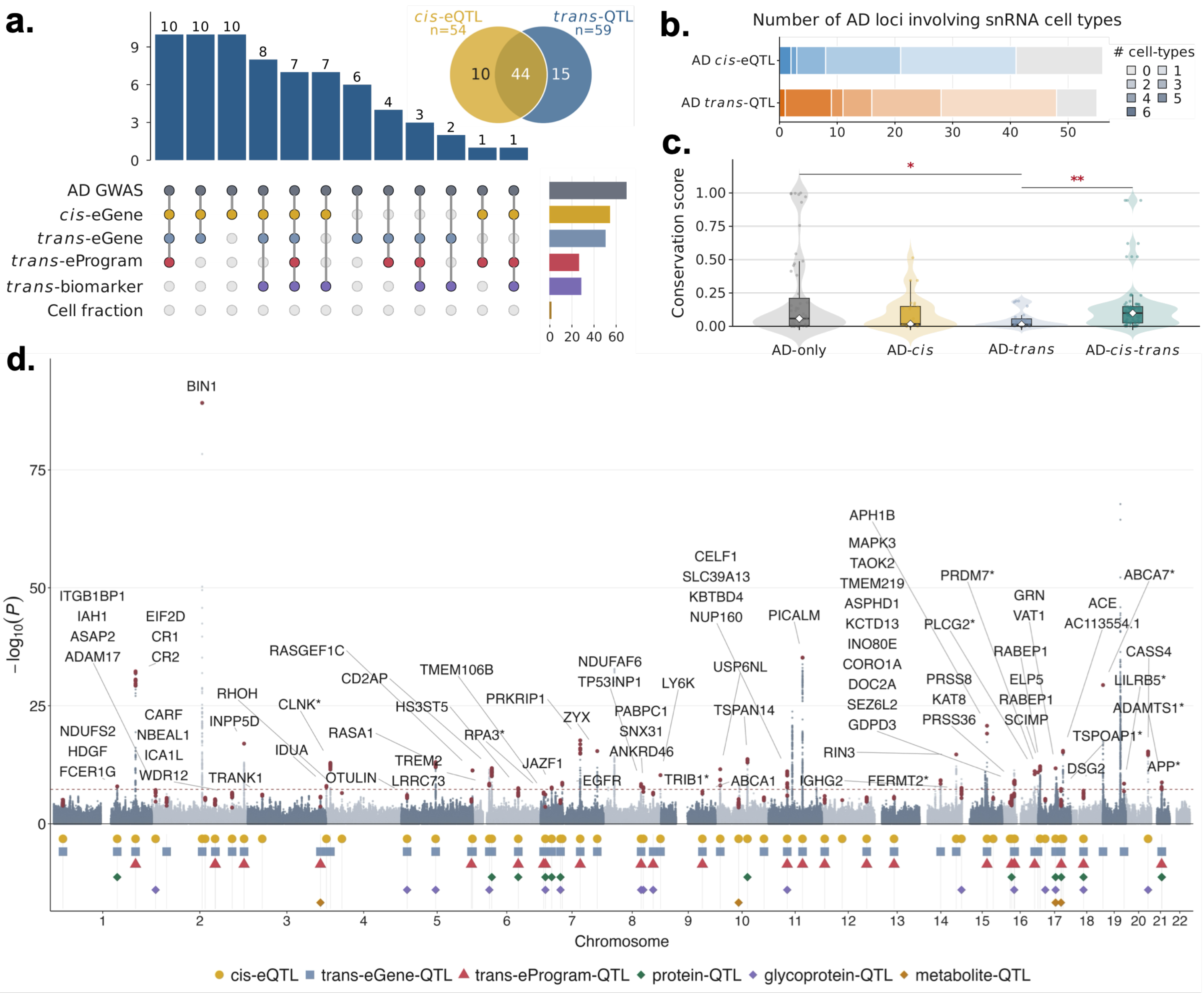

**Supplementary Figure 13. Supporting analyses of AD colocalized *cis-trans* signals at the *KNOP1* and *MS4A*-family gene loci. a.** Heatmap of indirect effect estimates within the chromosome 16 *KNOP1* colocalization set. Columns indicate proximal *cis*-eGenes by context and rows indicate distal *trans*-eGenes or *trans*-ePrograms by context. Color shows the signed indirect effects, asterisks mark nominal significant effects with p-value<0.05. **b.** Number of colocalized *trans* phenotypes by recipient context in the chromosome 11 *MS4A*-family gene loci colocalization. **c.** Pathway enrichment analysis for two colocalized inhibitory neuron *trans*-ePrograms with FDR<5%. **d-e.** STRING networks of inhibitory neuron *trans*-eProgram module 59 (**d**) and module 115 (**e**), respectively. In **d-e**, red edges denote physical proximity links, and gray edges denote functional associations only. Numerical results are reported in Supplementary Data.

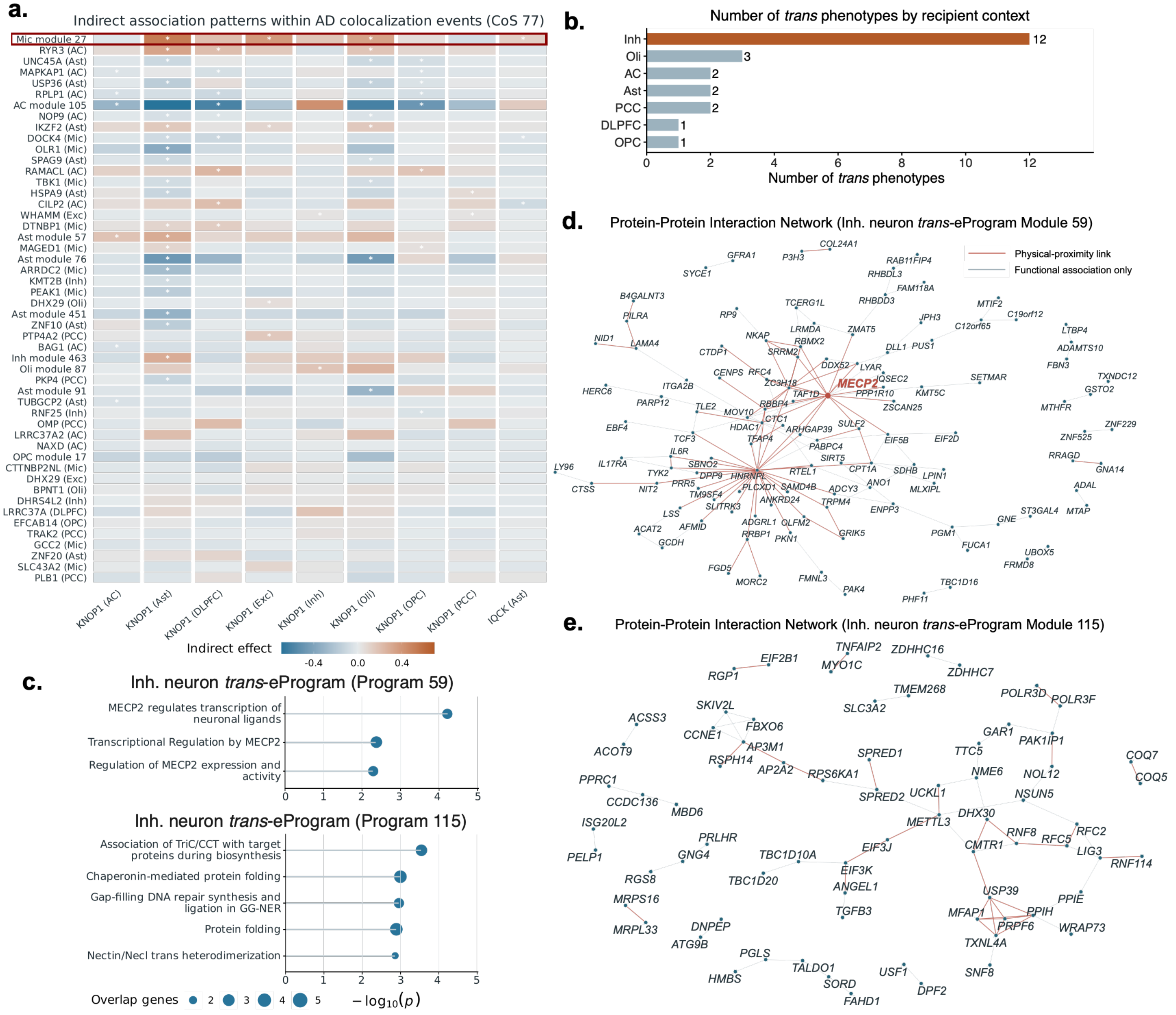

**Supplementary Figure 14. Supporting analyses of AD colocalized *cis-trans* signals at the 16p11.2 locus.** **a.** STRING PPI network of proteins encoded by genes in the *INO80E cis* region and the distal target *ABCA1*. **b.** Indirect associations between genotype of colocalized loci and local *cis*-eGenes, with *INO80E* as the mediator. **c.** Indirect associations with local *cis*-eGene as mediator and *INO80E* as the outcome. In **b-c**, points show estimated indirect effects with 95% confidence intervals. Numerical results are reported in Supplementary Data.

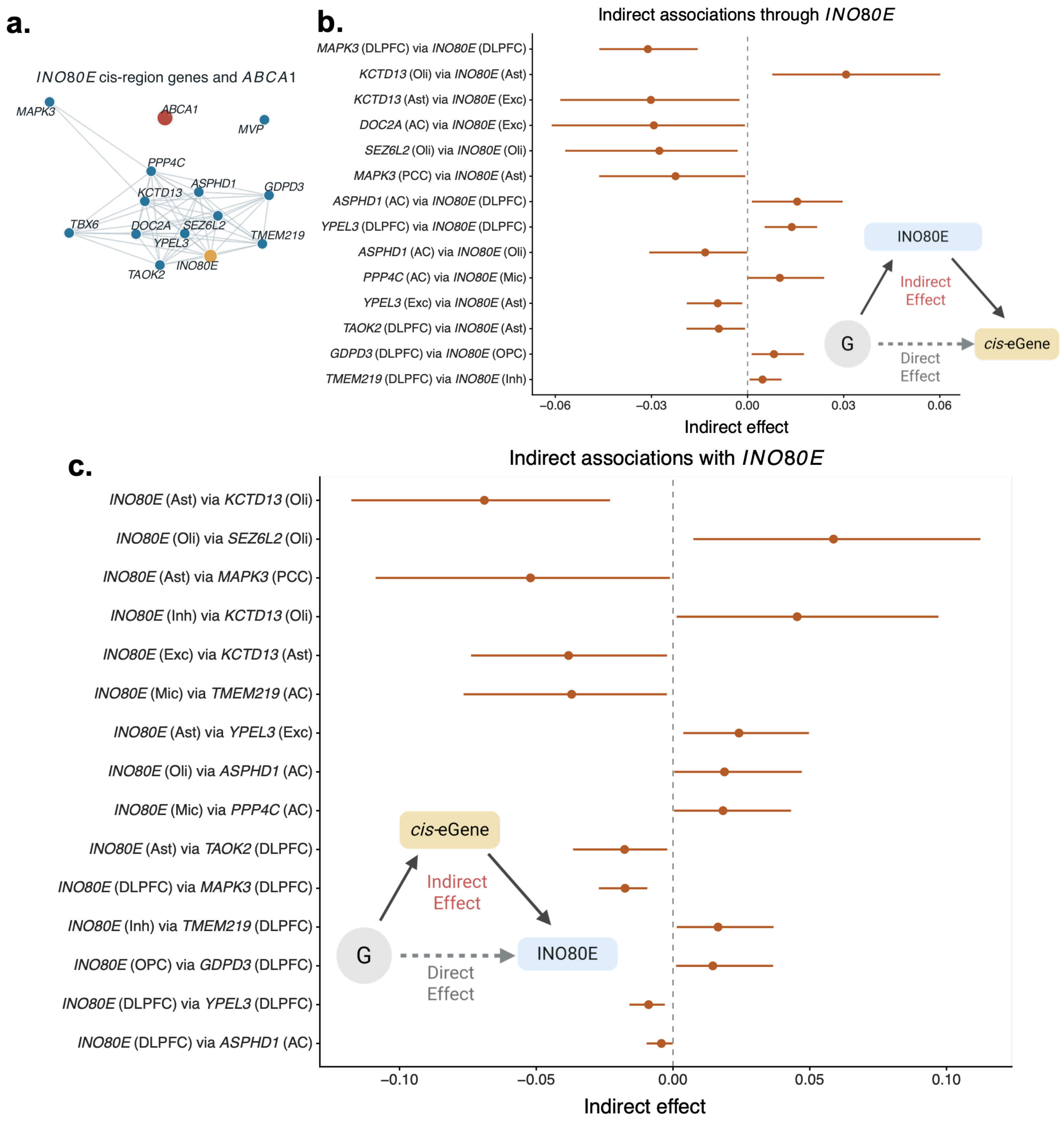
