## Supplementary Note for "A multi-scale cell-type resolved map of *trans*-regulatory architecture in the aging human brain"

### Supplementary Notes

#### S1. *Trans*-eQTL identifications from different studies

Large-scale *trans*-eQTL studies often preselected variants to limit the multiple testing burden of genome-wide variant-gene scans. MetaBrain performed *trans*-eQTL on 2,759 brain cortical samples and 228,819 variants with prior GWAS or *cis*-eQTL evidence[1]. By defining *trans* as more than 5Mb apart or on different chromosomes, it reported 737 *trans*-eQTL association involving 526 *trans*-eQTL and 108 *trans*-eGenes at FDR < 5%. eQTLGen Phase I similarly restricted *trans*-eQTL identifications to 10,317 trait-associated variants[2]. Using blood-derived expression data from up to 31,684 samples in 37 cohorts, it identified 59,786 significant *trans* variant-gene associations, involving 3,853 *trans*-eQTL and 6,298 *trans*-eGenes at FDR < 5%. In contrast, eQTLGen Phase II tested variants genome-wide across 43,301 samples from 52 datasets[3]. It reported 42,580 *trans* locus-gene associations at FDR < 5%, corresponding to nominal p-value $< 5 \times 10^{-8}$ from genome-wide scan. Then, they performed fine-mapping on those significant locus-gene associations and yielded 44,869 *trans*-eQTL involving 8,867 genes. For our program-coherence analyses, GTEx v8 supplied the brain cortex ($N = 208$) and whole blood ($N = 673$) expression profiles[4] used to constructed co-expression modules from Hotspot[5] and WGCNA[6], which were then evaluated against MetaBrain and eQTLGen Phase I *trans*-eQTL results. We directly used the normalized expression profiles and considered sex, age, genotype PCs, and PEER factors (20 for brain cortex and 60 for whole blood).

#### S2. Covariate adjustment and hidden factor exclusion

In genome-wide association studies (GWAS) and eQTL analysis, researcher routinely adjust for covariates as part of their analytical framework[7–9]. Adjustment of covariates in genomic studies serves two primary purposes: first, to control for potential confounding factors that may bias SNP effect estimates; and second, to improve statistical power by reducing residual variance in the outcome measure[10–12]. Common examples of covariate adjustments include the use of principal components derived from individual genotypes to account for population stratification[7,13], and principal components of gene expression profiles to capture technical batch effects[8,14,15]. The statistical intuition underlying power improvement is straightforward: when a true risk factor or source of variation is properly accounted for, the residual variance of the outcome decreases. This reduction in background noise increases the ratio of the true effect size of the predictor of interest relative to the total phenotypic variance, thereby enhancing statistical power to detect genuine associations.

We performed *trans*-eQTL analysis on 9 eQTL contexts from the ROSMAP cohort, comprising 6 pseudo-bulk eQTL and 3 bulk eQTL from brain regions. Since genome-wide *trans*-eQTL analysis would impose a substantial multiple testing burden considering billions of potential tests, we focused our analysis on 43,090 xQTL-informed independent variants (**Supplementary Note S3**). For each eQTL context, we removed covariates that incorporated technical factors (post-mortem interval), biological covariates (sex, age at death), the top 15 genotype principal components, and additional hidden confounders identified through PCA on normalized phenotype matrices (phenotype PCs). We repeated *trans*-eQTL mapping of 43,090 variants and consistent with sensitivity analyses in MetaBrain when excluded samples from AMPAD (*N*=1,755)[1]. We observed that the number and distribution of *trans* associations were broadly robust to the number of expression PCs included, with analyses excluding expression PCs yielding the most conservative number of *trans*-eQTL (**Supplementary Figure 3a**). Therefore, in our primary analysis, we did not include expression principal components as covariates because, for phenotypes constructed from expression covariance structure, these components may absorb biologically relevant program variation or induce collinearity with the derived phenotypes [REF].

#### S3. xQTL-informed independent variants selection

We assembled an xQTL-informed tagging variants for *TransMap-global* program learning and analyses restricted to previously implicated molecular-QTL variants (**Supplementary Figure 2b**). These tagging variants were selected from molecular-QTL resources used in the FunGen-xQTL project[16], indicating the prior evidence for molecular regulatory activity. We assembled the variants from multiple resources, including eQTLGen *cis*- and *trans*-eQTL[2], MetaBrain *cis*- and *trans*-eQTL[1], cerebrospinal fluid (CSF) protein QTL[17,18], and FunGen-xQTL *cis*-QTL across four cohorts and 77 molecular datasets[16]. In FunGen-xQTL resource, we retained top variants with the highest PIP of 95% credible sets from gene-level single-context fine-mapping of 77 molecular phenotypes using SuSiE[19], yielding 342,992 distinct variants. We retained top variants with the highest PIP of 95% credible sets from TAD region-level multi-context fine-mapping using mvSuSiE[20], yielding 38,516 and 6,643 distinct variants for ROSMAP and MSBB cohorts. We also processed xQTL-only multi-trait colocalization results using ColocBoost[21] from ROSMAP, MSBB, MiGA and Knight cohorts and retained the variant with the highest variant colocalization probability (VCP), yielding 75,381 distinct variants. For the eQTLGen dataset, we retained the top variants with the highest z-score per gene for *cis*-eQTL, yielding 15,896 distinct variants, and we matched reported *trans*-eQTL to ROSMAP genotypes, yielding 2,439 distinct variants. For the MetaBrain dataset, we matched reported *cis*- and *trans*-eQTL to ROSMAP genotypes, yielding 11,988 and 16,114 distinct variants, respectively. Finally, we retained common variants (MAF > 0.05) in the ROSMAP WGS reference panel and performed LD pruning with PLINK2 (`--indep-pairwise 50 1 0.2`) for all distinct variants, yielding 43,090 approximately independent xQTL-informed variants.

### S4. Detailed construction of local and glocal programs from *TransMap*

#### S4.1. Local co-expression module identification with Hotspot

*TransMap-local* identified co-expression modules for each cell type and bulk context using Hotspot[5]. Standardized doner-level expression matrix $\boldsymbol{E}$ were restricted to genes present in the protein-coding. Covariates were removed from expression before module detection. We used full donor-level matrix as the space used to measure donor similarity to obtain a stable estimation[5]. For each context, we estimated an unweighted donor-neighborhood graph with `n_neighbors=n/2` and `approx_neighbors=False` to make sure the reproducible results. Informative genes were selected as the significant local autocorrelation with Benjamini-Hochberg FDR<5%. We then grouped the retained informative genes by average linkage clustering of their local correlation z-scores as default[5]. To evaluate local program definitions across module granularity, from fine-grained co-expression substructures to broader co-expression genes, we specified `min_gene_threshold` to 10, 30 and 50. This threshold sets the minimum size for a module and prevents two branches that have each reached that size from being merged. Module assignments were then used to constructed WPC-based local program phenotypes from a LOCO strategy.

#### S4.2. Whitened principal components and empirical background estimation

Let $\{\mathcal{G}_m\}_{m=1}^{M}$ denote the $M$ co-expression modules identified from Hotspot in each cell type or bulk context, where $\mathcal{G}_m$ indexes the genes in module $m$. In *TransMap-local,* we excluded genes on the same chromosome from each module when testing of variants on chromosome $c$ to ensure genuine *trans* effects. To simplify notation, we ignored index of chromosome $c$ in this subsection. Let $\boldsymbol{E}_m$ denote the donor-by-gene expression matrix for module $m$ and define its covariance as $\widehat{\boldsymbol{\Sigma}}_m = \boldsymbol{E}_m^\top \boldsymbol{E}_m/(n-1)$.

##### S4.2.1. Conceptual rationale for standard and whitening principal components analysis

Ordinary principal components analysis (PCA) orders directions by their total module variance, irrespective of how much variance is expected from residual co-expression. For the $k$-th component, the loading vector is estimated by maximizing total variance:

$$\widehat{\boldsymbol{\omega}}_{m,k} = \underset{\substack{\boldsymbol{\omega}^\top\boldsymbol{\omega}=1 \\ \boldsymbol{\omega}^\top\widehat{\boldsymbol{\omega}}_{m,\ell}=0,\ \ell<k}}{\operatorname{argmax}} \quad \boldsymbol{\omega}^\top \widehat{\boldsymbol{\Sigma}}_m \boldsymbol{\omega},$$

where the corresponding eigenvalue are obtained from $\widehat{\boldsymbol{\Sigma}}_m \widehat{\boldsymbol{\omega}}_{m,k} = \hat{\lambda}_{m,k}\widehat{\boldsymbol{\omega}}_{m,k}$. However, when $\widehat{\boldsymbol{\Sigma}}_m$ has a dominant eigendirection from a shared residual correlation rather than genetic correlation, the 1-sth PC of $\widehat{\boldsymbol{\Sigma}}_m$ aligns with that residual direction rather than genetic effects.

Instead of maximizing total variance, let $\boldsymbol{\Sigma}_{0m}$ denote the matched-size background covariance for the same genes, which can be estimated and described below (**Supplementary Note S4.2.4**). Whitening PC (WPC) analysis instead compares observed and background variance along each direction[22]. For the $k$-th component, the loading vector is estimated by maximizing signal-to-noise ratio (SNR):

$$\widehat{\boldsymbol{\omega}}_{m,k} = \underset{\substack{\boldsymbol{\omega}^\top \boldsymbol{\Sigma}_{0m}\boldsymbol{\omega}=1 \\ \boldsymbol{\omega}^\top \boldsymbol{\Sigma}_{0m}\widehat{\boldsymbol{\omega}}_{m,\ell}=0,\ \ell<k}}{\operatorname{argmax}} \frac{\boldsymbol{\omega}^\top \widehat{\boldsymbol{\Sigma}}_m \boldsymbol{\omega}}{\boldsymbol{\omega}^\top \boldsymbol{\Sigma}_{0m}\boldsymbol{\omega}}.$$

The denominator normalizes each projected score by the background covariance, whereas the numerator measures module variance along the same direction. Therefore, WPC prioritizes axes whose variation exceeds the residual background rather than axes with the largest raw expression variance. The $k$-th WPC loading vector and corresponding eigenvalue are obtained from the generalized eigenvalue problem

$$\widehat{\boldsymbol{\Sigma}}_m \widehat{\boldsymbol{\omega}}_{m,k} = \hat{\lambda}_{m,k} \boldsymbol{\Sigma}_{0m} \widehat{\boldsymbol{\omega}}_{m,k}.$$

Equivalently, the background-normalized covariance matrix $\boldsymbol{A}_m = \boldsymbol{\Sigma}_{0m}^{-1/2} \widehat{\boldsymbol{\Sigma}}_m \boldsymbol{\Sigma}_{0m}^{-1/2}$ . Its eigenvalues are the generalized eigenvalues $\hat{\lambda}_{m,k}$. If $\widehat{\boldsymbol{u}}_{m,k}$ is a unit eigenvector of $\boldsymbol{A}_m$, the corresponding loading in the original expression space is $\widehat{\boldsymbol{\omega}}_{m,k} = \boldsymbol{\Sigma}_{0m}^{-1/2} \widehat{\boldsymbol{u}}_{m,k}$. This formulation makes explicit that module variation is evaluated relative to the independently estimated residual background.

#### S.4.2.2. One genetic signal model

Consider a simple case that a module of $q_m$ genes in module $\mathcal{G}_m$ are affected by a single and unique *trans*-eQTL. Without loss of generality, for individual $i = 1, \cdots, n$, the expression vector $\boldsymbol{E}_m$ follows:

$$\boldsymbol{E}_{i,m} = \boldsymbol{\beta}_m G_i + \boldsymbol{\varepsilon}_i, \boldsymbol{\varepsilon}_i \sim N(\mathbf{0}, \boldsymbol{\Sigma}_{0m}),$$

where $G_i$ is the genotype of *trans*-eQTL for individual $i$ and $\boldsymbol{\beta}_m = \left(\beta_{1,m}, \cdots, \beta_{q_m,m}\right)^\top$ is the vector of standardized *trans*-effect sizes. Here, $\boldsymbol{\varepsilon}_i$ is the residual error with residual covariance matrix $\boldsymbol{\Sigma}_{0m}$. The total variance of $\boldsymbol{E}_m$ is

$$\boldsymbol{\Sigma}_m = Var(G_i) \cdot \boldsymbol{\beta}_m \boldsymbol{\beta}_m^\top + \boldsymbol{\Sigma}_{0m},$$

where the genetic component $\boldsymbol{\beta}_m \boldsymbol{\beta}_m^\top$ is rank-1 and $\boldsymbol{\Sigma}_{0m}$ is rank-$q_m$. Therefore, the background-normalized covariance matrix $\boldsymbol{A}_m = \boldsymbol{\Sigma}_{0m}^{-1/2} \widehat{\boldsymbol{\Sigma}}_m \boldsymbol{\Sigma}_{0m}^{-1/2}$ can be written as

$$\boldsymbol{A}_m = \boldsymbol{\Sigma}_{0m}^{-1/2} (Var(G_i) \cdot \boldsymbol{\beta}_m \boldsymbol{\beta}_m^\top + \boldsymbol{\Sigma}_{0m}) \boldsymbol{\Sigma}_{0m}^{-1/2} = Var(G_i) \cdot \left(\boldsymbol{\Sigma}_{0m}^{-1/2} \boldsymbol{\beta}_m\right) \left(\boldsymbol{\Sigma}_{0m}^{-1/2} \boldsymbol{\beta}_m\right)^\top + \mathbf{I}_{q_m},$$

and the eigenvalue of $\boldsymbol{A}_m$ is given by

$$\lambda_{1,m} = 1 + Var(G_i) \cdot \boldsymbol{\beta}_m^\top \boldsymbol{\Sigma}_{0m}^{-1} \boldsymbol{\beta}_m \text{ (signal) and } \lambda_{k,m} = 1 \text{ for } k = 2, \cdots, q_m \text{ (noise)}$$

The corresponding loading direction is proportional to $\boldsymbol{\Sigma}_{0m}^{-1} \boldsymbol{\beta}_m$. Thus, $\lambda_{1,m} > 1$ and $\lambda_{1,m} - 1$ represents genetic variance relative to the residual background in the one genetic signal model.

#### S.4.2.3. Multiple genetic signals model

We next consider $r$ independent *trans*-eQTL affect genes in module $\mathcal{G}_m$, the expression vector $\boldsymbol{E}_m$ follows:

$$\boldsymbol{E}_{i,m} = \sum_{l=1}^{r} \boldsymbol{\beta}_{l,m} G_{i,l} + \boldsymbol{\varepsilon}_i, \boldsymbol{\varepsilon}_i \sim N(\mathbf{0}, \boldsymbol{\Sigma}_{0m}),$$

where $\mathbf{B}_m = (\boldsymbol{\beta}_{1,m}, \cdots, \boldsymbol{\beta}_{r,m}) \in \mathbb{R}^{q_m \times r}$ contain the effects across the module genes. Let $\mathbf{D}_G = cov(\boldsymbol{G}_i)$ be the diagonal when the genetic signals are uncorrelated. Extending the one genetic signal model, the total variance of $\boldsymbol{E}_m$ and the background-normalized covariance matrix are given by

$$\boldsymbol{\Sigma}_m = \mathbf{B}_m \mathbf{D}_G \mathbf{B}_m^\top + \boldsymbol{\Sigma}_{0m}, \text{ and } \boldsymbol{A}_m = \left(\boldsymbol{\Sigma}_{0m}^{-1/2} \mathbf{B}_m \mathbf{D}_G^{1/2}\right)\left(\boldsymbol{\Sigma}_{0m}^{-1/2} \mathbf{B}_m \mathbf{D}_G^{1/2}\right)^\top + \mathbf{I}_{q_m}.$$

Let $\mathbf{C}_m = \boldsymbol{\Sigma}_{0m}^{-1/2} \mathbf{B}_m \mathbf{D}_G^{1/2}$ and $r = rank(\mathbf{C}_m)$. Let $\delta_{1,m} \geq \cdots \geq \delta_{r,m} > 0$ be the nonzero eigenvalues of $\mathbf{C}_m \mathbf{C}_m^\top$ with corresponding eigenvectors $\widehat{\boldsymbol{\omega}}_{m,k}$ for $k = 1, \cdots, r$. Therefore, the eigenvalues of $\boldsymbol{\Sigma}_m$ are

$$\lambda_{k,m} = 1 + \delta_{k,m} \text{ for } k = 1, \cdots, r, \text{ (signal)}$$

$$\lambda_{k,m} = 1 \text{ for } k = r+1, \cdots, q_m. \text{ (noise)}$$

Thus, the number of WPC directions capturing genetic signals, that is, the number of eigenvalues satisfying $\lambda_{k,m} > 1$.

#### S.4.2.4. Data-driven estimation of module-specific background residual covariance matrix

The module-specific background residual covariance matrix, $\boldsymbol{\Sigma}_{0m}$, must be estimated independently of the module covariance being decomposed. If the sample correlation of $\boldsymbol{E}_m$ were used directly, whitening would force $\boldsymbol{E}_m \boldsymbol{\Sigma}_{0m}^{-1/2}$ to have nearly identity empirical covariance and would remove the eigen structure WPC is intended to detect. In the primary implementation, $\boldsymbol{\Sigma}_{0m}$ was estimated from marginal association z-scores computed using a genome-wide set of approximately independent variants, obtained by LD pruning to $r^2 < 0.01$ and excluding xQTL-informed tagging variants used for downstream discovery and evaluation. For a module with $q_m$ genes, let $Z_m \in \mathbb{R}^{p_0 \times q_m}$ denote the variant-by-gene z-score matrix for these pruned variants. Empirical-null variants were defined as

$$\mathcal{N}_m = \left\{ j : \max_{g=1,\cdots,q_m} |Z_{m,jg}| < 2 \right\},$$

and the residual background correlation was estimated as $\widehat{\boldsymbol{\Sigma}}_{0m} = cor(Z_m[\mathcal{N}_m, \cdot])$ [REF:mvSuSiE]. This strategy of estimating residual covariance from independent variants expected to have little or no effect on the analyzed phenotypes in a module.

### S4.3. Global program inference by sparse canonical correlation analysis

In *TransMap-global*, we created 22 LOCO phenotypes after removing variants and genes located at chromosome $c$. For simply notation, we ignored index of chromosome $c$ in this subsection. Let $\boldsymbol{G} \in \mathbb{R}^{n \times p}$ denote the genotype matrix for $p$ variants and $\boldsymbol{E} \in \mathbb{R}^{n \times q}$ denote the donor-level expression matrix for $q$ genes after removal of observed covariates. Sparse canonical correlation analysis (sCCA)[23] seeks sparse linear combinations of variants ($\boldsymbol{u} \in \mathbb{R}^p$) and genes ($\boldsymbol{v} \in \mathbb{R}^q$) that maximize the association between the variant score $\boldsymbol{G}\boldsymbol{u}_k$ and the expression score $\boldsymbol{E}\boldsymbol{v}_k$. For the $k$-th component,

$$(\widehat{\boldsymbol{u}}_k, \widehat{\boldsymbol{v}}_k) = \underset{\boldsymbol{u},\boldsymbol{v}}{\operatorname{argmax}} \{\boldsymbol{u}^\top \boldsymbol{G}^\top \boldsymbol{E} \boldsymbol{v}\}$$
$$\text{subject to} \quad \boldsymbol{u}^\top \boldsymbol{G}^\top \boldsymbol{G} \boldsymbol{u} \leq 1, \quad \boldsymbol{v}^\top \boldsymbol{E}^\top \boldsymbol{E} \boldsymbol{v} \leq 1, \quad \|\boldsymbol{u}\|_1 \leq c_u, \quad \|\boldsymbol{v}\|_1 \leq c_v,$$

where the $L_1$ bounds $c_u$ and $c_v$ control sparsity of the variant and gene loadings. Smaller values retain fewer non-zero entries, whereas $c_u = \sqrt{p}$ or $c_v = \sqrt{q}$ corresponds to the dense boundary under the unit-$L_2$ constraint. The optimization problem of sCCA can also be written in terms of

$$\boldsymbol{W} = \boldsymbol{\Sigma}_G^{-1/2} \boldsymbol{\Sigma}_{GE} \boldsymbol{\Sigma}_E^{-1/2},$$

where $\mathbf{\Sigma}_G$ and $\mathbf{\Sigma}_E$ are genotype and expression covariance matrices, and $\mathbf{\Sigma}_{GE}$ is the cross-covariance matrix between genotype and expression.

We first establish why sCCA objective is formulated using column-centered genotype and expression matrices. Let $\mathbf{1}_n \in \mathbb{R}^n$ be the length-$n$ vector of ones, and define the column-mean vectors and centered matrices as follows:

$$\boldsymbol{\mu}_G = \frac{\boldsymbol{G}^\top \mathbf{1}_n}{n}, \quad \boldsymbol{\mu}_E = \frac{\boldsymbol{E}^\top \mathbf{1}_n}{n}.$$

$$\boldsymbol{G}_{center} = \boldsymbol{G} - \mathbf{1}_n \boldsymbol{\mu}_G^\top, \quad \boldsymbol{E}_{center} = \boldsymbol{E} - \mathbf{1}_n \boldsymbol{\mu}_E^\top.$$

Because the centered columns sum to zero, the uncentered genotype and expression cross-product decomposes as $\boldsymbol{G}^\top \boldsymbol{E}/n = \boldsymbol{G}_{center}^\top \boldsymbol{E}_{center}/n + \boldsymbol{\mu}_G \boldsymbol{\mu}_E^\top$ . The first term is the empirical genotype-expression cross-covariance matrix, $\mathbf{\Sigma}_{GE}$, across donors, whereas the second is a rank-one term determined entirely by marginal means. For sCCA loading vectors $\boldsymbol{u}$ and $\boldsymbol{v}$, respectively, this mean term enters the sCCA objective directly:

$$\boldsymbol{u}^\top (\boldsymbol{G}^\top \boldsymbol{E}/n)\boldsymbol{v} = \mathbf{\Sigma}_{GE} + (\boldsymbol{u}^\top \boldsymbol{\mu}_G)(\boldsymbol{\mu}_E^\top \boldsymbol{v}).$$

Thus, nonzero mean genotype and expression can change the selected loading directions without reflecting genotype and expression cross-covariance. The same distinction applies to the genotype and expression covariance matrices, $\mathbf{\Sigma}_G$ and $\mathbf{\Sigma}_E$. In this covariance-based formulation, both the association term and its normalization describe variation across donors rather than differences in feature means.

We then establish how to estimate $\mathbf{\Sigma}_{GE}$ when only variant-gene association summary statistics are available. Let $\hat{\beta}_{ij}$ be the marginal regression coefficient for centered genotype at variant $i$ and centered expression of gene $j$. Let $\sigma_{G,i}$ and $\sigma_{E,j}$ denote their standard deviations, and define $\widehat{\boldsymbol{B}} = (\hat{\beta}_{ij})$, $\boldsymbol{D}_G = \mathrm{diag}(\sigma_{G,1}, \cdots, \sigma_{G,p})$ and $\boldsymbol{D}_E = \mathrm{diag}(\sigma_{E,1}, \cdots, \sigma_{E,q})$. Because

$$\hat{\beta}_{ij} = \frac{\boldsymbol{G}_{center,i}^\top \boldsymbol{E}_{center,j}}{\boldsymbol{G}_{center,i}^\top \boldsymbol{G}_{center,i}} = \frac{\widehat{\mathbf{\Sigma}}_{GE,ij}}{\sigma_{G,i}^2}, \widehat{\mathbf{\Sigma}}_{GE} = \boldsymbol{D}_G^2 \widehat{\boldsymbol{B}}.$$

If both genotype and expression were standardized before association testing, the effect matrix $\widehat{\boldsymbol{B}}_{std} = \boldsymbol{D}_G \widehat{\boldsymbol{B}} \boldsymbol{D}_E^{-1}$ estimates their marginal association and $\widehat{\mathbf{\Sigma}}_{GE} = \boldsymbol{D}_G \widehat{\boldsymbol{B}}_{std} \boldsymbol{D}_E$. When only z-scores available, $Z_{ij} = \hat{\beta}_{ij} / \widehat{se}(\hat{\beta}_{ij})$, where $\widehat{se}(\hat{\beta}_{ij}) = \hat{s}_{ij} / \sqrt{\boldsymbol{G}_{center,i}^\top \boldsymbol{G}_{center,i}}$ and $\hat{s}_{ij}^2 = \left\| \boldsymbol{E}_{center,j} - \hat{\beta}_{ij} \boldsymbol{G}_{center,i} \right\|^2 / (n-2)$. The residual sum of squared equals $n(\sigma_{E,j}^2 - \hat{\beta}_{ij}^2 \sigma_{G,i}^2)$. Therefore, z-score for the *trans* effect between variant $i$ and gene $j$ can be written as

$$Z_{ij} = \frac{\hat{\beta}_{ij} \sigma_{G,i} \sqrt{n-2}}{\sqrt{\sigma_{E,j}^2 - \hat{\beta}_{ij}^2 \sigma_{G,i}^2}} \Rightarrow \frac{\hat{\beta}_{ij} \sigma_{G,i}}{\sigma_{E,j}} = \frac{Z_{ij}}{\sqrt{n - 2 + Z_{ij}^2}},$$

where $\widehat{\boldsymbol{B}}_{std} = (\hat{\beta}_{ij} \sigma_{G,i} / \sigma_{E,j})$. We can also estimate the cross-covariance matrix from z-scores as $\widehat{\mathbf{\Sigma}}_{GE} = \boldsymbol{D}_G \widehat{\boldsymbol{B}}_{std} \boldsymbol{D}_E$, $\boldsymbol{D}_G$ from LD matrix of genotype and $\boldsymbol{D}_E$ from co-expression matrix of gene expression.

### S5. Identification of gene co-expression modules using WGCNA

Weighted gene co-expression networks were constructed separately for each cell type using the `WGCNA` R package (version 1.74)[24], where genes are connected through correlations among their residualized expression levels. We only considered the protein-coding genes located at autosomes 1-22. WGCNA uses hierarchical clustering to cut the networks into separate gene modules with highly correlated expression levels. We used the default parameter settings. The soft-thresholding power was selected using `pickSoftThreshold` from candidate powers of 1–20, using the default scale-free topology fit threshold of `R2=0.85`. Modules were identified using the hybrid dynamic tree cut algorithm with `deepSplit=4` and `pamRespectsDendro=FALSE`. We only specified the minimum module size parameter (`minModuleSize`) to 10 to obtain small gene modules used in TransPCO[6]. Additionally, to prevent overly large modules from gene hotspots with more than 500 genes, we further divided those hotspots by average linkage hierarchical clustering (`hclust, method=`average``) using the same dissimilarity matrix.

### S6. S-LDSC heritability analysis for trans-MaxPIP annotations

In this study, we highlighted a new idea to evaluate whether variants implicated by single gene *trans* fine-mapping inform disease risk, creating an annotation for fine-mapped *trans*-eQTL. We constructed a new variant-level *trans*-acting annotation based on the *maximum posterior inclusion probability* (*trans*-MaxPIP) from genome-wide fine-mapping for each resolution of *trans*-acting phenotype. For each *trans*-acting phenotype $y$, such as individual *trans*-eGenes, local WPC-based program scores or global sCCA-based program scores, let $\text{PIP}_j^y$ denote the PIP from fine-mapping of variant $j$ to *trans*-phenotype $y$. Let $\mathcal{H}$ denote the set of *trans*-phenotypes whose 95% CS contains variant $j$, we then defined

$$\textit{trans}\text{-MaxPIP}_j = \max_{y \in \mathcal{H}} \text{PIP}_j^y,$$

with variants do not present in any credible set assigned a score of zero. Separate *trans*-MaxPIP annotations were generated for *TransMap-eGene*, *TransMap-local* and *TransMap-global*, and analogous annotations were generated for competing methods where appropriate. Except for the cell-type-resolved variant annotation from each scale, we also generated cross-cell-types annotations by taking maximum value of *trans*-$\text{MaxPIP}_j$ for variant $j$.

We applied the stratified LD score regression (S-LDSC) to estimate per-SNP heritability, enrichment, and standardized effect size of *trans*-MaxPIP annotation[25,26]. In S-LDSC analysis, we modeled the disease trait of interest, $Y_{\text{disease}}$, as depending linearly on genotype, $Y_{\text{disease}} = X\beta + \epsilon$. Because each variant is standardized, S-LDSC assumes the variance of effect size, $\text{Var}(\beta_j)$, is a linear additive contribution to annotations. We also can call $\text{Var}(\beta_j)$ the per-variant heritability of variant $j$. Denote $h_g^2 = \sum_j \text{Var}(\beta_j)$ as the estimated variant heritability across all annotations, then $h_g^2/M$ represents the proportion of heritability explained by per-variant within all annotations, where $M$ is the total number of variants.

Given $C = 53$ functional annotations from the baseline model[21,26], including binary annotation, conserved, and regulatory annotations (e.g., promoter, enhancer, histone marks, TF binding sites). Let $a_{j,trans}$ and $a_{j,c}$ be the annotation value of variant $j$ based on *trans*-MaxPIP from *TransMap* and functional annotation $c$ from baseline LD model for $c = 1, \ldots, C$, respectively. The per-variant heritability of variant $j$ is given by

$$\text{Var}(\beta_j) = a_{j,trans} \cdot \tau_{trans} + \sum_c a_{j,c} \cdot \tau_c,$$

where $\tau_{trans}$ and $\tau_c$ quantify heritability explained by per-variant within *trans*-MaxPIP annotation and annotation $c$, respectively. We can estimate $\tau_{trans}$ under a linear model between association test statistics and LD score as follows,

$$\mathbb{E}[\chi_j^2] = N\left(\ell(j,trans) \cdot \tau_{trans} + \sum_c \ell(j,c)\tau_c\right) + 1,$$

where $\ell(j,c)$ is LD score of the variant $j$ to annotation $c$, defined as $\ell(j,c) = \sum_k a_{k,c}\, r_{jk}^2$ with $r_{jk}$ is correlation between variants $j$ and $k$. Similar definition of $\ell(j,trans) = \sum_k a_{k,trans}\, r_{jk}^2$. Enrichment is defined as the relative contribution of heritability explained by *trans*-MaxPIP annotation,

$$\text{Enrichment} = \frac{\tau_{trans}}{h_g^2/M},$$

where $\tau_{trans} = h^2_{g,trans}/\sum_j a_{j,trans}$ quantifies heritability explained by per-variant within *trans*-MaxPIP annotation, correcting for 53 functional annotations. Enrichment$>1$ represents *trans*-MaxPIP annotation enriched to disease heritability. Standardized effect size $\tau^*_{trans}$ is defined as the proportionate change in per-variant heritability associated with one standard deviation increase in the value of *trans*-MaxPIP annotation, conditional on other annotations included in the model. $\tau^*_{trans} > 0$ if *trans*-MaxPIP annotation increases per-variant heritability, accounting for all other annotations included. Both enrichment and $\tau^*_{trans}$ are estimated via a genomic block-jackknife procedure with 200 blocks that also provides standard errors[26].

We considered two versions of $\tau^*_{trans}$: a marginal version conditional on baseline annotations only, and a joint version where the conditioning set includes other *trans*-MaxPIP derived annotations in addition to baseline. In the joint analysis, we use a metric, joint $\tau^*$, to quantify the conditional informativeness of a heritability model[26], generalizing the joint $\tau^*$ metric to more than two annotations. In detail, given a joint model defined by $C$ annotations (conditional on a published set of annotations such as the baseline model), we use

$$\text{Joint } \tau^* = \sqrt{\sum_{c=1}^{C} (\tau_c^*)^2 + \sum_{c \neq l} r_{cl}\, \tau_c^* \tau_l^*},$$

where $r_{cl}$ is the pairwise correlation of the annotations $c$ and $l$, and $r_{cl}\tau_c^*\tau_l^*$ is expected to be positive since two positively correlated annotations typically have the same direction of effect[26]. We calculated standard errors for joint $\tau^*$ using a genomic block-jackknife with 200 blocks.
